# Social comparisons on social media and mental health: A systematic review and meta-analysis

**DOI:** 10.64898/2026.09.15.26363124

**Authors:** Thole H. Hoppen, Lotta Höfer, Pascal Schlechter, Heinz Holling, Julian Mutz, Nexhmedin Morina

## Abstract

Frequent comparisons with idealized portrayals of others on social media may be linked to poorer mental health. To test this hypothesis, we systematically searched PsycInfo, MEDLINE, and Web of Science for studies published through April 28, 2026. A total of 149 observational studies (*N* = 76,262) and 77 experimental studies (*N* = 15,404) were included in meta-analytic syntheses. Upward social comparison tendency correlated cross-sectionally with mental health outcomes, including psychopathology (*r* = .32) and body dissatisfaction (*r* = .54). Worse comparative self-evaluations were associated with poorer mental health and vice versa, with moderate-to-large effect sizes. Across experimental studies, upward social comparisons adversely affected mental health. Results were robust in sub-group analyses (e.g., female or young samples). Across study designs, there was no evidence that downward comparisons were linked to mental health. These findings identify upward social comparisons as a robust correlate and potential risk factor for poor mental health.

## Introduction

Over five billion people worldwide use social media, spending more than two hours per day on average, with even higher usage in younger people^1^. In some studies, social media use has been associated with worse mental health, yet results are mixed, particularly among adolescents^2–10^. One explanation is that how individuals engage with social media may matter more for psychological outcomes than time spent on it^9,11–16^.

Social comparison on social media has been proposed as a key mechanism through which social media use may influence mental health^12,17,18^. Social comparison is a ubiquitous process by which individuals evaluate themselves relative to others on attributes such as appearance, competence, or well-being. Since Festinger’s original formulation of social comparison theory^19^, research has shown that upward social comparisons (i.e., comparing to others that are perceived as better-off) in particular are associated with poor mental health^17,18,20–22^.

Social media has fundamentally changed the opportunities for social comparison^23^. Unlike offline interactions, social media feeds often feature carefully curated portrayals of others’ lives (e.g., accounts of influencers), increasing both upward comparison frequency and perceived discrepancies from upward standards^12^. Numerous studies have examined associations between social comparisons on social media and mental health and existing meta-analyses^4,11,24–28^ have focused on a single study design (e.g., experimental studies), population (e.g., adolescents), component of social comparison (e.g., social comparison tendency), or a specific mental health outcome (e.g., depression, pathological eating, or body image). However, a comprehensive and up-to date meta-analysis covering various study designs, populations, mental health indicators, and components of the social comparison process (e.g., social comparison tendency or frequency on social media, or affective responses to social comparison) is needed.

Crucially, such a broad synthesis can identify whether naturally occurring and experimentally induced social comparisons on social media provide converging findings across mental health outcomes, or whether population-specific or component-specific patterns emerge.

Here, we provide such a comprehensive meta-analysis. We conceptualized social comparison as a process involving: a) seeking or encountering social information, b) evaluating target-standard discrepancies, c) appraising the comparison outcome in relation to the individual’s motives and coping resources, and d) resulting emotional, cognitive, and behavioral responses (see below)^29^. In doing so, we focused on both experimental methods assessing cognitive, emotional and behavioral reactions to induced social comparison and observational methods assessing naturally occurring social comparison through self-report. We further conceptualized mental health as comprising both unfavorable (e.g., psychopathology and body dissatisfaction) and favorable outcomes (e.g., life satisfaction and positive affect)^3,30–32^. Finally, we examined whether findings differ across mental health indicators, sociodemographic factors (i.e., age and gender), or social media platforms, all of which have been proposed as potential moderators^24,25^. Following PRISMA guidelines^33^, we addressed two questions: (1) how are naturally occurring social comparisons on social media associated with mental health in observational studies, and (2) how do experimentally induced social comparisons on social media affects mental health outcomes.

## Results

### Search results and study characteristics

The systematic literature search yielded 18,094 records in PsycInfo and MEDLINE in the combined search via EBSCOhost and 16,123 in Web of Science. Following duplicate removal and screening of titles and abstracts, 1,016 full texts were screened thoroughly for eligibility. In total, 817 full texts were excluded (Figure 1). Eighteen articles^34–50^ and 14 articles^51–64^ reported experimental and observational data on social comparison on social media, respectively, but in insufficient detail, and corresponding authors did not respond to our data requests. A total of 17 publications^65–81^ reported ecological momentary assessment (EMA) studies investigating social comparison on social media and five publications^82–86^ reported intervention studies aimed at alleviating the adverse effects of social media usage, which fell outside the scope of the present review.

**Figure 1.**
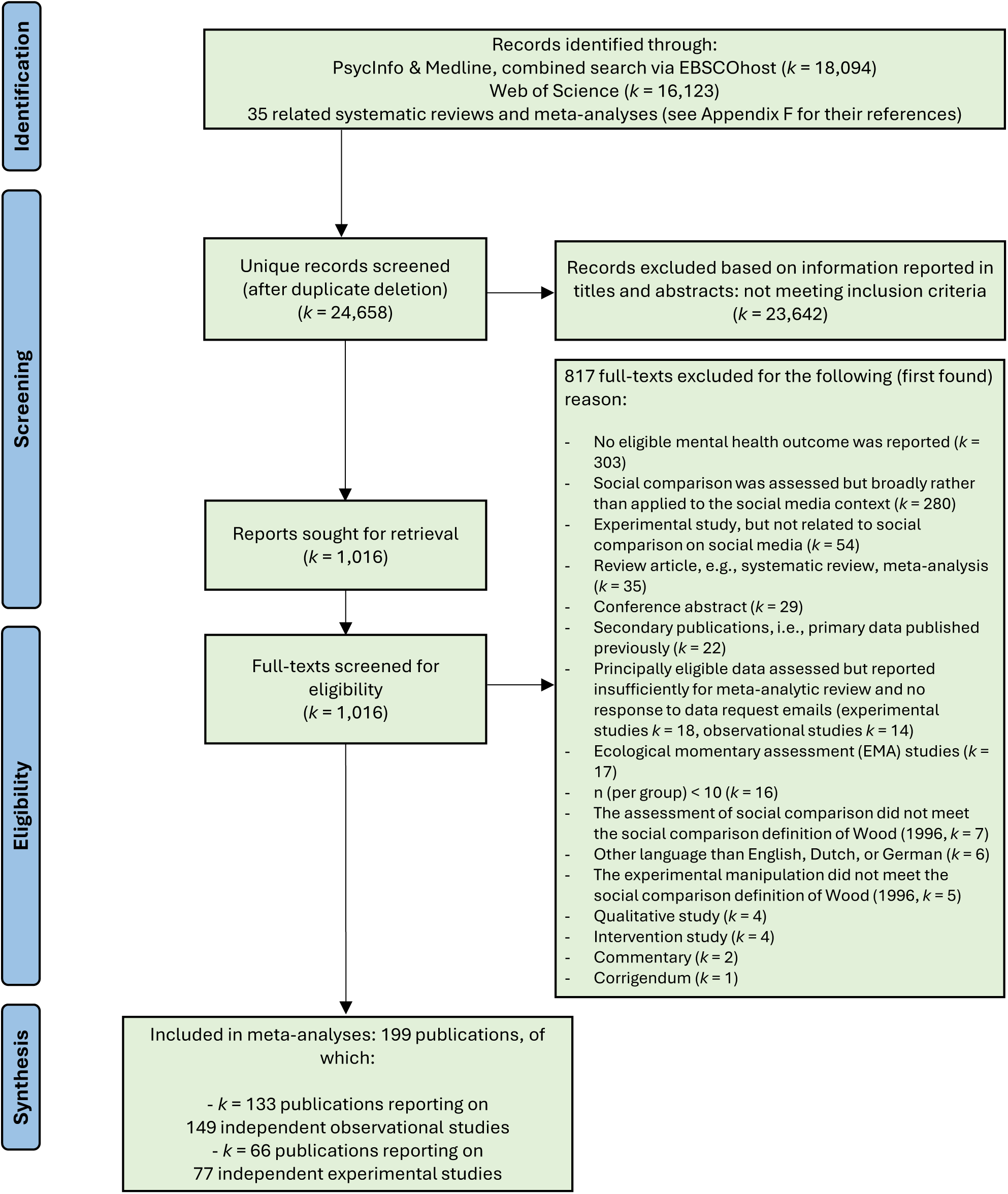
PRISMA flow chart depicting the study synthesis

A total of 199 eligible publications reporting on 228 independent studies (*N* = 91,666) were included in the present meta-analysis. Of these, *k* = 133 publications^44,87–218^ reported on 149 independent observational studies. We received data for 19 of these studies from the corresponding authors. One publication^44^ reported both an observational and an experimental study, but the data were only sufficiently reported for the observational study. While almost all observational studies (146 out of 149, 98%) reported bivariate correlations between a social comparison component (e.g., tendency) and a mental health outcome (e.g., depression), four studies^89,125,148,180^ (3%) reported standardized mean differences between individuals with and without clinical diagnosis. One study^125^ reported correlations and standardized mean differences. A total of *k* = 66 articles^219–284^ reported on 77 independent experimental studies. We received data for 10 of these studies from the corresponding authors.

The 149 independent observational studies comprised 76,262 participants, with sample sizes ranging from 26 to 2,616. See Supplementary Appendix A for the characteristics of all observational studies. The unweighted mean age across studies was 22.87 years (SD = 3.95), and 70% of participants were female. In 45 studies (30% of all observational studies), only female participants were included. Observational studies were conducted in 22 different countries from four continents, with most studies conducted in China (*k* = 34), the US (*k* = 33), and Australia (*k* = 16). Study quality was mixed. Only 44 of 149 observational studies (29%) assessed a social comparison component (e.g., social comparison tendency, social comparison-based self-evaluation, social rank, affective response) with a validated measure, 71 studies (48%) with a partly validated measure, and 34 studies (23%) with an unvalidated measure (e.g., self-construed, unvalidated items). Less than half of the studies (46%, 68 of 149) assessed the respective social comparison component with a clear social comparison direction (e.g., upward social comparison), whereas the remaining studies (i.e., 81 studies, 54%) did not. Most of the 146 observational studies assessing correlations (i.e., 93% or 136), assessed the given mental health outcome with a validated measure, whereas 4 (3%) studies used a partially validated measure, and six studies (4%) an unvalidated measure. Three observational studies only provided standardized mean differences between individuals with and without clinical diagnosis.

The 77 independent experimental studies comprised 15,404 participants, with sample sizes ranging from 49 to 625. See Supplementary Appendix B for the characteristics of all experimental studies. The unweighted mean age across studies was 22.03 years (SD = 3.04), and 89% of participants were female. In 57 studies (74% of the 77 studies) only females were included. Experimental studies were conducted in 10 different countries from four continents, with most studies conducted in Australia (*k* = 23), the US (*k* = 16), and the UK (*k* = 9). Study quality was high, with all studies using validated measures to assess mental health outcomes. Similarly, 97% (37 of 38) of the studies that aimed to manipulate the social comparison direction (i.e., upward standard, downward standard, lateral standard) could be clearly categorized in terms of social comparison direction. Experimental studies that did not aim to manipulate the social comparison direction involved positive body image manipulations and thus the given quality criterion was not applicable.

A summary and illustration of the main meta-analytic results and conclusions across observational and experimental studies is depicted in Figure 2. Forest and funnel plots for each meta-analysis can be reproduced by means of open code and data accessible on Zenodo (will be made available once published).

**Figure 2.**
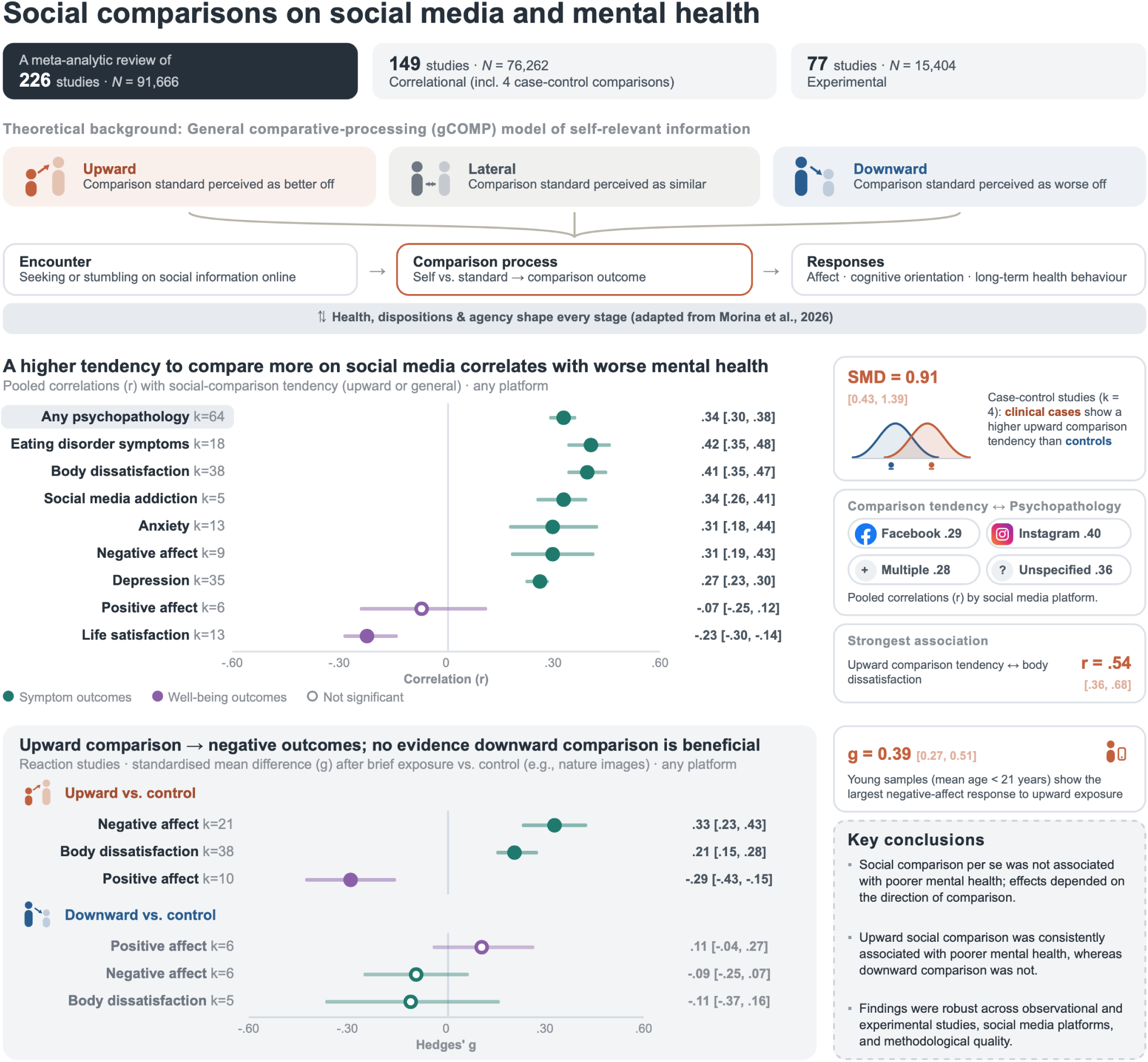
Main results and conclusions of meta-analytic review

### Cross-sectional and longitudinal bivariate correlations

The main meta-analytic results of the 149 observational studies are shown in Table 1, whereas platform-specific results are shown in Supplementary Appendix C. Across 64 independent studies assessing cross-sectional correlations, the tendency to compare with others on social media (studies assessing upward or non-directional social comparison tendency analyzed jointly) correlated positively with the severity of various psychopathology outcomes, with a moderate effect size (*r* = .34 (95% CI .30; .38), *P* < .001), yet with high between-study heterogeneity (*τ* = 0.18; *I*^2^ = 94.66%). Visual inspection of the funnel plot revealed limited asymmetry, and Egger’s test did not identify small study effects. No outlier was detected. The 95% prediction interval included the null (-.01; .62). Results were similar when psychopathologies (i.e., depression, eating disorders, anxiety disorders, and social media addiction) were analyzed individually. Some 95% prediction intervals excluded the null, indicating that the true effects in 95% of comparable future studies would be expected to fall in the same direction as the pooled effect. Upward or non-directional social comparison tendency also correlated positively with body dissatisfaction (*r* = .41 (.35; .47); *P* < .001; *k* = 38; *τ* = 0.22; *I*^2^ = 95.71%) and negative affect (*r* = .31 (.19; .43); *P* < .001; *k* = 9; *τ* = 0.19; *I*^2^ = 92.68%), negatively with life satisfaction (*r* = -.23 (-.30; -.14); *P* < .001; *k* = 13; *τ* = 0.14; *I*^2^ = 93.81%). Heterogeneity was generally high and no outliers or significant asymmetry (Egger’s test) were observed. Results remained similar in sub-group analyses examining female samples only, young samples only (i.e., sample mean age ≤ 21 years), and high-quality studies only (i.e., studies with quality sum score ≥ 7/8). Results were also similar when appearance-related social comparison tendency was analyzed and when upward social comparison tendency and non-directional social comparison tendency were analyzed separately. No statistically significant differences in the strength of associations by social media platform were found in most moderator analyses, with one exception (Supplementary Appendix C). There was some evidence that the associations between social comparison tendency and body dissatisfaction were stronger for Instagram relative to Facebook. Publication year and mean sample age were not found to be significant predictors of strength of association (see Table 2 for detailed results concerning all meta-regressions for observational studies).

**Table 1.**
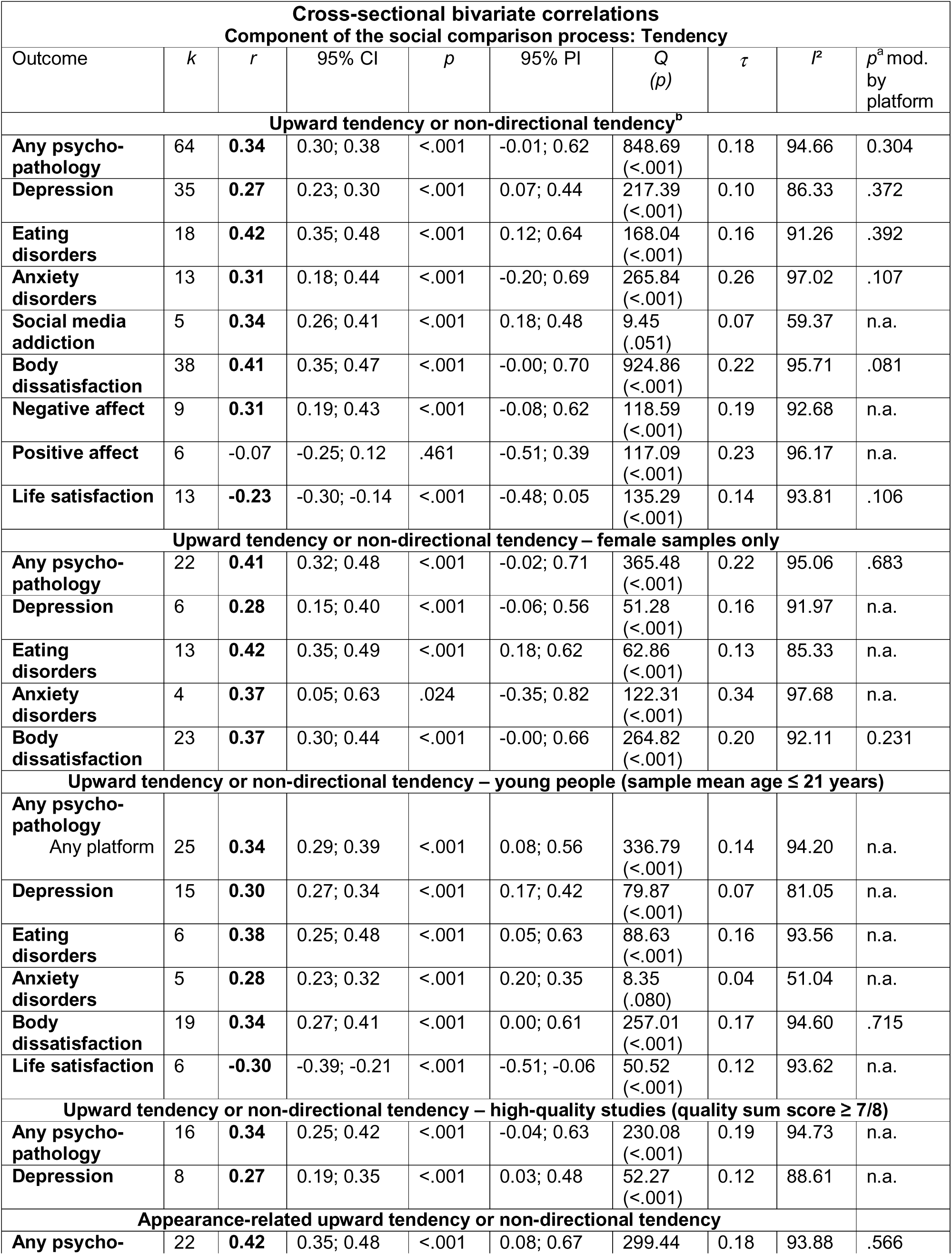
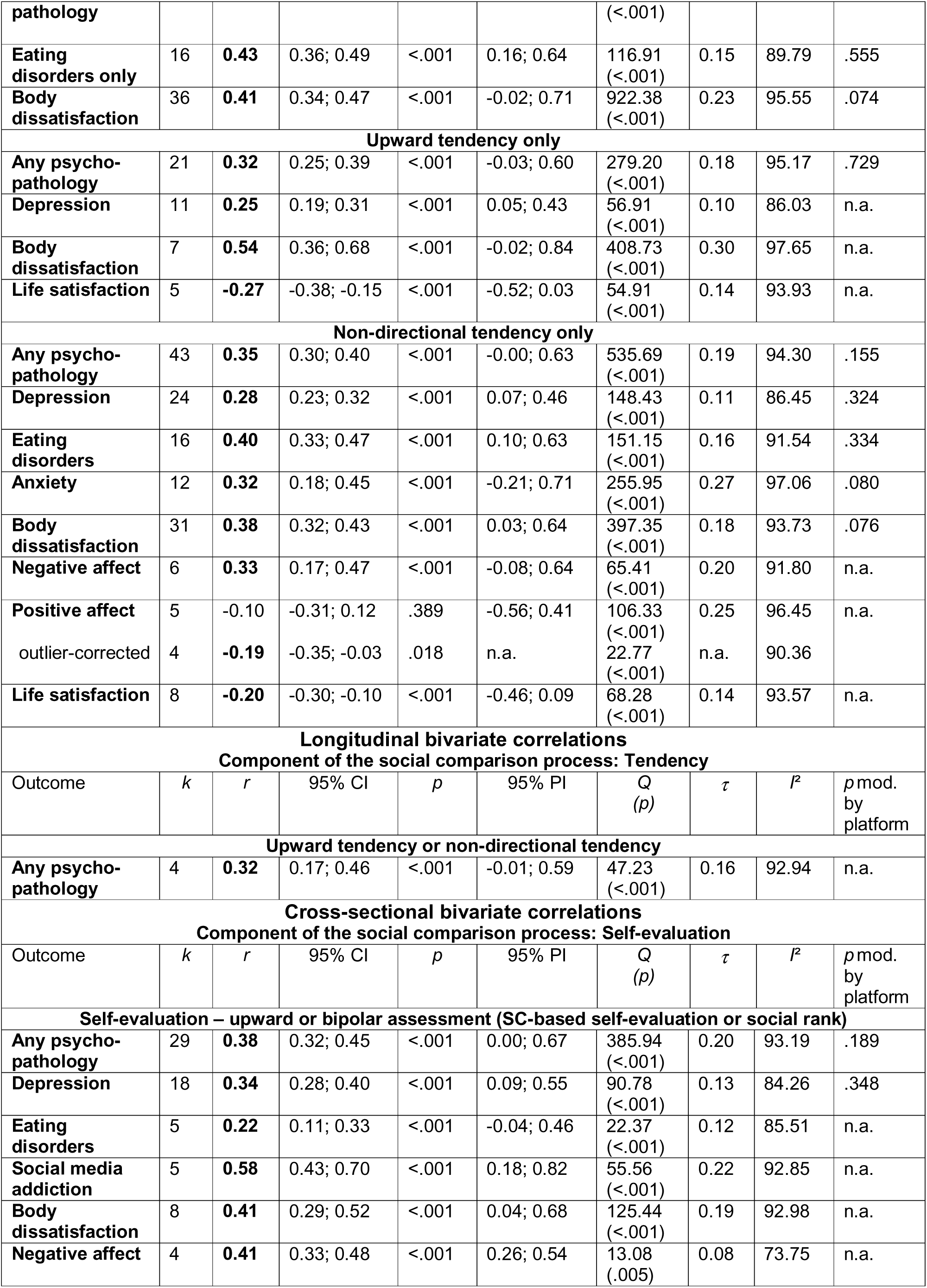
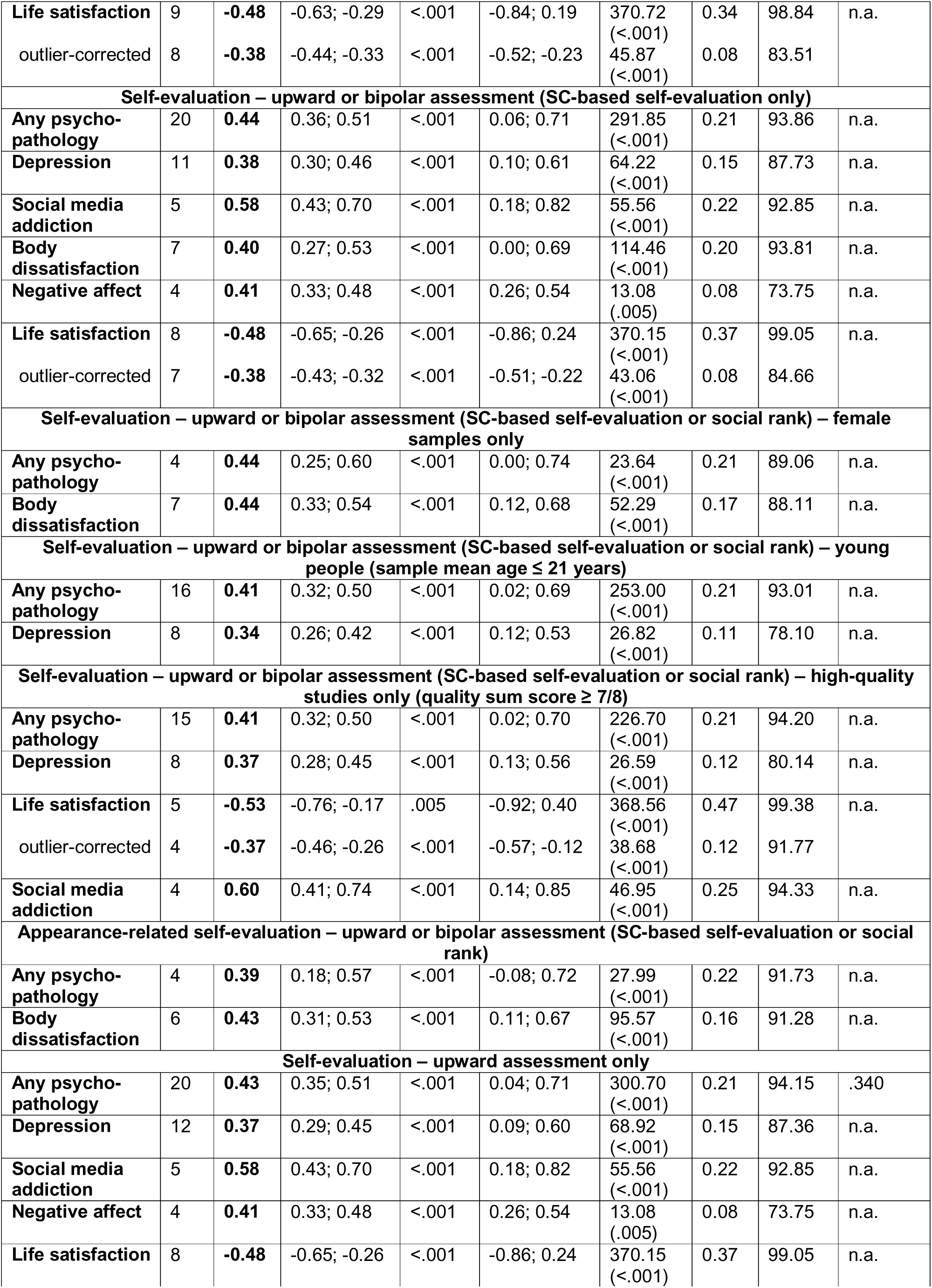
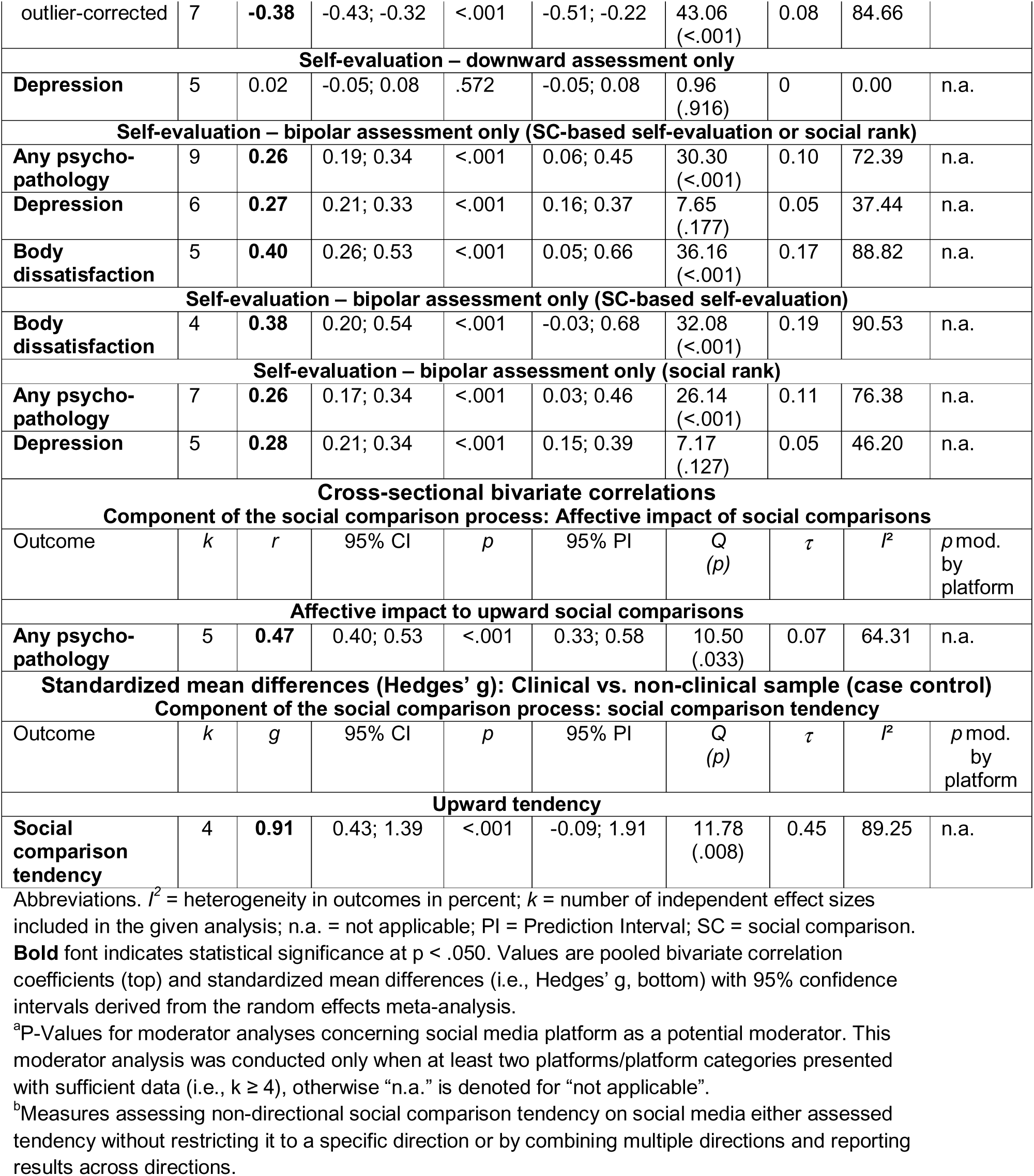
Meta-analytic results from observational studies assessing the relationship between social comparison on social media and mental health outcomes.

**Table 2.** Meta-regression results concerning publication year and mean age as potential moderators of correlations reported in observational studies.

| Cross-sectional bivariate correlations |  |  |  |  |  |  |  |
| --- | --- | --- | --- | --- | --- | --- | --- |
| Component of the social comparison process: Tendency |  |  |  |  |  |  |  |
| Outcome | Moderator | *k* | *b* | 95% CI | *p* | Res. $\tau$ | Res. *I*<sup>2</sup> |
| Upward tendency or non-directional tendency<sup>a</sup> |  |  |  |  |  |  |  |
| Any psycho-pathology | Publication year | 64 | 0.006 | -0.009; 0.021 | .428 | 0.18 | 94.62 |
| Depression |  | 35 | 0.005 | -0.007; 0.017 | .383 | 0.10 | 85.85 |
| Eating disorder |  | 18 | -0.007 | -0.036; 0.021 | .610 | 0.16 | 91.11 |
| Anxiety |  | 13 | 0.038 | -0.013; 0.088 | .142 | 0.25 | 96.41 |
| Body dissatisfaction |  | 38 | 0.014 | -0.014; 0.042 | .337 | 0.22 | 95.62 |
| Life satisfaction |  | 13 | -0.010 | -0.042; 0.021 | .525 | 0.14 | 94.01 |
| Any psycho-pathology | Mean age | 53 | -0.001 | -0.006; 0.005 | .789 | 0.14 | 91.80 |
| Depression |  | 27 | -0.004 | -0.009; 0.001 | .109 | 0.07 | 79.02 |
| Eating disorder |  | 17 | 0.001 | -0.008; 0.010 | .846 | 0.14 | 89.57 |
| Body dissatisfaction |  | 34 | 0.006 | -0.001; 0.014 | .077 | 0.18 | 93.69 |
| Life satisfaction |  | 12 | **0.013** | 0.007; 0.018 | <.001 | 0.07 | 79.29 |
| Cross-sectional bivariate correlations |  |  |  |  |  |  |  |
| Component of the social comparison process: Self-evaluation |  |  |  |  |  |  |  |
| Outcome | Moderator | *k* | *b* | 95% CI | *p* | Res. $\tau$ | Res. *I*<sup>2</sup> |
| Self-evaluation – upward or bipolar assessment (SC-based self-evaluation or social rank) |  |  |  |  |  |  |  |
| Any psycho-pathology | Publication year | 29 | 0.011 | -0.012; 0.034 | .341 | 0.20 | 93.18 |
| Depression |  | 18 | -0.001 | -0.021; 0.018 | .902 | 0.13 | 84.88 |
| Any psycho-pathology | Mean age | 28 | -0.004 | -0.018; 0.011 | .642 | 0.20 | 93.26 |
| Depression |  | 17 | 0.006 | -0.005; 0.017 | .298 | 0.12 | 82.62 |

Abbreviations. $I^2$ = heterogeneity in outcomes in percent; *k* = number of independent effect sizes included in the given random-effects meta-analysis.
<sup>a</sup>Measures assessing non-directional social comparison tendency, either without specifying direction or by combining multiple comparison directions.

Across four independent studies assessing longitudinal correlations, upward or non-directional social comparison tendency correlated positively with psychopathology outcomes assessed at a later timepoint (*r* = .32 (.17; .46); *P* < .001; *τ* = 0.16; *I*^2^ = 92.94). The time periods between assessments varied widely (i.e., 1 week, 2 months, 3 months, and 24 months). Too few studies (*k* < 4) examined downward social comparison tendency.

Across 29 independent studies assessing cross-sectional correlations, the social comparison-based self-evaluation/social rank (using either upwardly directed scales or bipolar scales ranging from lower to higher social standing) correlated positively with the severity of various psychopathology outcomes, with a moderate effect size (*r* = .38 (.32; .45), *P* < .001). High between-study heterogeneity was found (*τ* = 0.20; *I*^2^ = 93.19%). A visual inspection of the funnel plot revealed limited asymmetry, and Egger’s test did not identify small study effects. No outlier was detected. Results were similar when specific psychopathologies (i.e., depression, eating disorders, social media addiction) were analyzed. Social comparison-based self-evaluation/social rank also correlated positively with body dissatisfaction (*r* = .41 (.29; .52); *P* < .001; *k* = 8; *τ* = 0.19; *I^2^* = 92.98%) and negative affect (*r* = .41 (.33; .48); *P* < .001; *k* = 4; *τ* = 0.08; *I^2^* = 73.75%), and negatively with life satisfaction (*r* = -.48 (-.63; - .29); *P* < .001; *k* = 9; *τ* = 0.34; *I^2^* = 98.84%). Heterogeneity was generally high and no asymmetry was observed in these analyses. There were also no detected outliers, except in the analysis concerning satisfaction with life. Results remained similar after excluding this outlier (*r* = -.38 (-.44; -.33); *P* < .001; *k* = 8; *τ* = 0.08; *I^2^* = 83.51%). Results also remained similar in sub-group analyses examining female samples only, young samples only, high-quality studies only, and social comparison-based self-evaluation only (i.e., social rank data excluded). When appearance-related self-evaluation, upwardly directed and bipolar assessments were analyzed separately, results also remained similar. Again, no significant differences in the strength of associations by social media platform were found in the moderator analyses (Supplementary Appendix C). Additionally, there was no statistically significant association between self-evaluations in the downward direction and depression severity (*r* = .02 (-.05; .08); *P* = .572; *k* = 5), with no heterogeneity between studies (*τ* = 0.00; *I^2^* = 0.00%). Data on other mental health outcomes or longitudinal data on self-evaluations were too scarce for analysis (*k* < 4).

Across five independent studies assessing cross-sectional correlations, the self-reported affective response to online social comparison correlated positively with the severity of various psychopathology outcomes, with a moderate-to-large effect size (*r* = .47 (.40; .53), *P* < .001), indicating a positive association between psychopathology and the affective impact of social comparison. Moderate between-study heterogeneity was found (*τ* = 0.07; *I^2^* = 64.31%). Visual inspection of the funnel plot and Egger’s test were precluded (*k* < 10). No outlier was detected.

Across four independent case control studies assessing standardized mean differences, the self-reported upward social comparison tendency was, on average, higher in the clinical vs. non-clinical groups, with a large effect size (*g* = 0.91 (0.43; 1.39), *P* < .001). High between-study heterogeneity was found (*τ* = 0.45; *I^2^* = 89.25%). Visual inspection of the funnel plot and Egger’s test were precluded (*k* < 10). No outlier was detected.

### Effects of social comparisons on social media on mental health in experimental studies

The main meta-analytic results concerning the 77 experimental studies are reported in Table 3, whereas platform-specific results are shown in Supplementary Appendix D. Across 21 experimental studies, upward social comparisons (e.g., exposure to pictures of successful influencers on social media) relative to control conditions (e.g., exposure to landscape images on social media) increased negative affect, with a moderate effect size (Hedges’ *g* = 0.33 (0.23;0.43), *P* < .001) and with low-to-moderate between-study heterogeneity (*τ* = 0.14; *I^2^* = 35.74%). Visual inspection of the funnel plot revealed limited asymmetry, and Egger’s test did not identify small study effects. No outlier was detected. The 95% prediction interval indicated that the true effects in 95% of comparable future studies would be expected to fall in the same direction as the pooled effect. Upward social comparisons relative to control conditions also increased body dissatisfaction (Hedges’ *g* = 0.21 (0.15; 0.28), *P* < .001, *k* = 38) and reduced positive affect (Hedges’ *g* = -0.29 (-0.43; -0.15), *P* < .001, *k* = 10). Heterogeneity was small in both analyses, and no outliers or significant asymmetry (Egger’s test) were observed. Results remained similar in sub-group analyses examining female samples only and young samples only. Sub-group analyses restricted to appearance-based social comparison, or high-quality studies were not conducted for the experimental studies given that all included studies were of high-quality and manipulated appearance-based social comparison. No significant differences in the strength of effects by social media platform were found in moderator analyses, with one exception. Exposure to Fitspiration content on Instagram had a significantly larger effect on body dissatisfaction than Fitspiration content on TikTok relative to control conditions, respectively (*P* = .048; Supplementary Appendix D). Publication year and mean age of sample were also not found to be significant predictors of effect sizes in most analyses, with one exception. Younger mean age of sample was associated with larger effects of upward social comparison on body dissatisfaction (*P* = .004). See Table 4 for detailed results concerning all meta-regressions for experimental studies.

**Table 3.**
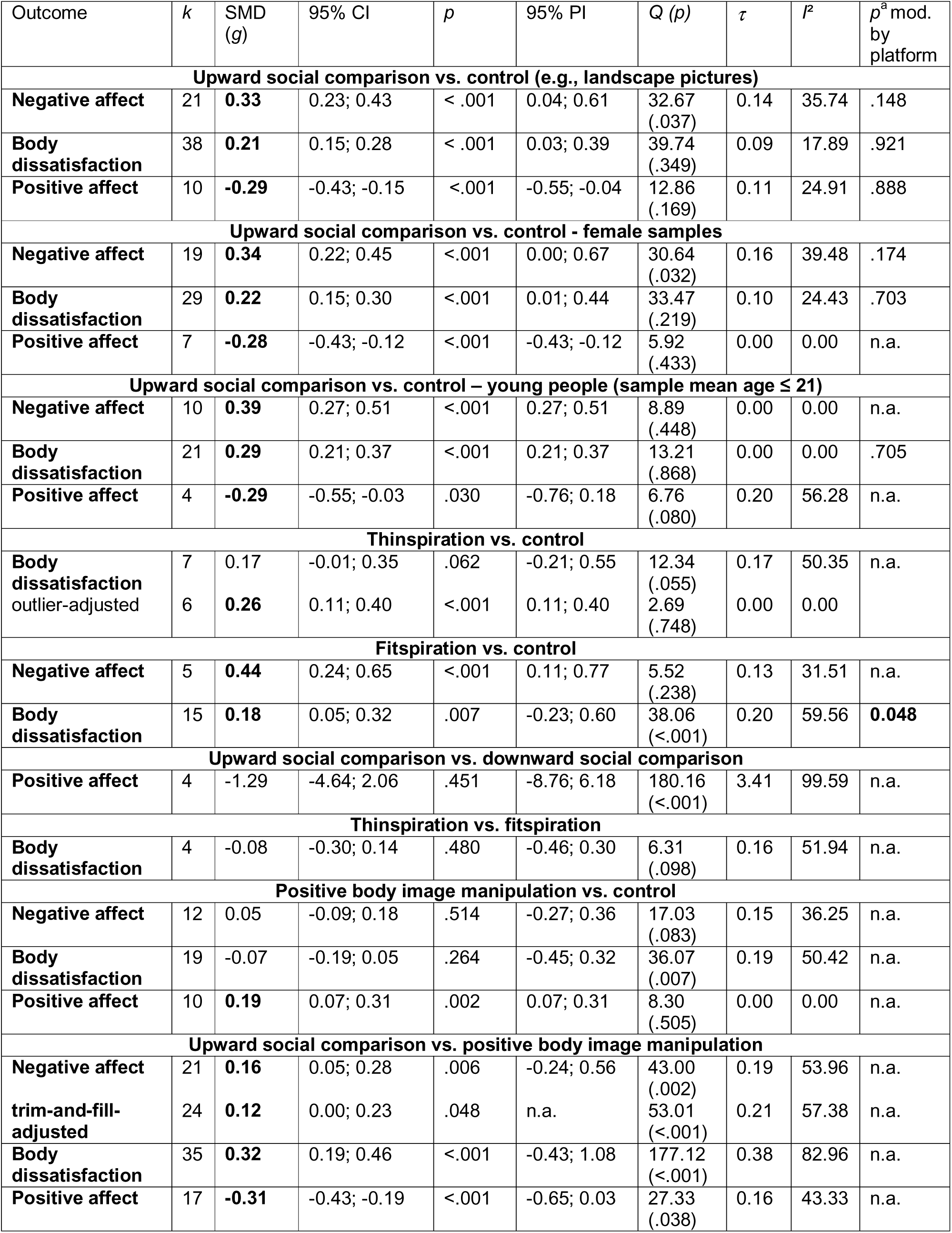
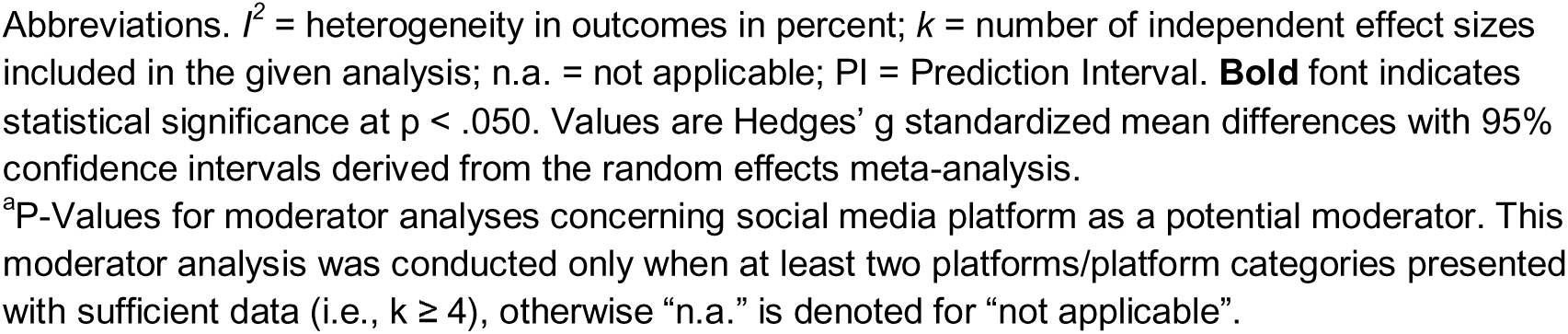
Meta-analytic results for experimental studies assessing the effects of social comparison on social media on mental health outcomes.

| Outcome | *k* | SMD (*g*) | 95% CI | *p* | 95% PI | *Q* (*p*) | $\tau$ | *I*<sup>2</sup> | *p*<sup>a</sup> mod. by platform |
| --- | --- | --- | --- | --- | --- | --- | --- | --- | --- |
| **Upward social comparison vs. control (e.g., landscape pictures)** |  |  |  |  |  |  |  |  |  |
| Negative affect | 21 | **0.33** | 0.23; 0.43 | < .001 | 0.04; 0.61 | 32.67 (.037) | 0.14 | 35.74 | .148 |
| Body dissatisfaction | 38 | **0.21** | 0.15; 0.28 | < .001 | 0.03; 0.39 | 39.74 (.349) | 0.09 | 17.89 | .921 |
| Positive affect | 10 | **-0.29** | -0.43; -0.15 | < .001 | -0.55; -0.04 | 12.86 (.169) | 0.11 | 24.91 | .888 |
| **Upward social comparison vs. control - female samples** |  |  |  |  |  |  |  |  |  |
| Negative affect | 19 | **0.34** | 0.22; 0.45 | < .001 | 0.00; 0.67 | 30.64 (.032) | 0.16 | 39.48 | .174 |
| Body dissatisfaction | 29 | **0.22** | 0.15; 0.30 | < .001 | 0.01; 0.44 | 33.47 (.219) | 0.10 | 24.43 | .703 |
| Positive affect | 7 | **-0.28** | -0.43; -0.12 | < .001 | -0.43; -0.12 | 5.92 (.433) | 0.00 | 0.00 | n.a. |
| **Upward social comparison vs. control – young people (sample mean age ≤ 21)** |  |  |  |  |  |  |  |  |  |
| Negative affect | 10 | **0.39** | 0.27; 0.51 | < .001 | 0.27; 0.51 | 8.89 (.448) | 0.00 | 0.00 | n.a. |
| Body dissatisfaction | 21 | **0.29** | 0.21; 0.37 | < .001 | 0.21; 0.37 | 13.21 (.868) | 0.00 | 0.00 | .705 |
| Positive affect | 4 | **-0.29** | -0.55; -0.03 | .030 | -0.76; 0.18 | 6.76 (.080) | 0.20 | 56.28 | n.a. |
| **Thinspiration vs. control** |  |  |  |  |  |  |  |  |  |
| Body dissatisfaction | 7 | 0.17 | -0.01; 0.35 | .062 | -0.21; 0.55 | 12.34 (.055) | 0.17 | 50.35 | n.a. |
| outlier-adjusted | 6 | **0.26** | 0.11; 0.40 | < .001 | 0.11; 0.40 | 2.69 (.748) | 0.00 | 0.00 |  |
| **Fitspiration vs. control** |  |  |  |  |  |  |  |  |  |
| Negative affect | 5 | **0.44** | 0.24; 0.65 | < .001 | 0.11; 0.77 | 5.52 (.238) | 0.13 | 31.51 | n.a. |
| Body dissatisfaction | 15 | **0.18** | 0.05; 0.32 | .007 | -0.23; 0.60 | 38.06 (<.001) | 0.20 | 59.56 | **0.048** |
| **Upward social comparison vs. downward social comparison** |  |  |  |  |  |  |  |  |  |
| Positive affect | 4 | -1.29 | -4.64; 2.06 | .451 | -8.76; 6.18 | 180.16 (<.001) | 3.41 | 99.59 | n.a. |
| **Thinspiration vs. fitspiration** |  |  |  |  |  |  |  |  |  |
| Body dissatisfaction | 4 | -0.08 | -0.30; 0.14 | .480 | -0.46; 0.30 | 6.31 (.098) | 0.16 | 51.94 | n.a. |
| **Positive body image manipulation vs. control** |  |  |  |  |  |  |  |  |  |
| Negative affect | 12 | 0.05 | -0.09; 0.18 | .514 | -0.27; 0.36 | 17.03 (.083) | 0.15 | 36.25 | n.a. |
| Body dissatisfaction | 19 | -0.07 | -0.19; 0.05 | .264 | -0.45; 0.32 | 36.07 (.007) | 0.19 | 50.42 | n.a. |
| Positive affect | 10 | **0.19** | 0.07; 0.31 | .002 | 0.07; 0.31 | 8.30 (.505) | 0.00 | 0.00 | n.a. |
| **Upward social comparison vs. positive body image manipulation** |  |  |  |  |  |  |  |  |  |
| Negative affect | 21 | **0.16** | 0.05; 0.28 | .006 | -0.24; 0.56 | 43.00 (.002) | 0.19 | 53.96 | n.a. |
| trim-and-fill-adjusted | 24 | **0.12** | 0.00; 0.23 | .048 | n.a. | 53.01 (<.001) | 0.21 | 57.38 | n.a. |
| Body dissatisfaction | 35 | **0.32** | 0.19; 0.46 | < .001 | -0.43; 1.08 | 177.12 (<.001) | 0.38 | 82.96 | n.a. |
| Positive affect | 17 | **-0.31** | -0.43; -0.19 | < .001 | -0.65; 0.03 | 27.33 (.038) | 0.16 | 43.33 | n.a. |

Abbreviations. $I^2$ = heterogeneity in outcomes in percent; $k$ = number of independent effect sizes included in the given analysis; n.a. = not applicable; PI = Prediction Interval. **Bold** font indicates statistical significance at $p < .050$ . Values are Hedges' $g$ standardized mean differences with 95% confidence intervals derived from the random effects meta-analysis.
<sup>a</sup>P-Values for moderator analyses concerning social media platform as a potential moderator. This moderator analysis was conducted only when at least two platforms/platform categories presented with sufficient data (i.e., $k \geq 4$ ), otherwise "n.a." is denoted for "not applicable".

**Table 4.** Meta-regression results concerning publication year and mean age as potential moderators of effects reported in experimental studies.

| Outcome | Moderator | *k* | *b* | 95% CI | *p* | Res. $\tau$ | Res. $I^2$ |
| --- | --- | --- | --- | --- | --- | --- | --- |
| **Upward social comparison vs. control (e.g., landscape pictures)** |  |  |  |  |  |  |  |
| **Positive affect** | Publication year | 10 | -0.037 | -0.087; 0.012 | .140 | 0.11 | 25.29 |
| **Negative affect** |  | 21 | -0.001 | -0.036; 0.033 | .934 | 0.15 | 39.25 |
| **Body dissatisfaction** |  | 38 | -0.017 | -0.045; 0.011 | .227 | 0.08 | 16.84 |
| **Positive affect** | Mean age | 10 | -0.008 | -0.037; 0.022 | .614 | 0.14 | 34.65 |
| **Negative affect** |  | 21 | -0.011 | -0.029; 0.008 | .247 | 0.13 | 33.41 |
| **Body dissatisfaction** |  | 38 | **-0.014** | -0.024; -0.005 | .004 | 0.01 | 0.33 |

Abbreviations. $I^2$ = heterogeneity in outcomes in percent; *k* = number of independent effect sizes included in the given random-effects meta-analysis.

In four studies comparing experimental groups with varying social comparison direction, upward vs. downward social comparison did not lead to differences in positive affect. Yet, the effects of these studies were very heterogenous. Similarly, the four studies assessing the effects of Thinspiration vs. Fitspiration on body dissatisfaction did not yield a significant difference (Hedges’ *g* = -0.08 (-0.30; 0.14), *P* = .480).

Given that many studies investigated the effect of positive body image manipulations on mental health, we further explored the influence of positive body image manipulations compared to control conditions (e.g., nature images) and upward social comparison conditions (e.g., social media profiles of influencers). We categorized conditions that depicted diverse body sizes, normalized physical imperfections, or promoted self-compassion toward one’s body as positive body image manipulations. Such manipulations had a statistically significant small effect on positive affect (Hedges’ *g* = 0.19 (0.07; 0.31), *P* = .002, *k* = 10). There was no heterogeneity (*τ* = 0). Positive body image manipulations had no statistically significant effect on negative affect or body dissatisfaction compared to control conditions. Compared to upward social comparison conditions, positive body image manipulations increased positive affect (Hedges’ *g* = 0.31 (0.19; 0.43), *P* < .001, *k* = 17), decreased negative affect (trim-and-fill adjusted Hedges’ *g* = -0.12 (-0.23; -0.00), *P* = .048, *k* = 24), and decreased body dissatisfaction (Hedges’ *g* = -0.32 (-0.46; - 0.19), *P* < .001, *k* = 35).

## Discussion

Social media usage and its implications for mental health are of broad societal concern^2–10^. This is the first comprehensive meta-analysis on the relationship between social comparisons occurring on social media and mental health. Findings from both observational and experimental research indicate that upward social comparisons are associated with worse mental health, whereas downward social comparisons were not. This suggests that social comparisons on social media are not inherently detrimental, rather their effects depend on comparison direction, comparison frequency, and perceived self-standard discrepancy. Upward comparisons indicate that one is doing worse than others, thereby drawing attention to discrepancies relative to others^18^. On social media, such discrepancies may be especially salient because users are frequently exposed to selectively presented positive information about others. Such exposure may not only increase opportunities for comparison, but also systematically bias both the standards against which the self is evaluated and the resulting discrepancies in an unfavorable direction. This interpretation is supported by the finding that a less favorable comparison-based self-evaluation was associated with greater psychopathology, body dissatisfaction, negative affect, and lower life satisfaction.

In observational studies, higher frequency of upward or non-directional social comparison as well as worse comparison-based self-evaluation were associated with adverse mental health. Experimental studies complemented these findings, showing that brief exposure to upward comparison content increased negative affect and body dissatisfaction and reduced positive affect. These effects emerged despite only brief experimental manipulations, suggesting that social comparison can rapidly influence affective and self-evaluative states. Overall, the convergence between observational and experimental findings strengthens the inference that upward social comparison adversely affects mental health. This is in line with a recent meta-analysis on the relationship between social comparison in general and mental health indicators in samples with mental disorders^18^. Notably, the effect sizes (e.g., magnitude of associations with mental health indicators) were very similar between the present work on online social comparisons and the prior work on general social comparison^18^. This may be particularly relevant for individuals with poor mental health, for whom the social media context could further amplify the adverse effects of upward social comparison.

Notably, the observational and experimental findings synthesized in the present meta-analysis serve different interpretive roles. Most observational studies were cross-sectional and therefore cannot establish temporal ordering or causality, whereas experimental provide evidence on short-term affective and cognitive responses to social comparisons. Accordingly, the experimental findings do not establish that naturally occurring social comparisons on social media cause persistent deterioration in mental health. Prospective findings from observational studies were consistent with an association between social comparison tendency and subsequent psychopathology, but were based on only four independent studies with widely varying follow-up periods. More longitudinal research is needed to determine whether short-term effects accumulate into more enduring changes in mental health. Additionally, future research should examine whether individuals experiencing current mental health problems are particularly vulnerable to frequent upward offline and online social comparisons.

The absence of significant associations for downward social comparison is also informative and in line with existing evidence on downward social comparison^285^. Upward vs. lateral vs. downward social comparison may serve different self-motives, with varying consequences for mental health^29^. Overall, downward comparisons are more likely to be processed as neutral, self-assuring or self-enhancing. Crucially, the available evidence was more limited for downward than upward comparisons. While these findings highlight the importance of considering social comparison direction when studying the psychological consequences of social media, more research on downward comparisons is needed. These findings may help explain heterogeneous associations between overall social media use and mental health, suggesting that how content is appraised may be more informative than exposure alone.

This perspective also has implications for how the social media environment itself is conceptualized. The current findings suggest that upward social comparison may reflect a cross-platform psychological process rather than a problem specific to some applications. Although social media platforms differ in their affordances and content, the opportunity of frequent self-evaluation relative to others who present idealized versions of themselves in remains a common feature. The one platform-specific difference observed in the present analyses—larger associations between comparison tendency and body dissatisfaction on Instagram than on Facebook, and a larger experimental effect of Fitspiration on Instagram than TikTok—suggests that platform affordances or content ecologies may nevertheless matter in specific domains. While these findings should be interpreted cautiously, they indicate that future research may benefit from platform-specific analysis (in addition to analysis across platforms).

The substantial heterogeneity observed across observational associations indicates that the role of upward social comparison may vary across users and contexts, while reflecting methodological differences in study design, measurement, and sampling. Although the pooled associations were robust across several sensitivity and subgroup analyses, considerable between-study heterogeneity remained. Individual differences in attitudes towards social comparison^286^, self-concept, mood and other psychological characteristics may influence both the likelihood of engaging in upward comparison and the engendered reactions. The higher upward comparison tendency in clinical vs. non-clinical groups further raises the possibility that social comparison processes are particularly salient among individuals experiencing psychopathology. However, because these case-control data were observational, they cannot establish whether heightened comparison contributes to psychopathology, results from it, or reflects reciprocal processes^18^.

The experimental finding that younger mean age was associated with larger effects of upward social comparison on body dissatisfaction suggests that younger users may be more sensitive to appearance-related social comparison, although the meta-regression was based on study-level rather than individual-level age information and should therefore not be interpreted as demonstrating that younger individuals are intrinsically more vulnerable. More generally, the included samples, on average, were relatively young and predominantly female, particularly in experimental research. This demographic concentration limits the generalizability of the findings. Future studies should explicitly examine developmental differences and recruit more diverse samples.

Several limitations should be considered. First, the measurement of observational data and its quality varied considerably. Only around one-third of observational studies assessed social comparison using a validated measure, and more than half did not explicitly distinguish the direction of comparison. Such measurement heterogeneity may contribute to the substantial between-study heterogeneity and makes it difficult to determine which specific components of social comparison are most important. In contrast, experimental studies generally used validated outcome measures and clearly defined comparison conditions, providing stronger evidence regarding short-term effects but addressing a narrower range of outcomes.

Second, most of the observational data were cross-sectional in nature. Therefore, associations between social comparison and mental health cannot be interpreted as causal or as reflecting a unidirectional temporal process. Individuals experiencing poorer mental health may be more likely to engage in upward comparison, while upward comparison may simultaneously contribute to poorer mental health. Reciprocal and dynamic processes may indeed be more plausible than a simple unidirectional pathway.

Third, evidence for several potentially important comparison processes was lacking. We identified no studies examining the selection of social-comparison standards, despite the abundance of idealized standards on social media. We likewise found no studies assessing how comparison outcomes are appraised in light of individuals’ motives and coping resources, even though such appraisals are likely to shape subsequent reactions, and ultimately, mental health^18^. Addressing neglected components of the comparison process should be a priority for future research.

To conclude, social comparisons on social media are not uniformly associated with poorer mental health. Adverse associations with mental health were concentrated on upward comparison, whereas downward comparison showed no significant associations. Experimental findings indicated that inducing upward social comparison can produce short-term deterioration in affective and body-image outcomes. Together, these findings identify upward social comparison as a robust and potentially modifiable process linking social media experiences to adverse mental health.

## Methods

### Protocol and Reporting

This systematic review and meta-analysis followed PRISMA guidelines^33^. The protocol was preregistered on PROSPERO (CRD42022330245; May 21, 2022, https://www.crd.york.ac.uk/PROSPERO/view/CRD42022330245). The systematic literature search, data extraction, and coding were performed independently by at least two raters, with any discrepancies and uncertainties resolved through discussions.

### Search strategy and eligibility criteria

We systematically searched Web of Science, PsycInfo, and MEDLINE from database inception to April 28, 2026 using all-field searches with the following terms: “social compar*” OR “upward comparison*” OR “downward comparison*” OR “lateral comparison*”. See the full search strategy in Appendix E in the online supplement. We further screened 35 reviews related to the present topic for further eligible studies (references in Appendix F).

A study was eligible if it met four inclusion criteria: 1) It assessed (observational study) or manipulated (experimental study) social comparison on social media, aligning with Wood’s^287^ definition of social comparison and Carr and Hayes’^288^ definition of social media; 2) It reported quantitative data linking social comparison on social media with at least one eligible mental health outcome (eligible outcomes below); 3) It included data of n ≥ 10 participants (per group, if multiple groups were compared); 4) It was published in English, German, or Dutch in a peer-reviewed scientific journal. Studies that assessed principally eligible data but reported them in insufficient detail for meta-analytic synthesis, and for which additional data could not be obtained from the authors via email, were excluded. Three kinds of research methods were eligible in line with Wood’s^287^ definition. One observational (i.e., labeled *narration studies*) design and two experimental designs (i.e., labeled *reaction studies* and *selection studies*). Observational or narration studies aim to examine naturally occurring social comparisons on social media, usually by means of self-reports assessing naturalistic social comparison behavior in retrospect. Experimental reaction studies test the causal effects of induced social comparisons on social media on mental health outcomes relative to a control condition in lab settings (randomized between-group design). Experimental selection studies examine which social comparison standard participants select when explicitly instructed to choose among available alternatives. Yet, our search identified no published selection studies of social comparison on social media.

### Social comparison as a process and process components

Our systematic review and meta-analysis was guided by the general comparative-processing model (gCOMP)^29^, which conceptualizes the social-comparison process as comprising four components: a) seeking or encountering social information, b) the basic comparison process of evaluating similarities and discrepancies between the target and the comparison standard, c) the valuation of the comparison outcome in relation to the individual’s motives and coping resources, and d) the resulting emotional, cognitive, and behavioral responses. A central feature of the model is its emphasis on the motivational and coping-related appraisal of comparison outcomes. This extension draws on appraisal theories, which propose that individuals evaluate the relevance of an outcome to their motives as well as its controllability and their capacity to cope with its implications^289^.

Accordingly, we sought to include studies addressing any of the four components of the comparison process: a) seeking or encountering social information, b) evaluating the discrepancy between the target and the comparison standard, c) appraising the comparison outcome, and d) responding to the comparison. Importantly, studies could operationalize these components in different ways. For example, the first component could be assessed in terms of the frequency of comparisons in everyday life, a general tendency to engage in social comparison, or preferences for selecting particular comparison standards. The second component could be assessed directly through perceived discrepancies between the target and the standard or more broadly through measures of relative standing. The third component could be assessed by examining the motivational meaning or personal significance attributed to the comparison outcome, including its relevance to personal needs or goals and perceived coping possibilities. Finally, responses to comparison could be assessed either in terms of individual’s typical responses to social comparisons or as responses elicited by a specific comparison, for example following experimental exposure to particular comparison standards.

Regarding study design, we followed Wood’s^287^ taxonomy of social-comparison methods, distinguishing selection, reaction, and narration studies. Selection studies used (quasi-)experimental designs to examine which comparison standards (e.g., upward vs. downward) participants selected. Reaction studies used (quasi-)experimental designs to assess responses to experimentally induced social comparisons. Narration studies assessed naturally occurring social comparisons using forced-choice or open-ended self-report or interview methods. In the context of narrative studies, we identified three distinct aspects of social comparison: Social comparison tendency, social comparison-based self-evaluation, and social rank.

Social comparison tendency refers to the frequency at which individuals engage in social comparison. An example of a measurement tool is the comparison-based social media use subscale of the Social Media Use Scale (SMUS)^290^. This scale assesses participants’ engagement in various social media activities during the past week (e.g., “I compared my body or appearance to others”). Responses are rated on a 9-point scale ranging from 1 (never) to 9 (hourly or more). Notably, this scale evaluates the tendency without restricting it to a specific direction (i.e., non-directional).

Social comparison-based self-evaluation refers to the evaluation of oneself relative to other on a particular dimension, such as physical appearance. To illustrate, the brief three-item scale developed by Lee^150^ includes items like “When I read news feeds (or see others’ photos), I often think that others are having a better life than me.” Participants rate their agreement with each item on a 5-point Likert scale, ranging from 1 (strongly disagree) to 5 (strongly agree). In this example, social comparison-based self-evaluation relates to life in general and is assessed in the upward direction.

In contrast, social rank refers to an individuals’ perceived position relative to others in general, encompassing multiple social comparison dimensions rather than a single specific dimension. The Social Comparison Scale by Allan and Gilbert^291^, adapted to the social media context by Feinstein et al.^123^, serves as an illustrative measure.

Participants rate themselves relative to others on Facebook on 11 bipolar dimensions (e.g., inferior–superior, incompetent–competent, unattractive–attractive), on a 10-point scale. The sum score across these dimensions yields an overall measure of perceived social rank, with higher scores indicating a higher perceived social rank. In our meta-analysis, social comparison-based self-evaluation and social ranking are first analysed jointly and then followed by separate analyses.

### Data Extraction

Full-texts and supplementary materials were screened for eligible data. Missing data were calculated whenever open data and codebook were provided. For each eligible study, we recorded bibliographic details (e.g., authors, publication year, number of studies reported in the given publication), sample characteristics (e.g., number of groups, sample size[s], mean age, proportion females), study design, other relevant design specifications (e.g., for observational studies: cross-sectional vs. longitudinal assessment), social comparison features (e.g., for observational studies: component of the social comparison process assessed, for observational and experimental studies: direction of social comparison, social comparison dimension), social media platform, mental health outcome, and quality criteria (details below). We included diverse mental health outcomes. In terms of unfavorable mental health outcomes, we included any psychopathology (i.e., any classified mental disorder), body dissatisfaction, and negative affect. In terms of favorable mental health, we included satisfaction with life, positive affect, and psychological well-being. For the experimental studies, we additionally included behavioral outcomes (e.g. eye fixation duration on body parts of the comparison standard). However, the number of studies with behavioral outcomes was too small for synthesis (*k* < 4).

### Quality Assessment

Study quality was rated using McCarthy and Morina’s^17^ criteria, adapted for the context of social media as the original items related to social comparison in general (see Appendix G for the items and their scoring). Higher scores indicated greater methodological rigor and vice versa.

### Data synthesis and statistical analysis

Analyses were conducted in R 4.5.1^292^ using *metafor* 4.8.0^293^. Meta-analyses were performed whenever *k* ≥ 4 independent data points were available^294^. Random-effects models were used, given expected variability in effects. Heterogeneity was quantified using *I²*^295^ and tested with Cochran’s *Q*^296^. *I²* values of 25%, 50%, and 75% were interpreted as low, moderate, and high heterogeneity, respectively^297^. Claude Design was used to visualize meta-analytic results depicted in Figure 2. We only performed independent meta-analyses. That is, only one effect per study was used for any given meta-analysis.

For observational studies, Pearson correlations were transformed using Fisher’s *Z*, pooled meta-analytically, and back-transformed. Correlational effects were interpreted as small (.10), medium (.30), and large (.50) correlations, respectively^298^. For experimental studies, standardized mean differences (Hedges’ *g*) were computed from means, standard deviations, and sample sizes of experimental vs. control condition^299^, and interpreted as small (0.20), medium (0.50), and large (0.80) effects, respectively^298^. We planned to analyze selection preferences of participants in % in selection studies but there were no selection studies in the current literature, precluding quantitative synthesis. Publication year and sample mean age were examined as potential moderators of effects via meta-regressions, performed only when the evidence base was sufficient with *k* ≥ 10 independent data points^299^. We performed sub-group analyses for specific psychopathology outcomes (e.g., depression only, eating disorder only), for social comparison directions (e.g. upward social comparison tendency only, non-directional social comparison tendency only), for each platform (e.g., Instagram only, Facebook only), for appearance-based social comparison only, for females only, for young people only (sample mean age ≤ 21), or for high-quality studies only (quality sum score ≥ 7/8) whenever *k* ≥ 4 data points had accumulated for the given analysis^294^. To evaluate the robustness of the meta-analytic findings, several outlier and influence diagnostics were calculated^293^. Externally studentized residuals and influence diagnostics were computed using the influence function from the *metafor* package, which yields multiple indices such as Cook’s distance and DFBETAS. Studies identified by these diagnostics were regarded as potentially influential. Lastly, leave-one-out analyses were performed to assess whether omitting any individual study led to substantial changes in the overall effect estimate. Whenever one or more outliers were detected, the given analysis was repeated excluding outliers. Potential effect of publication bias on pooled outcomes was examined via Egger’s test^300^ and corrected via the trim-and-fill method (Duval & Tweedie, 2000). In line with recommendations, we only performed Egger’s test when the evidence base was sufficient, i.e., analyses with *k* ≥ 10 independent data points^301^. Significant asymmetry detected by Egger’s test was corrected via the trim-and-fill adjustment by adding fictious studies until symmetry was reached^302^. Corrected results were reported alongside the original result.

## Data availability

The data that support the findings are available in the paper and are publicly available on Zenodo: will be made available once published

## Code availability

The codes generated for data analyses are publicly available on Zenodo: will be made available once published

## Acknowledgments

We thank Dr. Janna Nelson, M.Sc. Jana Payer, M.Sc. Anna Steinweg, M.Sc. Marielle Placzek, M.Sc. Maryse Müller, M.Sc. Valentin Rudloff, B.Sc. Elisabeth Falke, and B.Sc. Carolin Hans for their contributions to parts of the systematic literature search and data extraction.

## Author contributions

T.H.H.: conceptualization, methodology, investigation, supervision, visualization, formal analysis, writing—original draft, writing—review and editing. L.H.: investigation, methodology, formal analysis, writing—review and editing. P.S.: methodology, writing—review and editing, H.H.: methodology, formal analysis, writing—review and editing. J.M.: visualization, formal analysis, writing—review and editing. N.M.: conceptualization, methodology, supervision, writing—review and editing. All authors (T.H.H., L.H., P.S., H.H., J.M., and N.M.) reviewed and approved the paper.

