## Supplementary Appendix for "Social comparisons on social media and mental health: A systematic review and meta-analysis"

**Appendix A – Characteristics of observational studies**

**Appendix B – Characteristics of experimental studies**

**Appendix C – Social media platform-specific meta-analytic results for observational studies assessing the relationship between social comparison on social media and mental health outcomes**

**Appendix D – Social media platform-specific meta-analytic results for experimental studies assessing the effects of social comparison on social media on mental health outcomes**

**Appendix E – Search strategy**

**Appendix F – References of screened reviews as part of the literature search**

**Appendix G – Study quality assessment (items and scoring)**

**Appendix A – Characteristics of observational studies**

| Reference | Country | Sample  (n, age range, mean age, percentage female) | Social comparison component(s) assessed | Social comparison direction | Social comparison dimension | Social media platform | Effect-size | Outcome Variable (measure) | Study quality sum score (maximum of 8, unless indicated otherwise) |
| --- | --- | --- | --- | --- | --- | --- | --- | --- | --- |
| Ang, 2026 | Singapore | Sample recruited online via social media (*n* = 228, 18-40, *M* = 23.46, *SD* = 5.36, 82%) | Tendency | Non-directional | Ability | Unspecified | r | Happiness (Subjective Happiness Scale, Lyubomirsky and Lepper, 1999) | 5 |
| Aubry et al., 2024; Study 1 | Switzerland | Sample recruited online via Instagram (*n* = 463, 18-65, *M* = 28.94, *SD* = 8.73, 76%) | Tendency  Affective response | Upward  Upward | Unspecified | Instagram | r | Depression (PHQ-8) | 6 |
| Aubry et al., 2024; Study 2 | Switzerland | Sample recruited online via Instagram (*n* = 130, 18-36, *M* = 20.79, *SD* = 3.34, 15%) | Self-evaluation | Upward, downward | Unspecified | Instagram | r | Depression (PHQ-8) | 6 |
| Aubry et al., 2024; Study 3 | Switzerland | Students (*n* = 168, 18-52, *M* = 21.38, *SD* = 4.36, 80%) | Self-evaluation    Motives | Upward, downward  Upward | Unspecified | Instagram | r | Depression (PHQ-8) | 6 |
| Bachner-Melman et al., 2018 | Israel | Clinical vs. non-clinical (*n* = 122, 12-30, *M* = 21.60, *SD* = 4.40, 100%) | Tendency | Upward | Appearance | Unspecified | SMD | / | 3 (out of 4)^a^ |
| Berkout & Flynn, 2024 | US | Adults who self-identified as diagnosed with a mental health condition (*n* = 295, 18-64, *M* = 33.60, *SD* = 10.90, 60%) | Tendency | Upward | Unspecified | Facebook | r | Psychopathology (DASS-21)  Satisfaction with Life Scale (SWLS) | 8 |
| Bhat et al., 2023 | India | Students (*n* = 629, 19-25, *M* = 20.00, *SD* = 3.10, 44%) | Tendency | Mixed (upward & downward) | Unspecified | Multiple | r | Depression (PHQ-9) | 5 |
| Boer et al., 2021 | Netherlands | Students (*n* = 1642, 10-16, *M* = 13,1, *SD* = 0,80, 43%) | Tendency | Upward | Unspecified | Unspecified | r | Depression (Depressive Mood List, Kandel & Davies, 1982)  Satisfaction with Life (SLSS) | 6 |
| Boer et al., 2022 | Netherlands | Students (*n* = 1419, *M* = 12.51, *SD* = 0.60, 46%) | Tendency | Upward | Unspecified | Unspecified | r | Satisfaction with Life (SLSS) | 6 |
| Bonfanti et al., 2022 | Italy | Students (*n* = 491, 18-63, *M* = 24.55, *SD* = 7.25, 80%) | Tendency | Non-directional | Unspecified | Facebook | r | Depression (DASS-21, depression subscale)  Satisfaction with life (SWLS) | 5 |
| Brown et al., 2025 | Australia | Sample recruited online via social media and from a university student subject pool (*n* = 548, 18-84, *M* = 33.16, *SD* = 17.37, 73%) | Tendency | Non-directional | Appearance | Unspecified | r | Depression (PHQ-8)  Generalized anxiety disorder (GAD-7) | 6 |
| Casale et al., 2024 | Italy | Sample recruited online (*n* = 323, *M* = 29.97, *SD* = 12.05, 69%) | Tendency | Non-directional | Appearance | Instagram | r | Nondisplay of bodily imperfections (PCPS-BI) | 6 |
| Chae, 2018 | South Korea | Sample recruited via e-mail (*n* = 1064, 20-39, *M* = 29.3, *SD* = 5.32, 100%) | Tendency | Non-directional | Unspecified | Unspecified | r | Happiness (four items adapted from Koivumaa-Honkanen et al., 2001)  Satisfaction with life (1 item) | 0 |
| Chan & Lam, 2026 | China | Sample recruited online via social media and via e-mail systems of local universities (*n* = 330, 18-25, *M* = 21.24, *SD* = 1.95, 75%) | Tendency | Non-directional | Ability | Unspecified | r | Social media addiction (BSMAS) | 6 |
| Chang et al., 2019 | Singapore | Students (*n* = 303, 12-16, 100%) | Tendency  Self-evaluation | Non-directional  bipolar | Appearance | Instagram | r | Bipolar body esteem, reverse coded (modified Body esteem scale, Mendelson et al., 1996) | 7 |
| Chen et al., 2023 | China | Students (*n* = 1792, *M* = 23.27, *SD* = 5.44, 48%) | Self-evaluation | Upward | Unspecified | Unspecified | r | Depression (CES-D) | 8 |
| Choi, 2022 | South Korea | Students (*n* = 360, 53%) | Self-evaluation | Upward | Unspecified | Unspecified | r | Satisfaction with life (SWLS) | 6 |
| Christensen-Duerden & Coyne, 2025 | US | Sample recruited online via Qualtrics (*n* = 1231, 10-17, *M* = 14.49, *SD* = 1.99, 55%) | Tendency | Non-directional | Unspecified | Unspecified | r | Depression (PHQ-8) | 6 |
| Coyne et al., 2017 | US | Mothers recruited via e-mails and fliers (*n* = 703, *M* = 30.38, *SD* = 5.15, 100%) | Tendency | Non-directional | Unspecified | Unspecified | r | Depression (CES-D)  Satisfaction with life (SWLS) | 4 |
| Coyne et al., 2023 | US | Sample recruited online via Qualtrics (*n* = 1159, 10-17, *M* = 14.5, *SD* = 2.00, 55%) | Tendency | Non-directional | Unspecified | Unspecified | r | Depression (PHQ-8)  Conduct problems (SDQ) | 4 |
| De Vries & Kühne, 2015 | Netherlands | Sample recruited through online social networks (*n* = 231, 18-25, *M* = 22.3, *SD* = 2.20, 69%) | Self-evaluation | Upward | Unspecified | Facebook | r | Satisfaction with life (SWLS) | 6 |
| Di Gesto et al., 2022 | Italy | Students (*n* = 305, 19-32, *M* = 23, *SD* = 2.92, 100%) | Self-evaluation  Tendency | Upward  Non-directional | Physical appearance | Instagram | r | Body dissatisfaction (BSQ-14) | 6 |
| Díaz-Moreno et al., 2026 | Spain | Sample recruited from High School and other participants residing in Barcelona (*n* = 256, 16-35, *M* = 19.25, *SD* = 3.23, 66%) | Self-evaluation | Upward | Unspecified | Unspecified | r | Social media addiction (BSMAS)  Suicide risk (SBQ-R) | 8 |
| Dibb & Foster, 2021 | United Kingdom | Facebook users (*n* = 214, 18-72, *M* = 35.65, *SD* = 15.10, 81%) | Self-evaluation  Tendency | Upward, downward  Non-directional | Unspecified | Facebook | r | Depression (CES-D) | 7 |
| Dibb, 2019 | United Kingdom | Facebook users (*n* = 165, 18-70, *M* = 31.4, *SD* = 13.95, 67%) | Affective response | Upward | Unspecified | Facebook | r | Depression (HADS-D)  Anxiety (HADS-A)  Satisfaction with life (SWLS) | 7 |
| Dondzilo et al., 2023 | Australia | Students (*n* = 230, 17-30, *M* = 19.07, *SD* = 1.87, 78%) | Tendency | Upward | Unspecified | Unspecified | r | Eating disorder symptoms (EDE-Q) | 7 |
| Eid et al., 2023 | Lebanon | Adolescents currently residing in Lebanon (*n* = 379, 13-17, *M* = 16.07, *SD* = 1.19, 65%) | Self-evaluation | Upward | Unspecified | Unspecified | r | Addictive consequences (PUS) | 7 |
| Etherson et al., 2022 | United Kingdom | Students (*n* = 135, *M* = 14.70, *SD* = 0.46, 100%) | Self-evaluation | Bipolar | Appearance | Unspecified | r | Depression (3-item scale) | 2 |
| Faelens et al., 2019; Study 1 | Belgium | Sample recruited online via Prolific (*n* = 207, 18-35) | Tendency | Non-directional | Unspecified | Facebook | r | Depression (DASS-21) | 5 |
| Faelens et al., 2019; Study 2 | Belgium | Sample recruited online via Prolific (*n* = 468, 18-35) | Tendency | Non-directional | Unspecified | Facebook | r | Depression (DASS-21) | 5 |
| Faelens et al., 2021a | Belgium | Facebook users (*n* = 459, 18-35, *M* = 26.2, *SD* = 4.53, 51%) | Tendency | Non-directional | Unspecified | Facebook | r | Depression (DASS-21) | 5 |
| Faelens et al., 2021b - Facebook sample | Belgium | Sample recruited online via social media (*n* = 98, 18-35, *M* = 22.39, *SD* = 3.08, 76%) | Tendency | Non-directional | Unspecified | Facebook | r | Depression (DASS-21) | 5 |
| Faelens et al., 2021b - Instagram sample | Belgium | Sample recruited online via social media (*n* = 98, 18-35, *M* = 22.39, *SD* = 3.08, 76%) | Tendency | Non-directional | Unspecified | Instagram | r | Depression (DASS-21) | 5 |
| Faranda & Roberts, 2019 | Australia | Population-based (*n* = 181, 18-25, *M* = 21.9, *SD* = 2.14, 72%) | Self-evaluation  Tendency | Bipolar  Non-directional | Unspecified | Facebook | r | Depression (CES-D) | 5 |
| Fardouly & Vartanian, 2015 | Australia | Students (*n* = 227, *M* = 19.13, *SD* = 2.21, 100%) | Self-evaluation  Tendency | Bipolar  Non-directional | Appearance | Facebook |  | Body dissatisfaction (EDI-BD)  Drive for thinness (EDI-DT) | 6 |
| Fardouly et al., 2018a | Australia | Students (*n* = 284, 10-12, *M* = 11.2, *SD* = 0.56, 53%) | Tendency | Non-directional | Appearance | Unspecified | r | Depression (SMFQ-C)  Satisfaction with life (SLSS) | 4 |
| Fardouly et al., 2018b | Australia | Students (*n* = 276, 18-25, *M* = 22.83, *SD* = 3.57, 100%) | Tendency | Non-directional | Appearance | Instagram | r | Body dissatisfaction (EDI-BD)  Drive for thinness (EDI-DT) | 6 |
| Fardouly et al., 2020 | Australia | Students (*n* = 528, 10-12, *M* = 11.19, *SD* = 0.55, 49%) | Self-evaluation  Tendency | Bipolar  Non-directional | Appearance | Unspecified | r | Depression (SMFQ-C)  Eating disorder symptoms (Children’s Eating Attitude Test, Maloney et al., 1988)  Social anxiety disorder symptoms (Spence Children’s Anxiety Scale, Spence, 1998) | 6 |
| Feijoo & Vizcaino-Verdu, 2025 | Spain | Sample recruited through a panel service (*n* = 1082, 12-17, *M* = 14.5, 50%) | Tendency | Upward | Appearance | Unspecified | r | Bipolar body satisfaction, reverse coded (SATIS scale, Carlson-Jones & Crawford, 2005) | 8 |
| Feinstein et al., 2013 | US | Students (*n* = 268, *M* = 19.66, *SD* = 2.29, 62%) | Self-evaluation | Bipolar | Unspecified | Facebook | r | Depression (CES-D) | 7 |
| Ferdousi et al., 2023 | Australia | Students (*n* = 284, 17-19, *M* = 18.29, *SD* = 0.60, 100%) | Tendency | Upward | Physical appearance | Instagram | r | Body dissatisfaction (BISS) | 7 |
| Fiovaranti et al., 2024 | Italy | Clinical + students (*n* = 386, *M* = 26.04, *SD* = 6.73, 100%) | Tendency | Non-directional | Physical appearance | Instagram | r & SMD | Body uneasiness (BUT-A GSI) | 5 |
| Foster et al., 2022 | US | Students (*n* = 527, *M* = 19, *SD* = 1.88, 100%) | Tendency | Non-directional | Physical appearance | Unspecified | r | Drive for thinness (EDI-DT) | 4 |
| Fox & Vendemia, 2016 | US | Sample recruited online via Qualtrics (*n* = 1686, 18-40, *M* = 29.31, *SD* = 6.34, 54%) | Self-evaluation | Upward, downward | Appearance | Multiple | r | Body image, reverse coded (7-item scale) | 2 |
| Frison & Eggermont, 2016 | Belgium | Students (*n* = 1621, 12-19, *M* = 14.76, *SD* = 1.41, 48%) | Self-evaluation | Upward | Unspecified | Facebook | r | Satisfaction with life (SWLS) | 6 |
| Gao et al., 2023 | China | Students (*n* = 616, 16-27, *M* = 19.59, *SD* = 1.79, 57%) | Tendency | Upward | Unspecified | Unspecified | r | Online compulsive buying (Online compulsive shopping scale, Zeng et al., 2014) | 8 |
| Gerson et al., 2016 | United Kingdom | Sample recruited onlie via MTurk and social media (*n* = 337, 18-70, *M* = 36.5, *SD* = 11.3, 60%) | Self-evaluation | Bipolar | Unspecified | Facebook | r | Satisfaction with life (SWLS) | 7 |
| Glatz et al., 2026 | Sweden | Parents (*n* = 278, *M* = 32.65, *SD* = 5.17) | Tendency  Affective response | Non-directional  Upward, downward | Unspecified | Unspecified | r | Depression (CES-D) | 7 |
| Hai & Yang, 2022 | China | Students (*n* = 320, 17-39, *M* = 21.6, *SD* = 3.20, 100%) | Tendency | Non-directional | Appearance | Unspecified | r | Anxiety (DASS-21, anxiety subscale) | 6 |
| Hanna et al., 2017 - females | US | Students (*n* = 690, *M* = 19.11, 100%) | Tendency | Mixed (upward & downward) | Unspecified | Unspecified | r | Depression (BSI)  Anxiety (BSI) | 5 |
| Hanna et al., 2017 - males | US | Students (*n* = 414, *M* = 19.43, 0%) | Tendency | Mixed (upward & downward) | Unspecified | Unspecified | r | Depression (BSI)  Anxiety (BSI) | 5 |
| Hartlaub & Hill, 2025 | US | Sample recruited online via Prolific (*n* = 199, 18-40, *M* = 29.23, *SD* = 5.67, 55%) | Tendency | Non-directional | Physical appearance | Unspecified | r | Bipolar body satisfaction, reverse coded (BESAA) | 5 |
| Hendrickse et al., 2017 | US | Students (*n* = 185, *M* = 21.04, *SD* = 3.55, 100%) | Tendency | Non-directional | Physical appearance | Instagram | r | Drive for thinness (EDI-DT)  Body dissatisfaction (EDI-BD) | 5 |
| Her & Timmermans, 2021 | US | Tinder users recruited online via MTurk (*n* = 296, 18-29, *M* = 26.3, *SD* = 2.90, 39%) | Self-evaluation | Upward | Unspecified | Tinder | r | Anxiety (1 item adapted from Dhir et al., 2018) | 6 |
| Jabeen et al., 2023a; Study B | India | Students (*n* = 479, *M* = 24, 39%) | Affective response | Upward | Unspecified | Facebook | r | Social media addiction (items validated in the study, Dhir et al. 2021) | 5 |
| Jabeen et al., 2023a; Study C | India | Students (*n* = 618, *M* = 23, 35%) | Affective response | Upward | Unspecified | Facebook | r | Social media addiction (items validated in the study, Dhir et al. 2021) | 5 |
| Jabeen et al., 2023b | US | Sample recruited online via Prolific (*n* = 305, 25-55, 43%) | Tendency | Non-directional | Unspecified | Unspecified | r | Social anxiety disorder symptoms (items validated in the study, Dhir et al. 2018) | 6 |
| Jarman et al., 2021 | Australia | Students (*n* = 1432, 11-17, *M* = 13.45, *SD* = 1.15, 41%) | Tendency  Self-evaluation | Upward, Non-directional  Upward | Appearance | Unspecified | r | Satisfaction with life (SWLS) | 7 |
| Jarman et al., 2024 | Australia | Students (*n* = 915, 11-16, *M* = 14.27, *SD* = 1.08, 44%) | Tendency | Non-directional | Unspecified | Unspecified | r | Body satisfaction, reverse coded (modified version of the BSS) | 1 |
| Jian & An, 2026 | China | Students (*n* = 1140, *M* = 20.62, *SD* = 1.87, 63%) | Tendency | Upward | Unspecified | Unspecified | r | Depression (CES-D) | 8 |
| Jung et al., 2022 | US | Sample recruited online via social media and university students (*n* = 579, 17-26, *M* = 20.21, *SD* = 1.38, 100%) | Tendency | Non-directional | Physical appearance | Unspecified | r | Body esteem, reverse coded (BESAA) | 6 |
| Kaminger et al., 2023 | Austria and Germany | Instagram users (*n* = 268, 18-39, *M* = 22.84, *SD* = 3.85, 63%) | Tendency | Non-directional | Unspecified | Instagram | r | Negative affect (PANAS)  Positive affect (PANAS) | 5 |
| Kim, 2018 | South Korea | Sample recruited through online panels (*n* = 305, 18–29, *M* = 23.44, *SD* = 2.64, 100%) | Tendency | Non-directional | Appearance | Facebook | r | Drive for thinness (EDI-DT) | 5 |
| Kinsella & Chin, 2024 | US | Sample recruited online via Prime Panels (*n* = 830, 18-30, *M* = 24, *SD* = 3.90, 63%) | Self-evaluation | Upward | Life | Multiple | r | Insomnia (ISI) | 7 |
| Kumar & Kumar, 2024 | India | Sample recruited online via social media and e-mails (*n* = 127) | Tendency | Non-directional | Unspecified | Unspecified | r | Compulsive buying (Sneath et al., 2009) | 6 |
| Lajunen & Olsen Haug, 2023 | Norway | Sample recruited online via Facebook and university campuses (*n* = 315, *M* = 24.5, *SD* = 9.07, 73%) | Tendency | Non-directional | Unspecified | Instagram | r | Satisfaction with life (SWLS) | 5 |
| Lavell et al., 2023 | Australia | Clinical vs. non-clinical (*n* = 51, 12-17, *M* = 14.05, *SD* = 1.57, 71%) | Tendency | Upward | Appearance | Unspecified | SMD | / | 4 (out of 4) |
| Le Blanc-Brillon et al., 2025; Study 2 – Facebook sample | US and Canada | Sample recruited online via MTurk (*n* = 207, 18-35, *M* = 28.54, *SD* = 3.93, 59%) | Self-evaluation  Tendency | Upward, downward  Upward, downward | Unspecified | Facebook | r | Depression (BDI) | 8 |
| Le Blanc-Brillon et al., 2025; Study 2 – Instagram sample | US and Canada | Sample recruited online via MTurk (*n* = 206, 18-35, *M* = 28.53, *SD* = 3.89, 57%) | Self-evaluation  Tendency | Upward, downward  Upward, downward | Unspecified | Instagram | r | Depression (BDI) | 8 |
| Lee & Lee, 2021 | South Korea | Sample recruited online through a survey company (*n* = 385, 20-39, *M* = 29.83, *SD* = 4.93, 100%) | Tendency | Non-directional | Physical appearance | Unspecified | r | Body satisfaction, reverse coded (MBSRQ-AE) | 6 |
| Lee, 2014 | US | Students (*n* = 191, 18-23, *M* = 19.9, 38%) | Self-evaluation  Tendency | Upward  Non-directional | Unspecified | Facebook | r | Depression (CES-D)  Anxiety (Scale by Costello & Comrey, 1967) | 6 |
| Lee, J. K. 2022 | South Korea | University students and general public (*n* = 236, *M* = 25.8, 44%) | Tendency | Upward | Unspecified | Unspecified | r | Psychological well-being (12 items adapted from Kim et al., 2001) | 3 |
| Lee, M. 2022 | South Korea | Sample recruited online through an online research firm (*n* = 321, 20-29, *M* = 25.29, *SD* = 2.60, 100%) | Tendency | Non-directional | Appearance | Instagram | r | Drive for thinness (EDI-DT) | 5 |
| Li & Liu, 2024 | China | Students (*n* = 1078, 17-24, *M* = 19.74, *SD* = 1.26, 54%) | Tendency | Upward | Unspecified | Unspecified | r | Psychological well-being (OHQ-S) | 7 |
| Li et al., 2024 | China | Students (*n* = 329, *M* = 21.37, *SD* = 2.54, 16%) | Tendency | Upward | Unspecified | Unspecified | r | Anxiety (anxiety scale compiled by Zung, 1971)  Alcohol dependence (ADS) | 8 |
| Li, 2019 | China | Students (*n* = 934, 16-21, *M* = 17.86, *SD* = 1.27, 59%) | Tendency | Upward | Unspecified | Unspecified | r | Depression (CES-D) | 7 |
| Liu et al., 2017 | China | Students (*n* = 1205, 17-24, *M* = 19.86, *SD* = 1.27, 51%) | Tendency | Upward | Unspecified | Unspecified | r | Depression (CES-D) | 7 |
| Liu et al., 2019 | China | Students (*n* = 430, 17-24, *M* = 19.27, *SD* = 1.10, 58%) | Tendency | Upward | Unspecified | Unspecified | r | Negative affect (PANAS) | 7 |
| Liu et al., 2024 | China | Social media users (*n* = 338, 18-34, 100%) | Tendency | Non-directional | Unspecified | Unspecified | r | Anxiety (GAD-7) | 5 |
| Lup et al., 2015 | US | Instagram users (*n* = 117, 18-29, *M* = 24.81, *SD* = 2.51, 84%) | Self-evaluation | Bipolar | Unspecified | Instagram | r | Depression (CES-D) | 7 |
| Mancin et al., 2024 | Italy | Students (*n* = 149, 19-27, *M* = 21.58, *SD* = 1.44, 100%) | Self-evaluation  Tendency | Upward  Non-directional | Physical appearance | Instagram | r | Body dysmorphic disorder symptoms (QDC) | 5 |
| Masciantonio et al., 2021 | Belgium | Sample recruited through academic mailing lists from social science (*n* = 793, 18-77, *M* = 33.75, *SD* = 14.70, 77%) | Self-evaluation | Upward | Unspecified | Unspecified | r | Negative affect (7-item scale)  Positive affect (6-item scale)  Satisfaction with life (SWLS) | 7 |
| Masood et al., 2020 | China | Students (*n* = 505, 50%) | Tendency | Non-directional | Unspecified | Unspecified | r | Negative affect (not reported)  Positive affect (not reported) | 1 |
| Meier & Schäfer, 2018 | Germany | Instagram users (*n* = 385, 18-52, *M* = 22.64, *SD* = 4.00, 82%) | Tendency | Non-directional | Unspecified | Instagram | r | Negative affect (PANAS short version)  Positive affect (PANAS short version) | 5 |
| Modica, 2019 | US | Sample recruited online via MTurk (*n* = 232, 20-72, *M* = 35.79, *SD* = 11.08, 100%) | Tendency | Non-directional | Physical appearance | Facebook | r | Body esteem, reverse coded (BESAA) | 3 |
| Moroney et al., 2023 | US | Students (*n* = 2616, 13-19, *M* = 16.10, *SD* = 1.21, 47%) | Tendency | Upward | Unspecified | Unspecified | r | Externalizing disorders (YSR)  Internalizing disorders (YSR) | 6 |
| Nagl et al., 2021 | Germany | Postpartum women (*n* = 252, 18-45, *M* = 30.65, *SD* = 4.43, 100%) | Tendency | Non-directional | Physical appearance | Multiple | r | Eating disorder symptoms (EDE-Q)  Body dissatisfaction (BSQ) | 4 |
| Niu et al., 2018 | China | Students (*n* = 764, 12-19, *M* = 14.23, *SD* = 1.75, 47%) | Self-evaluation | Upward | Unspecified | Qzone | r | Depression (CES-D) | 7 |
| Niu et al., 2025 | China | Sample recruited via online referrals and posters (*n* = 970, 16-30, *M* = 19.42, *SD* = 2.01, 84%) | Self-evaluation | Upward | Unspecified | Unspecified | r | Negative affect (PANAS)  Positive affect (PANAS)  Satisfaction with life (SWLS)  Subjective well-being (calculated by adding the standardized scores of life satisfaction and positive affect and then subtracting the standardized score of negative affect) |  |
| Ouvrein et al., 2023 | Belgium | Unspecified sample (*n* = 385, 18-83, *M* = 42.24, *SD* = 14.46, 60%) | Tendency | Upward | Unspecified | Unspecified | r | Psychopathology (K6 Psychological Distress Scale, Kessler et al., 2010) | 6 |
| Pacewicz & Mellano, 2024 | US | Students (*n* = 162, 18-23, *M* = 19.3, *SD* = 2.10, 40%) | Self-evaluation | Bipolar | Unspecified | Unspecified | r | Burnout (ABQ) | 8 |
| Padoa et al., 2018 | Australia | Mothers (*n* = 201, 22-53, *M* = 37.38, *SD* = 5,04, 100%) | Tendency | Non-directional | Parenting | Multiple | r | Depression (DASS-21)  Anxiety (DASS-21) | 5 |
| Pang et al., 2023 | China | Students (*n* = 335, 18-33, *M* = 27, *SD* = 0.87, 50%) | Self-evaluation | Upward | Unspecified | Unspecified | r | Satisfaction with life (5-item scale by Guo et al., 2014) | 7 |
| Pang, 2021 | China | Students (*n* = 318, 18-29, *M* = 22.5, *SD* = 0.92, 50%) | Self-evaluation | Upward | Unspecified | Wechat | r | Depression (PROMIS, 4 adapted items) | 6 |
| Park & Baek, 2018 | South Korea | Sample recruited from the Embrain online panel (*n* = 331, *M* = 32.05, *SD* = 12.74, 53%) | Tendency | Non-directional | Ability | Facebook | r | Satisfaction with life (SWLS)  Negative affect (Smith’s classification, 2000) | 5 |
| Pedalino & Camerini, 2022 | Italy | Sample recruited online via online survey and social media (*n* = 291, *M* = 19.8, *SD* = 4.57, 100%) | Self-evaluation | Bipolar | Unspecified | Instagram | r | Body dissatisfaction (BDS) | 6 |
| Piccoli et al., 2022; Study 1 | Italy | Instagram users (*n* = 115, 19-33, *M* = 25.42, *SD* = 2.56, 100%) | Tendency | Non-directional | Appearance | Instagram | r | Drive for thinness (EDI-DT) | 7 |
| Piccoli et al., 2022; Study 2 | Italy | Instagram users (*n* = 120, 18-29, *M* = 21.38, *SD* = 2.30, 100%) | Tendency | Non-directional | Appearance | Instagram | r | Drive for thinness (EDI-DT) | 7 |
| Prieler et al., 2021 – Austria sample | Austria | Students (*n* = 199, 11-15, *M* = 13.46, *SD* = 1.10, 100%) | Tendency | Non-directional | Appearance | Facebook | r | Body esteem, reverse coded (BESAA) | 6 |
| Prieler et al., 2021 – Belgium sample | Belgium | Students (*n* = 292, 11-15, *M* = 13.46, *SD* = 1.10, 100%) | Tendency | Non-directional | Appearance | Facebook | r | Body esteem, reverse coded (BESAA) | 6 |
| Prieler et al., 2021 - Spain sample | Spain | Students (*n* = 306, 11-15, *M* = 13.46, *SD* = 1.10, 100%) | Tendency | Non-directional | Appearance | Facebook | r | Body esteem, reverse coded (BESAA) | 6 |
| Prieler et al., 2021 - South Korea sample | South Korea | Students (*n* = 184, 11-15, *M* = 13.46, *SD* = 1.10, 100%) | Tendency | Non-directional | Appearance | Facebook | r | Body esteem, reverse coded (BESAA) | 6 |
| Primo-Simoes et al., 2025 | Portugal | Sample recruited online via e-mail and social media (*n* = 322, 18-57, *M* = 26.25, *SD* = 8.19, 100%) | Self-evaluation | Bipolar | Appearance | Multiple | r | Dysmorphic concern (DCQ)  Body appreciation, reverse coded (BAS-2) | 6 |
| Qi et al., 2024; Study 1 | China | Students (*n* = 609, *M* = 15.18, *SD* = 0.59, 50%) | Tendency | Upward | Unspecified | Unspecified | r | Negative affect (PANAS) | 8 |
| Rajaei & Abraham, 2024 | US | Active social media users recruited online and offline (*n* = 200, 18-44, 56%) | Self-evaluation | Upward | Unspecified | Multiple | r | Depression (Mental Health Well-being Assessment, Gupta & Sharma, 2021)  Anxiety (Mental Health Well-being Assessment, Gupta & Sharma, 2021) | 5 |
| Robinson et al., 2019 | US | Clinical vs. non-clinical (*n* = 504, 18-38, *M* = 20.4, *SD* = 3.70, 82%) | Tendency | Upward | Appearance | Unspecified | SMD | / | 3 (out of 4) |
| Rodgers et al., 2024 | US | Sample recruited through flyer distribution and social media (*n* = 192, 40-70+, *M* = 50.8, *SD* = 8.40, 100%) | Tendency | Upward | Appearance | Unspecified | r | Shape concern (EDE-Q, weight concern and shape concern subscales) | 7 |
| Rodgers & Nowicki, 2024 | US | Sample recruited through flyer distribution and social media (*n* = 192, ≥40, *M* = 51.03, *SD* = 8.65, 100%) | Tendency | Upward, downward | Appearance | Unspecified | r | Body dissatisfaction (EDI-BD) | 7 |
| Rousseau et al., 2017 | Belgium | Students (*n* = 1621, 12-19, *M* = 14.76, *SD* = 1.41, 48%) | Tendency | Non-directional | Unspecified | Facebook | r | Body dissatisfaction (BAT – Body dissatisfaction subscale)  Satisfaction with life (SWLS) | 4 |
| Saffran et al., 2016 | US | Clinical (*n* = 333, 18-56, *M* = 28.15, *SD* = 8.41, 98%) | Tendency | Upward | Eating disorder (weight, exercise, food) | Facebook | r | Eating disorder symptoms (EDE-Q)  Shape Concern (EDE-Q – shape concerns subscale) | 4 |
| Samra & Dryer, 2024 | Australia | Pregnant women (*n* = 225, 19-40, *M* = 31.91, *SD* = 4.39, 100%) | Self-evaluation | Upward | Unspecified | Unspecified | r | Depression (EPDS)  Eating disorder symptoms (EAT-26)  Negative body image (BIPS)  Social media addiction (BSMAS) | 8 |
| Samra et al., 2022 | Australia | Students (*n* = 119, *M* = 20.15, *SD* = 3.94, 45%) | Self-evaluation  Tendency | Upward  Upward | Unspecified | Unspecified | r | Depression (PROMIS, 8-item version)  Social media addiction (BSMAS) | 8 |
| Scheiber et al., 2023 | Germany and Austria | Sample recruited via an online panel (*n* = 647, *M* = 23.7, *SD* = 4.70, 55%) | Tendency | Upward | Physical appearance | Unspecified | r | Body dissatisfaction (EDI-BD) | 5 |
| Schettino et al., 2023 | Italy and Portugal | Sample recruited online through social network groups (*n* = 340, 18-30, *M* = 23.08, *SD* = 3.46, 74%) | Tendency | Non-directional | Appearance | Instagram | r | Body shame (OBCS) | 6 |
| Schmuck et al., 2019 | Germany | Sample recruited through a panel (*n* = 833, 16-65, *M* = 45.44, *SD* = 14.83, 54%) | Self-evaluation | Upward | Unspecified | Unspecified | r | Satisfaction with life (SWLS) | 6 |
| Scully et al., 2023 | Ireland | Students (*n* = 210, 12-17, *M* = 15.16, *SD* = 1.17, 100%) | Self-evaluation  Tendency | Bipolar  Non-directional | Physical appearance | Facebook | r | Body dissatisfaction (EDI-BD) | 4 |
| Seekis & Barker, 2022 | Australia | Students (*n* = 399, 17-25, *M* = 19.36, *SD* = 1.97, 100%) | Tendency | Upward | Appearance | Unspecified | r | Body dismorphic disorder (BICI) | 7 |
| Spitzer et al., 2023 – Facebook sample | US | Students (*n* = 358, *M* = 19.16, *SD* = 1.61, 87%) | Self-evaluation | Bipolar | Unspecified | Facebook | r | Suicidal Ideation (DSI-SS) | 5 |
| Spitzer et al., 2023 – Instagram sample | US | Students (*n* = 449, *M* = 19.16, *SD* = 1.61, 87%) | Self-evaluation | Bipolar | Unspecified | Instagram | r | Suicidal Ideation (DSI-SS) | 5 |
| Steers et al., 2014; Study 1 – females | US | Students (*n* = 107, 100%) | Tendency | Non-directional | Unspecified | Facebook | r | Depression (CES-D) | 5 |
| Steers et al., 2014; Study 1 – males | US | Students (*n* = 26, 0%) | Tendency | Non-directional | Unspecified | Facebook | r | Depression (CES-D) | 5 |
| Stefana et al., 2022 | Italy | Population-based (*n* = 1172, 18-35, *M* = 26.3, *SD* = 4.90, 100%) | Tendency | Non-directional | Physical appearance | Instagram | r | Depression (CES-D)  Eating disorder symptoms (EAT-26) | 5 |
| Sun et al., 2023 | China | Students (*n* = 778, *M* = 13.01, *SD* = 0.49, 47%) | Self-evaluation | Upward | Unspecified | Wechat Moments | r | Eating disorder symptoms (TFEQ-R18) | 8 |
| Tarabay et al., 2023 | Lebanon | Teenagers (*n* = 379, 13-17, *M* = 16.07, *SD* = 1.19, 65%) | Self-evaluation | Upward | Unspecified | Unspecified | r | Addictive consequences (PUS) | 8 |
| Teo & Collinson, 2019 | Singapore | Instagram users recruited via e-mail and social media (*n* = 358, 18-34, *M* = 22.69, *SD* = 2.10, 69%) | Self-evaluation  Tendency | Bipolar  Non-directional | Unspecified | Instagram | r | Eating disorder symptoms (EAT-26) | 5 |
| Thompson et al., 2023 | US | Mothers (*n* = 347, 40-63, *M* = 50.13, *SD* = 4.64, 100%) | Tendency | Non-directional | Body/appearance, romantic relationship, children, profession/career achievements, and overall life | Unspecified | r | Bulimia (EDI-B)  Dietary restraint (EDE-Q, restraint subscale)  Eating disorder symptoms (EAT-26) | 4 |
| Tosun & Kasdarma, 2020 | Turkey | Students (*n* = 319, 52%) | Tendency | Upward | Unspecified | Facebook | r | Depression (STDI) | 6 |
| Van Oosten et al., 2023 | Netherlands | Sample recruited through a panel (*n* = 1852, 13-25, *M* = 18.61, 53%) | Tendency | Non-directional | Physical appearance | Multiple | r | Depression (MHI-5)  Body dissatisfaction (BAT) | 4 |
| Walker et al., 2015 | US | Students (*n* = 128, 18-23, 100%) | Tendency | Non-directional | Appearance | Facebook | r | Depression (BDI-II)  Eating disorder symptoms (EDE-Q)  Anxiety (STAI) | 5 |
| Wang et al., 2020 | China | Married adults (*n* = 514, 20-59, 62%) | Tendency | Upward | Unspecified | Unspecified | r | Depression (CES-D) | 7 |
| Wang et al., 2022 - male sample | China | Students (*n* = 196, 0%) | Tendency | Non-directional | Appearance | Unspecified | r | Body dissatisfaction(MBSRQ-BASS) | 6 |
| Wang et al., 2022 - female sample | China | Students (*n* = 280, 100%) | Tendency | Non-directional | Appearance | Unspecified | r | Body dissatisfaction(MBSRQ-BASS) | 6 |
| Wang et al., 2023 - female sample | China | Students (*n* = 430, 100%) | Tendency | Non-directional | Appearance | Unspecified | r | Facial dissatisfaction (NPSS-FAC) | 5 |
| Wang et al., 2023 - male sample | China | Students (*n* = 454, 0%) | Tendency | Non-directional | Appearance | Unspecified | r | Facial dissatisfaction (NPSS-FAC) | 5 |
| Wang et al., 2025 | China | Students (*n* = 795, 17-25, *M* = 20.17, *SD* = 1.65, 60%) | Tendency | Non-directional | Appearance | Unspecified | r | Body shame (OBCS, body shame subscale) | 6 |
| Xu & Chen, 2025 | China | Students (*n* = 2000, *M* = 20.94, *SD* = 3.46, 39%) | Tendency | Non-directional | Appearance | Unspecified | r | Eating disorder symptoms (EDI) | 6 |
| Xu et al., 2023 | China | Sample recruited online via social media (*n* = 807, 49%) | Tendency | Upward | Physical appearance | Multiple | r | Body dissatisfaction (items adapted from the NPSS and BCS) | 5 |
| Yan et al., 2024 | China | Students (*n* = 568, 18–24, 78%) | Tendency | Upward | Unspecified | Unspecified | r | Depression (PHQ-9) | 7 |
| Yang et al., 2020 | China | Students (*n* = 481, 17-22, *M* = 19.44, *SD* = 1.18, 58%) | Tendency | Non-directional | Appearance | Unspecified | r | Facial dissatisfaction (NPSS-FAC) | 5 |
| Yang et al., 2025 | US | Sample recruited online via Qualtrics survey panel (*n* = 2105, 12-18, *M* = 15.39, *SD* = 1.82, 49%) | Tendency | Non-directional | Unspecified | Multiple | r | Depression (CES-D, short form)  Anxiety (GAD-7) | 6 |
| Yao et al., 2021 | China | Students (*n* = 567, 17-23, *M* = 19.97, *SD* = 1.37, 100%) | Tendency | Non-directional | Appearance | Unspecified | r | Body shame (OBCS, body shame subscale)  Restrained eating (DEBQ, restrained eating subscale) | 6 |
| Yayun et al., 2024 | China | Students (*n* = 333, 18-26, *M* = 21.9, *SD* = 3.47, 67%) | Tendency | Non-directional | Appearance | Unspecified | r | Concern about appearance (items adapted from the NPSS and CONAPP) | 6 |
| Yu & Cingel, 2025 | US | Students (*n* = 462, 18-42, *M* = 19.75, *SD* = 2.09, 76%) | Motives | Upward | Unspecified | Unspecified | r | Negative affect (3-items scale)  Positive affect (3-items scale)  Satisfaction with life (SWLS) | 5 |
| Zhang et al., 2023 | China | Students (*n* = 350, 15-17, *M* = 16.07, *SD* = 0.70, 45%) | Self-evaluation | Upward | Unspecified | Unspecified | r | Anxiety (DASS-21, anxiety subscale)  Positive affect (WHO Happiness Indicator Scale, Topp et al., 2015) | 8 |
| Zhang et al., 2025 | China | Students (*n* = 789, *M* = 19.63, *SD* = 1.20, 41%) | Tendency | Upward | Unspecified | Unspecified | r | Social anxiety (SCS – social anxiety subscale) | 8 |
| Zhang et al., 2026 | China | Sample recruited online via social media (*n* = 378, 18-50+, 100%) | Tendency | Non-directional | Appearance | Unspecified | r | Facial dissatisfaction (NPSS-FAC) | 5 |
| Zhong, 2022 | South Korea | Students (*n* = 367, 100%) | Tendency | Non-directional | Appearance | Xiaohongshu | r | Body image disturbance (AAS) | 5 |
| Zuo & Zan, 2025 | China | Students (*n* = 500, 53%) | Tendency | Upward | Unspecified | Unspecified | r | Satisfaction with life (SWLS) | 8 |

*Note*. Abbreviations: AAS = Appearance Anxiety Scale-Brief Version ( Dion et al.,1990); ABQ = Athlete Burnout Questionnaire (Raedeke & Smith, 2001, 2009); ADS = Alcohol Dependence Scale (Skinner & Horn, 1982); BAS-2 = Body Appreciation Scale-2 (Tylka & Wood- Barcalow, 2015); BAT = Body Attitude Test (Probst et al., 1995); BCS = Body Comparison Scale (Dijkstra & Barelds, 2011); BDI = Beck Depression Inventory (Beck, 1987); BDI-II = Beck Depression Inventory version 2 (Beck et al., 1996); BDS = Body Dissatisfaction Scale (Mutale et al. 2016); BESAA = Body Esteem Scale for Adolescents and Adults (Mendelson et al. 2001); BICI = Body Image Concern Inventory (Littleton et al., 2005); BIPS = Negative Body Image in Pregnancy Scale (Watson et al., 2017); BISS = Body Image States Scale (Cash et al., 2002); BSI = Brief Symptom Inventory (Derogatis & Melisaratos, 1983); BSMAS = Bergen Social Media Addiction Scale (Andreassen et al., 2016); BSQ-14 = Body Shape Questionnaire-14 (Dowson & Henderson, 2001); BSS = Body Shape Satisfaction Scale (Pingitore et al., 1997); BUT-A GSI = Body Uneasiness Test A Global Severity Index (Cuzzolaro et al., 2006); CES-D = Center for Epidemiologic Studies Depression Scale (Radloff, 1977); CONAPP = Questionnaire for Assessing Concern About Appearance on Social Networks (Gonzalez-Nuevo et al., 2021); DASS-21 = Depression Anxiety Stress Scale (Lovibond & Lovibond, 1995); DCQ = Dysmorphic Concern Questionnaire (Oosthuizen et al. 1998); DEBQ = Dutch Eating Behavior Questionnaire (van Strien et al., 1986); DSI-SS = Depressive Symptom Inventory Suicidality Subscale (Metalsky & Joiner, 1997); EAT-26 = Eating Attitudes Test (Garner et al., 1982); (EDE-Q = Eating Disorder Examination Questionnaire (Fairburn & Beglin, 2008); EDI = Eating Disorder Inventory (Garner et al., 1983); EDI-B = Eating Disorder Inventory (Garner et al., 1983) - subscale bulimia; EDI-BD = Eating Disorder Inventory (Garner et al., 1983) - subscale body dissatisfaction; EDI-DT = Eating Disorder Inventory (Garner et al., 1983) - subscale drive for thinness; EPDS = Edinburgh Postnatal Depression Scale (Cox et al., 1987); GAD-7 = Generalized Anxiety Disorder-7 (Spitzer et al., 2006); HADS = Hospital Anxiety & Depression Scale (Zigmond & Snaith, 1983); ISI = Insomnia Severity Index (Bastien et al., 2001); MBSRQ-AE = Multidimensional Body-Self Relations Questionnaire-Appearance Evaluation scale (Cash, 2000); MBSRQ-BASS = Multidimensional Body-Self Relations Questionnaire-Body Areas Satisfaction Scale (Cash, 2000); MHI-5 = Mental Health Inventory (Rumpf et al., 2001); NPSS-FAC = Negative Physical Self Scale-Facial Appearance Concern subscale (Chen et al., 2006); OBCS = Objectified Body Consciousness Scale (McKinley & Hyde, 1996); OHQ-S = Oxford Happiness Questionnaire Short Scale (Hills & Argyle, 2002); PANAS = Positive and Negative Affect Schedule (Watson et al., 1988); PCPS-BI = Perfectionistic Self-Presentation Scale – Body Image (Ferreira et al. 2018); PHQ = Patient Health Questionnaire (Strine et al., 2008); PROMIS = Patient-Reported Outcomes Measurement System scale (Cella et al., 2010); PUS = Problematic Use of Social Networks Scale (González-Nuevo et al., 2021); QDC = Questionario sul Dismorfismo Corporeo (English translation: “Body Dysmorphic Disorder Questionnaire”, Cerea et al., 2017); SBQ-R = Revised Suicidal Behaviors Questionnaire (Osman et al., 2001); SCS = Self-Consciousness Scale (Fenigstein et al.,1975); SDQ = Strengths and Difficulties Questionnaire (Goodman et al., 2000); SLSS = Student Life Satisfaction Scale (Huebner, 1991); SMFQ-C = Short Mood and Feelings Questionnaire-Child (Angold et al., 1995); STAI = State-Trait Anxiety Inventory (Spielberger et al., 1970); STDI = State-Trait Depression Inventory (Spielberger et al., 2003); SWLS = Satisfaction With Life Scale (Diener et al., 1985); TFEQ-R18 = revised 18-item Three-Factor Eating Questionnaire (Karlsson et al., 2000); YSR = Youth Self Report (Achenbach et al., 2004)

^a^ quality criterion c was not rated for clinical vs. non-clinical comparisons; see Appendix Quality Assessment

**Appendix B – Characteristics of experimental studies**

| Reference | Country | Sample (n, age range, mean age, percentage female) | Manipulations for  experimental groups (EG) vs. control groups (CG) | Comparison category  /categories for meta-analysis | Direction of social comparison manipulation(s) in experimental groups | Social comparison dimension | Experimental stimuli presented on the following social media platform | Outcome variable (measure) | Study quality sum score (max-imum of 8, unless indica-ted other-wise) |
| --- | --- | --- | --- | --- | --- | --- | --- | --- | --- |
| Anixiadis et al., 2019 | Multiple | Sample recruited online via Prolific Academic (*n* = 126, 18-29, *M* = 23.28, *SD* = 2.58, 100%) | Two groups: exposure to thin-idealized images of users’ bodies (EG)  vs. scenery images (CG) | EG vs. CG | EG: upward | appearance | Instagram | Bipolar body dissatisfaction (VAS)  Negative mood (VAS) | 8 |
| Barron et al., 2021; Study 1 | USA | Students (*n* = 180, 18-33, *M* = 19.21, *SD* = 1.81, 65%) | Four groups: exposure to either same-gender images of users portraying fitspiration (EG1), users portraying self-compassion (EG2), users portraying a combination of fitspiration and self-compassion (EG3),  vs. architectural images (CG) | EG vs. EG  EG vs. CG | EG1: upward  EG2: positive body image interventions  EG3: positive body image interventions | appearance | Instagram | Bipolar body satisfaction, reverse coded (VAS) | 8 |
| Barron et al., 2021; Study 2 | USA | Sample recruited online via Mturk  (*n* = 296, 20-30,  *M* = 26.79, *SD* = 2.90, 100%) | Four groups: exposure to same-gender images of users portraying fitspiration (EG1), users portraying self-compassion (EG2), users portraying a combination of fitspiration and self-compassion (EG3),  vs. architectural images (CG) | EG vs. EG  EG vs. CG | EG 1: upward  EG 2: positive body image interventions  EG 3: positive body image interventions | appearance | Instagram | Bipolar body satisfaction, reverse coded (VAS) | 8 |
| Boehm et al., 2022; Study 1 | USA | Students (*n* = 214, *M* = 19.35, *SD* = 5.65, 84.10%) | Three groups: exposure to fortunate written posts (EG1), unfortunate written posts (EG2),  vs. neutral written posts (EG3) | EG vs. EG | EG 1: upward  EG 2: downward  EG 3: not clearly directed | Not specified | Facebook | Affect balance (i.e. difference between self-reported  positive and negative states; items similar to Scale of Positive and Negative Experience) | 8 |
| Boehm et al., 2022; Study 2 | USA | Students (*n*= 201,  *M* = 19.15, *SD* = 0.95, 63.18%) | Two groups: exposure to fortunate written posts, spontaneous or instructed social comparison (EG1) vs. unfortunate written posts, spontaneous or instructed social comparison (EG2). | EG vs. EG | EG 1: upward  EG 2: downward | Not specified | Facebook | Affect balance (i.e. difference between self-reported  positive and negative states; items similar to Scale of Positive and Negative Experience) | 8 |
| Brabandere et al., 2025; Study 1 | Belgium | Students  (*n* = 221,  14-18,  *M* = 16.22,  *SD* = 1.16,  62%) | Two groups: exposure to videos emphasizing same-gender fitfluencers’ body and muscle posing (EG1), vs. fitfluencers’ workouts and exercise (EG2) | EG vs. EG | EG 1: upward  EG 2: upward | appearance | TikTok | Bipolar body satisfaction (Cash et al., 2002), reverse coded | 8 |
| Brabandere et al., 2025; Study 2 | Belgium | Students  (*n* = 176,  14-18,  *M* = 14.94,  *SD* = 0.83,  38.6%) | Two groups: exposure to videos emphasizing same-gender fitfluencers’ body and muscle posing (EG1), vs. lifehack videos (CG) | EG vs. CG | EG: upward | appearance | TikTok | Bipolar body satisfaction (Cash et al., 2002), reverse coded | 8 |
| Brabandere et al., 2025; Study 3 | Belgium | Students  (*n* = 154,  14-18,  *M* = 17.06,  *SD* = 0.67,  38.3%) | Three groups: exposure to videos emphasizing same-gender fitfluencers’ body and muscle posing (EG1), authenticity fitfluencer videos (EG2), vs. lifehack videos (CG) | EG vs. EG  EG vs. CG | EG 1: upward  EG 2: positive body image interventions | appearance | TikTok | Bipolar body satisfaction (Cash et al., 2002), reverse coded | 8 |
| Breves et al., 2024 | Germany | Instagram users  (*n* = 175, 16-29,  *M* = 23.05, *SD* = 9.46, 100%) | Four groups: exposure to thin-ideal influencer body images with positive peer comments (EG1), thin-ideal influencer body images without positive peer comments (EG2), larger-sized influencer body images with positive peer comments (EG3), vs.  larger-sized influencer body images without positive peer comments (EG4) | EG vs. EG | EG 1: upward  EG 2: upward  EG 3: positive body image interventions  EG 4: positive body image interventions | appearance | Instagram | Actual-ideal body discrepancy (Female Figure Rating Scale)  Negative affect (PANAS)  Positive affect (PANAS) | 8 |
| Brown & Tiggemann, 2016 | Australia | Undergraduate students  (*n* = 138,  18-30  *M* = 20.10,  *SD* = 2.61,  100%) | Three groups:  Exposure to attractive celebrity images (EG1), attractive peer images (EG2), vs. images of travel destinations (CG) | EG vs. EG  EG vs. CG | EG 1: upward  EG 2: upward | appearance | Instagram | Negative mood (VAS)  Body dissatisfaction (VAS) | 8 |
| Burnell et al., 2020 | USA | Psychology students (*n* = 405, 18-25, *M* = 20.05, *SD* = 1.62, 84%) | Three groups: exposure to same-gender influencer Instagram profile (EG1), Instagram profile of a same-gender acquaintance  (EG2), vs. own Instagram profile (CG) | EG vs. EG  EG vs. CG | EG 1: upward  EG 2: upward | appearance | Instagram | Positive affect (three items adapted from Gross (2009)) | 8 |
| Casale et al., 2021 - males | Italy | College students without Instagram accounts (*n* = 65, *M* = 23.29, *SD* = 1.77, 0%) | Two groups: exposure to appearance-focused real Instagram profiles of attractive same-sex user (EG)  vs. re-assessment only control group, i.e., not exposed to Instagram (CG) | EG vs. CG | EG: upward | appearance | Instagram | Men: Body fat dissatisfaction (FSM)  Men: Muscular dissatisfaction (MSM) | 8 |
| Casale et al., 2021 - females | Italy | College students without Instagram accounts (*n* = 65, *M* = 23.22, *SD* = 1.73, 100%) | Two groups: exposure to appearance-focused real Instagram profiles of attractive same-sex user (EG)  vs. re-assessment only control group, i.e., not exposed to Instagram (CG) | EG vs. CG | EG: upward | appearance | Instagram | Women: Body dissatisfaction (CDRS) | 8 |
| Castellanos Silva & Steins, 2023 | Germany | Sample recruited online via the LimeSurvey Platform, 22 of the participants were psychology students  (*n* = 226, 18-40,  *M* = 23.13, *SD* = 4.58, 82.30%) | Two groups: exposure to same-gender hegemonic beauty ideal images (EG1)  vs. same-gender body diversity images (EG2) | EG vs. EG | EG 1: upward  EG 2: positive body image interventions | appearance | Instagram | Bipolar body dissatisfaction (VAS)  Bipolar mood (VAS) | 4 (out of 4)^a^ |
| Claeys et al., 2023; Study 1 | Belgium | Sample recruited online, followers of influencers (*n* = 171,  *M* = 23.44, *SD* = 4.64, 100%) | Two groups: exposure to post of female nongenuine influencer (EG1) vs. female genuine influencer (EG2) | EG vs. EG | EG 1: upward  EG 2: positive body image interventions | appearance | Instagram | Bipolar well-being (five items adopted from SWLS) | 4 (out of 4) |
| Claeys et al., 2023; Study 2 | Belgium | Sample recruited online, followers of influencers (*n* = 154,  *M* = 24.67, *SD* = 3.61, 51.30%) | Two groups: exposure to post of same-gender nongenuine influencer (EG1) vs. same-gender genuine influencer (EG2) | EG vs. EG | EG 1: upward  EG 2: positive body image interventions | appearance | Instagram | Bipolar well-being (five items adopted from SWLS) | 4 (out of 4) |
| Cohen et al., 2019 | Australia | Sample recruited via fliers and social media (*n* = 195, 18-30, *M* = 21.69, *SD* = 3.49, 100%) | Three groups: exposure to thin-idealized body images (EG1), body positivity images (EG2),  vs. nature/scenery images (CG) | EG vs. EG  EG vs. CG | EG 1: upward  EG 2: positive body image interventions | appearance | Instagram | Positive mood (VAS)  Negative mood (VAS) | 8 |
| Couture Bue & Harrison, 2020 | USA | Sample recruited via University of Michigan Health Research pool website  (n = 185, 18-35,  100%)  Eye-tracking  (n = 149,  18-35,  *M* = 23.42,  *SD* = 5.05,  100%) | Two groups: exposure to thin-ideal images with traditional comments that idealized the image (EG1), vs. thin-ideal images with disclaimer comments that critiqued the images as unrealistic (EG2) | EG vs. EG | EG 1: upward  EG 2: positive body image interventions | appearance | Instagram | Body anxiety (PASTAS)  Fixation duration (eye tracking) | 4 (out of 4) |
| Cowles et al., 2023 | UK | Young women (*n* = 195, 18-30, *M* = 21.21, *SD* = 2.90, 100%) | Three groups: exposure to thin-ideal body posts (EG1), body positive images and body positive captions (EG2), vs. body positive images without captions (EG3) | EG vs. EG | EG 1: upward  EG 2: positive body image interventions  EG 3: positive body image interventions | appearance | Instagram | Positive mood (VAS)  Negative mood (VAS)  Bipolar body satisfaction, reverse coded (VAS) | 4 (out of 4) |
| Davies et al., 2020 | UK | Instagram users  (*n* = 109,  18-25,  *M* = 21.58,  *SD* = 1.54,  100%) | Three groups: exposure to fitspiration images with fitspiration captions (EG1), fitspiration images with body positive captions (EG2), vs. fitspiration images with neutral captions (EG3) | EG vs. EG | EG1: upward  EG2: positive body image interventions  EG3: upward | appearance | Instagram | Positive mood (PANAS-SF)  Negative mood (PANAS-SF) | 8 |
| de Vries et al., 2018 | Netherlands | University students  (*n* = 126,  18-30,  *M* = 21.4,  *SD* = 2.4,  81%) | Two groups: exposure to posts with positively toned caption (EG1), vs. posts with negatively toned captions (EG2) | EG vs. EG | EG 1: upward  EG 2: downward | appearance | Instagram | Positive affect (PANAS)  Negative affect (PANAS) | 8 |
| Dhadly et al., 2023 | Canada | University students  (*n* = 156,  18-28  *M* = 18.44,  *SD* = 1.07,  100%) | Three groups: exposure to body-focused videos (EG1), body-positive videos (EG2), vs. #DIY, #howto, #nature videos (CG) | EG vs. EG  EG vs. CG | EG 1: upward  EG 2: positive body image interventions | appearance | TikTok | Positive affect  (PANAS)  Negative affect (PANAS)  Bipolar body image, reverse coded (BISS) | 8 |
| Di Michele et al., 2023; Study 1 | Italy | Sample recruited via social media (*n* = 356, 19-32, *M* = 24.98, *SD* = 3.69, 100%) | Three groups: exposure to sexualized beauty ideal (EG1), sexualized body positivity (EG2), vs. non-sexualized body positivity posts (EG3) | EG vs. EG | EG 1: upward  EG 2: positive body image interventions  EG 3: positive body image interventions | appearance | Instagram | Positive mood (VAS)  Negative mood (VAS) | 4 (out of 4) |
| Di Michele et al., 2023; Study 2 | Italy | Sample recruited via social media (*n* = 316, 19-32, *M* = 23.87, *SD* = 4.19, 100%) | Three groups: exposure to sexualized beauty ideal (EG1), sexualized body positivity (EG2), vs. non-sexualized body positivity videos (EG3) | EG vs. EG | EG 1: upward  EG 2: positive body image interventions  EG 3: positive body image interventions | appearance | TikTok | Positive mood (VAS)  Negative mood (VAS) | 4 (out of 4) |
| Dignard & Jarry, 2020 – Caucasian sample | Canada | University students (*n* = 225, 100%) | Three groups: exposure to fitspiration (EG1), thinspiration (EG2), vs. travel posts (CG) | EG vs. EG  EG vs. CG | EG 1: upward  EG 2: upward | appearance | Instagram | Depressive symptoms (BDI-II)  Bipolar body satisfaction, reverse coded (BISS) | 8 |
| Dignard & Jarry, 2020 – Non-Caucasian sample | Canada | University students (*n* = 106, 100%) | Three groups: exposure to fitspiration (EG1), thinspiration (EG2), vs. travel posts (CG) | EG vs. EG  EG vs. CG | EG 1: upward  EG 2: upward | appearance | Instagram | Depressive symptoms (BDI-II)  Bipolar body satisfaction, reverse coded (BISS) | 8 |
| Fardouly & Holland, 2018 | Australia | Sample recruited online via MTurk  (*n* = 164,  18-25,  *M* = 23.09,  *SD* = 1.69,  100%) | Three groups: exposure to images of an attractive woman (EG1), attractive woman with disclaimer comments (EG2), vs. travel images (CG) | EG vs. EG  EG vs. CG | EG 1: upward  EG 2: positive body image interventions | appearance | Instagram | Negative mood (VAS)  Bipolar body dissatisfaction (VAS) | 8 |
| Fardouly & Rapee, 2019 | Australia | Undergraduate students  (*n* = 175,  18-25,  *M* = 19.26,  *SD* = 1.55  100%) | Three groups: exposure to idealized images of women wearing make-up (EG1), images of women without make-up interspersed among images of women wearing make-up (EG2), travel-focused images (CG) | EG vs. EG  EG vs. CG | EG 1: upward  EG 2: positive body image interventions | appearance | Instagram | Negative mood (VAS)  Bipolar appearence satisfaction, reverse coded (VAS)  Bipolar facial satisfaction, reverse coded (VAS) | 8 |
| Fardouly et al., 2023 | Australia | Facebook users (*n* = 159, 18-25, *M* = 20.92, *SD* = 2.21, 100%) | Three groups: two-week exposure to daily posts in body positive Facebook group (EG), appearance neutral (e.g. travel, nature, science) Facebook group (CG1), vs. usual/unrestricted Facebook usage (CG2) | EG vs. CG | EG: positive body image interventions | appearance | Facebook | Body dissatisfaction (subscale of EDI)  Positive affect  (PANAS-SF)  Negative affect (PANAS-SF) | 4 (out of 4) |
| Gurtala & Fardouly, 2023 | Australia | Undergraduate psychology students  (*n* = 211,  17-28,  *M* = 19.19,  *SD* = 1.80,  100%) | Four groups: exposure to appearance-ideal images (EG1), appearance-ideal videos (EG2), appearance-neutral images (CG1), appearance-neutral videos (CG2) | EG vs. EG  EG vs. CG | EG 1: upward  EG 2: upward | appearance | Instagram  TikTok | Bipolar appearance satisfaction, reverse coded (VAS)  Negative mood (VAS) | 8 |
| Haferkamp & Krämer, 2011 | Germany | Unspecified sample  (*n* = 91,  *M* = 22.53,  *SD* = 2.75,  52.7% | Two groups: exposure to profiles showing very attractive users (EG1), vs. Profiles showing unattractive users (EG2) | EG vs. EG | EG 1: upward  EG 2: downward | appearance | Hot or not | Positive affect  (PANAS)  Real-ideal body discrepancy  (Body image scale by Luczak, 1999) | 8 |
| Hogue & Mills, 2019 | Canada | University students (*n* = 118, 17-27, *M* = 19.59, *SD* = 2.00, 100%) | Two groups: exposure to + commenting a more attractive female peer profile (EG1), vs. a non-peer, not-more-attractive family member profile (EG2) | EG vs. EG | EG 1: upward  EG 2: not clearly directed | appearance | Facebook, Instagram | Body dissatisfaction (VAS-BD) | 6 |
| Jia, 2025 | China | Unspecified sample  (*n* = 479,  15-62,  *M* = 33.5,  *SD* = 8.45,  56.6%) | Two groups: social media posts of strangers focusing on portraits featuring scenes from daily life (EG), vs. social media posts of strangers focusing on objects (CG). | EG vs. CG | EG: upward | appearance | Not specified | Online social anxiety (SAS-SMU) | 8 |
| Joiner et al., 2023; Study 1 | United Kingdom | Sample recruited online  (*n* = 262,  18-25,  *M* = 20.09,  *SD* = 2.45,  100%) | Three groups: exposure to thin dancer videos (EG1), large dancer videos (EG2), animal videos (CG) | EG vs. EG  EG vs. CG | EG 1: upward  EG 2: positive body image interventions | appearance | TikTok | Bipolar body satisfaction, reverse coded (VAS) | 8 |
| Joiner et al., 2023; Study 2 | United Kingdom | Sample recruited online  (*n* = 280,  18-25,  *M* = 21.29,  *SD* = 1.80,  100%) | Three groups: exposure to thin dancer videos (EG1), large dancer videos (EG2), mixture of animal videos and videos of people dressed in oversize animal costumes (CG) | EG vs. EG  EG vs. CG | EG 1: upward  EG 2: positive body image interventions | appearance | TikTok | Bipolar body satisfaction, reverse coded (VAS) | 8 |
| Joiner et al., 2023; Study 3 | United Kingdom | Sample recruited online  (*n* = 375,  18-25,  *M* = 22.54,  *SD* = 1.84,  100%) | Two groups: exposure to thin dancer videos (EG1), vs. large dancer videos (EG2) | EG vs. EG | EG 1: upward  EG 2: positive body image interventions | appearance | TikTok | Bipolar body satisfaction, reverse coded (VAS) | 4 (out of 4) |
| Kharkwal et al., 2024 | USA | University students (*n* = 217, *M* = 20.80, *SD* = 1.87, 100%) | Four groups: exposure to Instagram gym advertisement showing a thin-sized model + physique slogan (EG1), thin-sized model + health and wellness slogan (EG2), plus-size model + physique slogan (EG3), vs. plus-size model + health and wellness slogan (EG4) | EG vs. EG | EG 1: upward  EG 2: upward  EG 3: positive body image interventions  EG 4: positive body image interventions | appearance | Instagram | Bipolar body satisfaction, reverse coded (VAS) | 8 |
| Kilby & Mickelson, 2025 | USA | Sample recruited online  (*n* = 326,  18-67,  *M* = 35.01,  *SD* = 14.96,  61.3%) | Four groups: exposure to thin-ideal videos (EG1), body positivity videos (EG2), body neutrality videos (EG3), vs. travel videos (CG) | EG vs. EG  EG vs. CG | EG 1: upward  EG 2: positive body image interventions  EG 3: positive body image interventions | appearance | TikTok | Body dissatisfaction (VAS) | 8 |
| Kleemans et al., 2016 | Nether-lands | Students (*n* = 144, 14-18, *M* = 15.92, *SD* = 1.16, 100%) | Two groups: exposure to manipulated (retouched and reshaped) selfies (EG1), vs. original selfies (EG2) | EG vs. EG | EG 1: upward  EG 2: positive body image interventions | appearance | Instagram | Bipolar body satisfaction, reverse coded (BISS) | 4 (out of 4) |
| Leung & MacDonald, 2022 | Canada | Undergraduate single students  (*n* = 272,  *M* = 18.52,  *SD* = 2.29,  100%) | Three groups: browsing through the Facebook profile of a close friend (EG1), a friend (EG2), vs. an acquaintance (EG3) known to be in a relationship | EG vs. EG | EG 1: upward  EG 2: upward  EG 3: upward | Relationship status | Facebook | Positive Affect (PANAS)  Negative Affect (PANAS) | 8 |
| Livingston et al., 2020 | Australia | Psychology students (*n* = 201, 18-25, *M* = 18.93, *SD* = 1.24, 100%) | Three groups: exposure to images of an attractive woman only (EG1), an attractive woman with disclaimer (EG2), vs. landscapes (CG) | EG vs. EG  EG vs. CG | EG 1: upward  EG 2: positive body image interventions | appearance | Instagram | Bipolar body dissatisfaction (VAS)  Negative mood (VAS) | 8 |
| Lowe-Calverley & Grieve, 2021 | Australia | Australian female participants  (*n* = 111,  17-40  *M* = 23.39,  *SD* = 6.49,  100%) | Three groups:  Exposure to images of high-popularity female influencers (EG1), images of low-popularity female influencers (EG2), vs. Nature images (CG) | EG vs. EG  EG vs. CG | EG 1: upward  EG 2: upward | appearance | Instagram | Negative mood (VAS)  Body dissatisfaction  (VAS) | 8 |
| Manning & Mulgrew, 2022 | Australia | Mainly university students (*n* = 233, 18-30, *M* = 22.5, *SD* = 3.52, 100%) | Three groups: exposure to non-idealized body positivity posts (EG1), body positivity posts with captions (EG2), vs. cityscapes (CG) | EG vs. EG  EG vs. CG | EG 1: positive body image interventions  EG 2: positive body image interventions | appearance | Instagram | Positive mood (VAS) | 4 (out of 4) |
| McComb & Mills, 2021 | Canada | Psychology students (*n* = 142, 18-24, *M* = 19.06, *SD* = 1.34, 100%) | Two groups: forced comparison of own body parts with thin idealized images (EG), vs. forced art critique task of landscape painting images (CG) | EG vs. CG | EG: upward | appearance | Instagram | Body dissatisfaction (VAS) | 8 |
| McComb & Mills, 2022 | Canada | Psychology students (*n* = 402, 18-25, *M* = 19.53, *SD* = 1.74, 100%) | Four groups: exposure to thin-ideal (EG1), fit-ideal (EG2), slim-thick-ideal body type (EG3), vs. home décor/furniture images (CG) | EG vs. EG  EG vs. CG | EG 1: upward  EG 2: upward  EG 3: upward | appearance | Instagram | Bipolar body dissatisfaction (VAS)  Bipolar weight dissatisfaction (VAS) | 8 |
| McComb et al., 2021 | Canada | University students  (*n* = 311,  18-25,  *M* = 19.13,  *SD* = 1.43,  100%) | Four groups: exposure to thin-ideal images (EG1), thin-ideal images + generic caption (EG2), thin-ideal images + specific caption (EG3), vs. thin-ideal images + warning caption (EG4) | EG vs. EG | EG 1: upward  EG 2: positive body image interventions  EG 3: positive body image interventions  EG 4: positive body image interventions | appearance | Instagram | Bipolar body satisfaction, reverse coded (BISS)  Happiness (VAS)  Depression (VAS) | 4 (out of 4) |
| Morry et al., 2018; study 2 | Canada | Students  (*n* = 180,  *M* = 19.56,  *SD* = 2.76,  62.78%) | Two groups: exposure to a positive relationship Facebook profile (EG1) vs. less positive relationship Facebook profile (EG2) | EG vs. EG | EG 1: upward  EG 2: downward | Relationship quality | Facebook | Bipolar life satisfaction (5-item life satisfaction scale; Diener et al., 1985) | 8 |
| Mulgrew & Courtney, 2022 | Australia | University students  (*n* = 318,  17-30  *M* = 22.19,  *SD* = 3.86,  100%) | Four groups: exposure to fitspiration images (EG1), fitspiration images accompanied by captions showing appreciation for the body’s functionality (EG2), images of women of various body sizes accompanied by captions showing appreciation for the body’s functionality (EG3), vs. images of cityscapes (CG) | EG vs. EG  EG vs. CG | EG 1: upward  EG 2: positive body image interventions  EG 3: positive body image interventions | appearance | Instagram | Bipolar appearence satisfaction (VAS), reverse coded | 8 |
| Nelson et al., 2022 | USA | Samples recruited online via Amazon MTurk  (*n* = 205,  18-76,  *M* = 40.97,  *SD* = 13.41,  100%) | Three groups: exposure to thin-ideal images (EG1), body-positive images (EG2), vs. images of architecture or landscapes (CG) | EG vs. EG  EG vs. CG | EG 1: upward  EG 2: positive body image interventions | appearance | Instagram | Bipolar body satisfaction, reverse coded  (VAS) | 8 |
| Nerini et al., 2022 | Italy | Instagram users (*n* = 105, 18-30, *M* = 24.3, *SD* = 3.47, 100%) | Three groups: exposure to enhanced image of a woman (EG1), enhanced and original images of a woman with disclaimer (EG2), vs. a dog (CG) | EG vs. EG  EG vs. CG group | EG 1: upward  EG 2: positive body image interventions | appearance | Instagram | Body dissatisfaction (BSQ-14) | 8 |
| Politte-Corn & Fardouly, 2020 | USA | Sample recruited online via M-Turk (*n* = 394, 18-25, *M* = 23.29, *SD* = 1.60, 100%) | Four groups: exposure to idealized images with  appearance-related comments (EG1), idealized images with appearance-neutral comments (EG2), no-makeup  images with appearance-related comments (EG3), vs. no-makeup images with appearance-neutral com-  ments (EG4) | EG vs. EG | EG 1: upward  EG 2: upward  EG 3: positive body image interventions  EG 4: positive body image interventions | appearance | Instagram | Negative mood (VAS)  Appearance discrepancy (state version of the SDI) | 8 |
| Prichard et al., 2020 | Australia | University students (*n* = 108, 17-25, *M* = 20.24, *SD* = 1.86, 100%) | Two groups: exposure to fitspiration (EG), vs. travel destination posts (CG) | EG vs. CG | EG: upward | appearance | Instagram | Negative mood (VAS)  Bipolar body dissatisfaction (VAS) | 8 |
| Prichard et al., 2023 | Australia | Undergraduate psychology students and Facebook users  (*n* = 230, 17-25,  *M* = 20.46,  *SD* = 2.31,  100% | Three groups: exposure to images of female Influencers wearing fashionable attire (EG1), sexualized images revealing clothing such as lingerie and bikinis (EG2), vs. images of fashion products (CG) | EG vs. EG  EG vs. CG | EG 1: upward  EG 2: upward | appearance | Instagram | Negative mood (VAS)  Bipolar body dissatisfaction (VAS) | 8 |
| Pryde & Prichard, 2022 | Australia | Sample recruited online & Psychology students  (*n* = 120,  17-25,  *M* = 21.06,  *SD* = 2.40,  100%) | Two groups: exposure to fitspiration videos (EG) vs. art videos (CG) | EG vs. CG | EG: upward | appearance | TikTok | Body dissatisfaction (VAS)  Negative mood (VAS) | 8 |
| Rutter et al., 2023; Study 1 | USA | Sample recruited online via MTurk  (*n* = 105, 19-26,  *M* = 23.00, *SD* = 1.70, 100%) | Three groups: exposure to fitspiration body photos (EG1), body-positive body photos (EG2), vs. landscape posts (CG) | EG vs. EG  EG vs. CG | EG 1: upward  EG 2: positive body image interventions | appearance | Instagram | Negative mood  (PANAS)  Positive mood (PANAS) | 8 |
| Rutter et al., 2023; Study 2 | USA | Sample recruited online via MTurk  (*n* = 105, 21-30,  *M* = 26.82, *SD* = 2.44, 100%) | Three groups: exposure to appearance-ideal makeup faces (EG1), no-makeup faces (EG2), vs. landscapes (CG) | EG vs. EG  EG vs. CG | EG 1: upward  EG 2: positive body image interventions | appearance | Instagram | Negative mood  (PANAS)  Positive mood (PANAS) | 8 |
| Sampson et al., 2020 | United Kingdom | University students (*n* = 132, 18-35, *M* = 20.50, *SD* = 2.21, 60,60%) | Two groups: exposure to idealized smile images (EG), vs. nature images (CG) | EG vs. CG | EG: upward | appearance | Instagram | Bipolar facial dissatisfaction (Facial Satisfaction Scale)  Bipolar body dissatisfaction (Body Satisfaction Scale) | 8 |
| Seekis & Kennedy, 2023 | Australia | Psychology students  (*n* = 115,  17-25,  *M* = 19.37, *SD* = 1.89, 100%) | Three groups: exposure to videos showing beauty (EG1), self-compassion (EG2), vs. travel landscape content (CG) | EG vs. EG  EG vs. CG | EG 1: upward  EG 2: positive body image interventions | appearance | TikTok | Appearance anxiety (PASTAS)  Appearance shame (adapted version of PBSS-R)  Negative mood (VAS) | 8 |
| Seekis & Lawrence, 2023 | Australia | Psychology students  (*n* = 189, 17-28,  *M* = 19.25, *SD* = 1.98, 100.%) | Three groups: exposure to videos showing thin-ideal (EG1), body neutrality (EG2), vs. art content (CG) | EG vs. EG  EG vs. CG | EG 1: upward  EG 2: positive body image interventions | appearance | TikTok | Positive mood (VAS)  Bipolar body satisfaction, reverse coded (VAS) | 8 |
| Simon & Hurst, 2021 | UK | Sample recruited online via social media advertisements and university mailing lists  (*n* = 167, 18-50+, *M* = 27.74, 100%) | Three groups: exposure to posts with body-positivity content with average size model (EG1), body-positivity content with larger size model (EG2), vs. travel content (CG) | EG vs. EG  EG vs. CG | EG 1: positive body image interventions  EG 2: positive body image interventions | appearance | Instagram | Positive mood (PANAS)  Negative mood (PANAS)  Bipolar body satisfaction, reverse coded (BSS)  Food choice (calories)  Food choice (nutritious score) | 4 (out of 4) |
| Slater et al., 2019 | UK | Sample recruited via social media  (*n* = 102,  18-30,  *M* = 23.55,  *SD* = 2.33,  100%) | Two groups: exposure to images of thin-ideal celebrities (EG1), vs. parody images (EG2) | EG vs. EG | EG 1: upward  EG 2: positive body image interventions | appearance | Instagram | Negative mood “anxious” (VAS)  Positive mood “happy” (VAS) | 4 (out of 4) |
| Taylor &Armes, 2024 | UK | Psychology students  (*n* = 49,  18-35,  *M* = 20.00,  *SD* = 2.91,  87.75% | Three groups: exposure to images showing a toned body, luxurious holiday or happy couple + positive comment/ caption (EG1), images of someone crying or overweight individual + negative comment/ caption (EG2), vs. stone wall or people talking with no facial expressions visible + neutral comment/ caption (CG) | EG vs. EG  EG vs. CG | EG 1: upward  EG 2: downward | appearance | Instagram | Bipolar Body-esteem (BESAA), reverse coded | 8 |
| Tiggemann & Anderberg, 2020a | Australia | Sample recruited online via TurkPrime (*n* = 284, 18-30, *M* = 24.94, *SD* = 2.96, 0%) | Three groups: exposure to fashion images (EG1), fitspiration images (EG2), vs. scenery images (CG) | EG vs. EG  EG vs. CG | EG 1: upward  EG 2: upward | appearance | Instagram | Bipolar body satisfaction, reverse coded (VAS)  Bipolar facial satisfaction, reverse coded (VAS) | 8 |
| Tiggemann & Anderberg, 2020b | Australia | Sample recruited online via TurkPrime  (*n* = 305,  18-30,  *M* = 25.34,  *SD* = 2.98,  100%) | Three groups: exposure to idealized images (EG1), paired side-by-side images of idealized and real images (EG2), vs. real images (EG3) | EG vs. EG | EG 1: upward  EG 2: positive body image interventions  EG 3: positive body image interventions | appearance |  | Body dissatisfaction (VAS) | 4 (out of 4) |
| Tiggemann & Barbato, 2018 |  | University students  (*n* = 128,  18-30,  *M* = 20.12,  *SD* = 2.46,  100%) | Two groups: exposure to attractive full-body images of women paired with appearance-related comment (EG1), vs. same images paired with place-related comment (EG2) | EG vs. EG | EG 1: upward  EG 2: upward | appearance | Instagram | Body dissatisfaction (VAS) | 8 |
| Tiggemann & Zaccardo, 2015 | Australia | Students (*n* = 130, 17-30, *M* = 19.91, *SD* = 2.80, 100%) | Two groups: exposure to fitspiration (EG), vs. travel images (CG) | EG vs. CG | EG: upward | appearance | Instagram | Body dissatisfaction (VAS)  Negative mood (VAS) | 8 |
| Tiggemann & Zinoviev, 2019 | Australia | University students (*n* = 204, 17-30, *M* = 20.26, *SD* = 2.62, 100%) | Three groups: exposure to idealised attractive and thin images (EG1), enhancement-free images (EG2), vs. enhancement-free images with hashtags (EG3) | EG vs. EG | EG 1: upward  EG 2: positive body image interventions  EG 3: positive body image interventions | appearance | Instagram | Body dissatisfaction (VAS)  Facial dissatisfaction (VAS) | 4 (out of 4) |
| Tiggemann et al., 2018 | Australia | University students  (*n* = 220,  18-30,  *M* = 20.13,  *SD* = 2.58,  100%) | Four groups: exposure to thin-ideal images + low likes (EG1), thin-ideal images + high-likes (EG2), average-size images + low likes (EG3), vs. average-size images (EG4) | EG vs. EG | EG 1: upward  EG 2: upward  EG 3: positive body image interventions  EG 4: positive body image interventions | appearance | Instagram | Body dissatisfaction (VAS)  Facial dissatisfaction (VAS) | 8 |
| Tiggemann et al., 2020a | Australia | University students (*n* = 130, 17-29, *M* = 21.17, *SD* = 2.85, 100%) | Two groups: exposure to thin-sized (EG1), vs. average-sized women images (EG2) | EG vs. EG | EG 1: upward  EG 2: positive body image interventions | appearance | Instagram | Body dissatisfaction (VAS)  Facial dissatisfaction (VAS)  Negative mood (VAS)  Number of selfies taken  Time to take a selfie (in seconds)  Time spent editing the selfie (in seconds)  Extend of editing (score ranging between 0 – 1500) | 4 (out of 4) |
| Tiggemann et al., 2020b | Australia | University students  (*n* = 384,  17-30,  *M* = 20.27,  *SD* = 2.52,  100%) | Four groups: exposure to thin-ideal images + no caption (EG1), thin-ideal images + body-positive caption (EG2), average-size images + no caption (EG3), average-size images + body-positive caption (EG4) | EG vs. EG | EG 1: upward  EG 2: positive body image interventions  EG 3: positive body image interventions  EG 4: positive body image interventions | appearance | Instagram | Body dissatisfaction (VAS) | 4 (out of 4) |
| T´ng et al., 2024 | Singapore | University students (*n* = 80, *M* = 21.48, *SD* = 2.33 100%) | Two groups: exposure to similar (EG1), vs. dissimilar (EG2) influencer profiles in terms of both gender and race | EG vs. EG | EG 1: upward  EG 2: upward | appearance | Instagram | Satisfaction with life (Satisfaction With Life Scale) | 8 |
| Veldhuis et al., 2024 | Nether-lands | Adolescent girls from public secondary schools (*n* = 216, 11-18, *M* = 14.15, *SD* = 1.47, 0%) | Two groups: exposure to a screenshot of an extremely thin model + “3kg underweight” comment (EG1), vs. thin-normal model + “3kg underweight” comment (EG2) | EG vs. EG | EG 1: upward  EG 2: upward | appearance | YouTube | Bipolar body dissatisfaction (adapted version of the Body Dissatisfaction Subscale  from the Eating Disorder Inventory) | 8 |
| Vogel et al., 2015; Study 2 | USA | University students  (*n* = 120,  *M* = 18.93,  *SD* = 3.94,  76.67%) | Three groups: browsing through the Facebook profile of an acquaintance (EG), browsing through own Facebook profile (CG1), vs. Reading reviews of a consumer product (CG2) | EG vs. CG | EG: upward | appearance | Facebook | Affect balance (i.e., negative affect score subtracted from the positive affect score, PANAS) | 8 |
| Weber et al., 2025 | Germany | Sample recruited online  (*n* = 76,  18-63,  *M* = 23.0,  100%) | Two groups: exposure to #ThatGirl video clips (EG), vs. #nature video clips (CG) | EG vs. CG | EG: upward | appearance | TikTok | Positive affect (PANAS)  Negative affect (PANAS)  Bipolar body satisfaction (VAS), reverse coded | 8 |
| Wenhold et al., 2025 | USA | Sample recruited online  (*n* = 193,  19-43,  *M* = 31.65,  *SD* = 4.28,  100%) | Three groups: exposure to pregnancy/ shortly after giving birth fitspiration videos (EG1), body positivity videos showing post-partum body changes (EG2), vs. baby-led weaning videos (CG) | EG vs. EG  EG vs. CG | EG 1: upward  EG 2: positive body image interventions | appearance | TikTok | Bipolar body satisfaction (VAS), reverse coded  State anxiety (STAI short-form) | 8 |
| Wolfe & Yakabovits, 2022 | USA | University students (*n* = 95,  18-49, *M* = 21.51, *SD* = 4.93, 100%) | Three groups: exposure to edited images (EG1), unedited images (EG2), vs. animal/ objects/ scenery images (CG) | EG vs. EG  EG vs. CG | EG 1: upward  EG 2: positive body image interventions | appearance | SNS mock-up, not branded | Bipolar affect (VAS)  Bipolar physical attractiveness, reverse coded (VAS)  Number of photos taken  Edit time | 8 |
| Yang & Yin, 2023 | USA | Sample recruited online via MTurk  (*n* = 625, 18-72,  *M* = 34.56, *SD* = 9.46, 100%) | Four groups: exposure to tweets with images of thin and less muscular athletes (EG1), images of muscular and less thin athletes (EG2), images of combination of the conditions before (EG3), vs. images of cars (CG) | EG vs. EG  EG vs. CG | EG 1: upward  EG 2: upward  EG 3: upward | appearance | Twitter | Bipolar body satisfaction, reverse coded (items adapted from Bell et al., 2016) | 8 |

*Note.* Abbreviations: BASES = Body and appearance related self-conscious emotions scale (Castonguay et al., 2014); BDI-II = Beck Depression Inventar (Beck, Steer, Ball, & Ranieri, 1996); BESAA = Body-Esteem Scale for Adolescents and Adults (Mendelson et al., 1997); BISS = Body Image States Scale (Cash et al., 2002); BSQ-14 = Body Shape Questionnaire-14 items (Dowson & Henderson, 2001); BSS = Body Satisfaction Scale (Slade et al., 1990); CDRS = Contour Drawing Rating Scale (M. A. Thompson & Gray, 1995); EDI = Eating Disorder Inventory (Garner et al., 1983); FSM = Fat Silhouette Measure (Frederick et al., 2007); FSS = Facial Satisfaction Scale (Slade et al., 1990); MSM = Muscle Silhouette Measure (Frederick et al., 2007); PANAS = Positive and Negative Affect Schedule (Watson et al., 1988); PANAS-SF = Positive and Negative Affect Scale – Short Form (Watson et al., 1988); PANAS-X = Positive and Negative Affect Scale – Expanded Form (Watson & Clark, 1994); PASTAS = Physical Appearance State and Trait Anxiety Scale (Reed et al, 1991); PBSS = Phenomenological Body Shame Scale (Siegel et al., 2021); RSS = Relationship Satisfaction subscale of the Relationship Stability Scale (Rusbult et al., 1998); SAS-SMU = Social Anxiety Scale for Social Media Users (Alkis et al., 2017); SDI = State-Version des Self-Discrepancy Index (Dittmar et al., 1996; Fardouly et al., 2015; Halliwell & Dittmar, 2006); STAI short-form = six-item short-form of the Spielberger State—Trait Anxiety Inventory (Marteau & Bekker, 1992); SV = sozialer Vergleich; SWLS = The Satisfaction with Life Scale (Diener et al., 1985); VAS = Visual Analogue Scales (Heinberg & Thompson, 1995).

^a^quality criterion a was not rated for body positivity interventions; see Appendix Quality Assessment

**Appendix C – Social media platform-specific meta-analytic results for observational studies assessing the relationship between social comparison on social media and mental health outcomes**

| **Cross-sectional bivariate correlations**  **Component of the social comparison process: Tendency** | | | | | | | | | | | | | |
| --- | --- | --- | --- | --- | --- | --- | --- | --- | --- | --- | --- | --- | --- |
| Outcome  platform-specific results | *k* | *r* | 95% CI | *p* | 95% PI | *Q (p)* | | | *τ* | | | *I*² | *p*^a^ mod. by platform |
| **Upward tendency or non-directional tendency^b^** | | | | | | | | | | | | | |
| **Any psycho-pathology**  Any platform | 64 | **0.34** | 0.30; 0.38 | <.001 | -0.01; 0.62 | 848.69 (<.001) | | | 0.18 | | | 94.66 | 0.304 |
| Facebook | 17 | **0.29** | 0.20; 0.38 | <.001 | -0.10; 0.60 | 128.89 (<.001) | | | 0.20 | | | 90.28 | ref. group |
| Instagram | 10 | **0.40** | 0.31; 0.48 | <.001 | 0.13; 0.61 | 49.32  (<.001) | | | 0.14 | | | 85.58 | .113 |
| Multiple^c^ | 5 | **0.28** | 0.23; 0.32 | <.001 | 0.20; 0.36 | 9.20  (.056) | | | 0.04 | | | 53.66 | .877 |
| Unspecified^d^ | 32 | **0.36** | 0.30; 0.42 | <.001 | -0.02; 0.65 | 622.47 (<.001) | | | 0.20 | | | 96.28 | .194 |
| **Depression**  Any platform | 35 | **0.27** | 0.23; 0.30 | <.001 | 0.07; 0.44 | 217.39 (<.001) | | | 0.10 | | | 86.33 | .372 |
| Facebook | 13 | **0.23** | 0.15; 0.31 | <.001 | -0.03; 0.47 | 47.29 (<.001) | | | 0.13 | | | 79.56 | ref. group |
| Multiple | 4 | **0.27** | 0.22; 0.31 | <.001 | 0.19; 0.34 | 6.61 (.086) | | | 0.03 | | | 53.80 | .656 |
| Unspecified | 15 | **0.29** | 0.24; 0.35 | <.001 | 0.08; 0.48 | 137.69 (<.001) | | | 0.11 | | | 89.70 | .162 |
| **Eating disorders**  Any platform | 18 | **0.42** | 0.35; 0.48 | <.001 | 0.12; 0.64 | 168.04 (<.001) | | | 0.16 | | | 91.26 | .392 |
| Facebook | 4 | **0.50** | 0.30; 0.65 | <.001 | 0.03; 0.78 | 34.07 (<.001) | | | 0.23 | | | 92.37 | ref. group |
| Instagram | 7 | **0.42** | 0.33; 0.50 | <.001 | 0.17; 0.62 | 31.89 (<.001) | | | 0.13 | | | 83.28 | .422 |
| Unspecified | 6 | **0.37** | 0.24; 0.48 | <.001 | 0.04; 0.62 | 93.41  (<.001) | | | 0.16 | | | 94.07 | .172 |
| **Anxiety disorders**  Any platform | 13 | **0.31** | 0.18; 0.44 | <.001 | -0.20; 0.69 | 265.84 (<.001) | | | 0.26 | | | 97.02 | .107 |
| Facebook | 4 | **0.17** | 0.11; 0.22 | <.001 | 0.11; 0.22 | 1.97  (.579) | | | 0 | | | 0 | ref. group |
| Unspecified | 7 | **0.40** | 0.19; 0.58 | <.001 | -0.24; 0.80 | 225.88  (<.001) | | | 0.32 | | | 97.95 | .107 |
| **Social media addiction**  Unspecified | 5 | **0.34** | 0.26; 0.41 | <.001 | 0.18; 0.48 | 9.45 (.051) | | | 0.07 | | | 59.37 | n.a. |
| **Body dissatisfaction**  Any platform | 38 | **0.41** | 0.35; 0.47 | <.001 | -0.00; 0.70 | 924.86 (<.001) | | | 0.22 | | | 95.71 | .081 |
| Facebook | 9 | **0.34** | 0.22; 0.46 | <.001 | -0.05; 0.64 | 87.18  (<.001) | | | 0.19 | | | 92.78 | ref. group |
| Instagram | 8 | **0.49** | 0.38; 0.59 | <.001 | 0.14; 0.74 | 71.16 (<.001) | | | 0.19 | | | 91.56 | **.044** |
| Unspecified | 17 | **0.37** | 0.29; 0.43 | <.001 | 0.06; 0.61 | 200.26 (<.001) | | | 0.16 | | | 92.74 | .754 |
| **Negative affect**  Any platform | 9 | **0.31** | 0.19; 0.43 | <.001 | -0.08; 0.62 | 118.59  (<.001) | | | 0.19 | | | 92.68 | n.a. |
| Unspecified | 4 | **0.25** | 0.06; 0.42 | .011 | -0.17; 0.59 | 56.07  (<.001) | | | 0.19 | | | 94.40 |  |
| **Positive affect**  Any platform | 6 | -0.07 | -0.25; 0.12 | .461 | -0.51; 0.39 | 117.09 (<.001) | | | 0.23 | | | 96.17 | n.a. |
| Instagram | 4 | -0.07 | -0.35; 0.22 | .639 | -0.62; 0.53 | 116.32 (<.001) | | | 0.30 | | | 97.99 |  |
| **Life satisfaction**  Any platform | 13 | **-0.23** | -0.30; -0.14 | <.001 | -0.48; 0.05 | 135.29  (<.001) | | | 0.14 | | | 93.81 | .106 |
| Facebook | 4 | **-0.15** | -0.23; -0.06 | <.001 | -0.31; 0.03 | 14.11 (.003) | | | 0.08 | | | 75.93 | ref. group |
| Unspecified | 8 | **-0.28** | -0.38; -0.17 | <.001 | -0.54; 0.04 | 113.79 (<.001) | | | 0.16 | | | 95.62 | .106 |
| **Upward tendency or non-directional tendency – female samples only** | | | | | | | | | | | | | |
| **Any psycho-pathology**  Any platform | 22 | **0.41** | 0.32; 0.48 | <.001 | -0.02; 0.71 | 365.48  (<.001) | | | 0.22 | | | 95.06 | .683 |
| Facebook | 4 | **0.38** | 0.09; 0.61 | <.001 | -0.27; 0.79 | 38.02 (<.001) | | | 0.31 | | | 94.37 | ref. group |
| Instagram | 7 | **0.46** | 0.40; 0.52 | <.001 | 0.31; 0.59 | 18.76 (.005) | | | 0.08 | | | 63.53 | .500 |
| Unspecified | 9 | **0.38** | 0.22; 0.54 | <.001 | -0.19; 0.77 | 292.91 (<.001) | | | 0.29 | | | 97.88 | .967 |
| **Depression**  Any platform | 6 | **0.28** | 0.15; 0.40 | <.001 | -0.06; 0.56 | 51.28  (<.001) | | | 0.16 | | | 91.97 | n.a. |
| **Eating disorders**  Any platform | 13 | **0.42** | 0.35; 0.49 | <.001 | 0.18; 0.62 | 62.86 (<.001) | | | 0.13 | | | 85.33 | n.a. |
| Instagram | 6 | **0.45** | 0.38; 0.51 | <.001 | 0.30: 0.57 | 15.14 (.010) | | | 0.08 | | | 63.44 |  |
| **Anxiety disorders**  Any platform | 4 | **0.37** | 0.05; 0.63 | .024 | -0.35; 0.82 | 122.31  (<.001) | | | 0.34 | | | 97.68 | n.a. |
| **Body dissatisfaction**  Any platform | 22 | **0.38** | 0.31; 0.45 | <.001 | 0.01; 0.66 | 245.99  (<.001) | | | 0.19 | | | 91.95 | 0.223 |
| Facebook | 7 | **0.29** | 0.16; 0.42 | <.001 | -0.08; 0.60 | 49.65 (<.001) | | | 0.18 | | | 88.55 | ref. group |
| Instagram | 6 | **0.46** | 0.31; 0.59 | <.001 | 0.05; 0.74 | 60.29  (<.001) | | | 0.21 | | | 92.68 | .085 |
| Unspecified | 7 | **0.39** | 0.25; 0.51 | <.001 | -0.00; 0.68 | 92.23 (<.001) | | | 0.20 | | | 93.49 | .322 |
| **Upward tendency or non-directional tendency – young people (sample mean age ≤ 21 years)** | | | | | | | | | | | | | |
| **Any psycho-pathology**  Any platform | 25 | **0.34** | 0.29; 0.39 | <.001 | 0.08; 0.56 | 336.79  (<.001) | | | 0.14 | | | 94.20 | n.a. |
| Unspecified | 20 | **0.35** | 0.29; 0.41 | <.001 | 0.07; 0.59 | 315.51 (<.001) | | | 0.15 | | | 95.00 |  |
| **Depression**  Any platform | 15 | **0.30** | 0.27; 0.34 | <.001 | 0.17; 0.42 | 79.87  (<.001) | | | 0.07 | | | 81.05 | n.a. |
| Unspecified | 11 | **0.31** | 0.27; 0.36 | <.001 | 0.16; 0.45 | 69.46 (<.001) | | | 0.08 | | | 82.95 |  |
| **Eating disorders**  Any platform | 6 | **0.38** | 0.25; 0.48 | <.001 | 0.05; 0.63 | 88.63  (<.001) | | | 0.16 | | | 93.56 | n.a. |
| Unspecified | 5 | **0.38** | 0.23; 0.51 | <.001 | 0.02; 0.66 | 87.39 (<.001) | | | 0.18 | | | 95.31 |  |
| **Anxiety disorders**  Any platform | 5 | **0.28** | 0.23; 0.32 | <.001 | 0.20; 0.35 | 8.35  (.080) | | | 0.04 | | | 51.04 | n.a. |
| Unspecified | 4 | **0.25** | 0.22; 0.29 | <.001 | 0.21; 0.29 | 2.64 (.451) | | | 0.01 | | | 2.62 |  |
| **Body dissatisfaction**  Any platform | 19 | **0.34** | 0.27; 0.41 | <.001 | 0.00; 0.61 | 257.01  (<.001) | | | 0.17 | | | 94.60 | .715 |
| Facebook | 7 | **0.31** | 0.17; 0.44 | <.001 | -0.09; 0.63 | 69.19 (<.001) | | | 0.19 | | | 92.78 | ref. group |
| Unspecified | 10 | **0.34** | 0.25; 0.43 | <.001 | 0.01; 0.61 | 152.19 (<.001) | | | 0.17 | | | 94.56 | .715 |
| **Life satisfaction**  Any platform | 6 | **-0.30** | -0.39; -0.21 | <.001 | -0.51; - 0.06 | 50.52  (<.001) | | | 0.12 | | | 93.62 | n.a. |
| Unspecified | 5 | **-0.31** | -0.42; -0.21 | <.001 | -0.54; -0.05 | 48.95 (<.001) | | | 0.13 | | | 94.13 |  |
| **Upward tendency or non-directional tendency – high quality studies (quality sum score ≥ 7/8)** | | | | | | | | | | | | | |
| **Any psycho-pathology**  Any platform | 16 | **0.34** | 0.25; 0.42 | <.001 | -0.04; 0.63 | 230.08  (<.001) | | | 0.19 | | | 94.73 | n.a. |
| Unspecified | 10 | **0.33** | 0.20; 0.45 | <.001 | -0.11; 0.66 | 203.03  (<.001) | | | 0.22 | | | 96.89 |  |
| **Depression**  Any platform | 8 | **0.27** | 0.19; 0.35 | <.001 | 0.03; 0.48 | 52.27  (<.001) | | | 0.12 | | | 88.61 | n.a. |
| Unspecified | 5 | **0.25** | 0.14; 0.36 | <.001 | -0.03; 0.50 | 44.90  (<.001) | | | 0.13 | | | 93.58 |  |
| **Appearance-related upward tendency or non-directional tendency** | | | | | | | | | | | | | |
| **Any psycho-pathology**  Any platform | 22 | **0.42** | 0.35; 0.48 | <.001 | 0.08; 0.67 | 299.44  (<.001) | | | 0.18 | | | 93.88 | .566 |
| Facebook | 4 | **0.50** | 0.30; 0.65 | <.001 | 0.03; 0.78 | 34.07  (<.001) | | | 0.23 | | | 92.37 | ref. group |
| Instagram | 8 | **0.43** | 0.35; 0.51 | <.001 | 0.19; 0.63 | 37.18  (<.001) | | | 0.13 | | | 82.51 | .560 |
| Unspecified | 8 | **0.39** | 0.25; 0.51 | <.001 | -0.05; 0.70 | 158.45  (<.001) | | | 0.22 | | | 96.44 | .293 |
| **Eating disorders only**  Any platform | 16 | **0.43** | 0.36; 0.49 | <.001 | 0.16; 0.64 | 116.91  (<.001) | | | 0.15 | | | 89.79 | .555 |
| Facebook | 4 | **0.50** | 0.30; 0.65 | <.001 | 0.03; 0.78 | 34.07  (<.001) | | | 0.23 | | | 92.37 | ref. group |
| Instagram | 7 | **0.42** | 0.33; 0.50 | <.001 | 0.17; 0.62 | 31.89 (<.001) | | | 0.13 | | | 83.28 | .401 |
| Unspecified | 4 | **0.39** | 0.28; 0.50 | <.001 | 0.13; 0.61 | 38.76 (<.001) | | | 0.13 | | | 92.02 | .296 |
| **Body dissatisfaction**  Any platform | 36 | **0.41** | 0.34; 0.47 | <.001 | -0.02; 0.71 | 922.38  (<.001) | | | 0.23 | | | 95.55 | .074 |
| Facebook | 8 | **0.33** | 0.19; 0.46 | <.001 | -0.09; 0.65 | 77.38 (<.001) | | | 0.21 | | | 91.13 | ref. group |
| Instagram | 8 | **0.49** | 0.38; 0.59 | <.001 | 0.14; 0.74 | 71.16 (<.001) | | | 0.19 | | | 91.56 | **.039** |
| Unspecified | 16 | **0.36** | 0.29; 0.43 | <.001 | 0.04; 0.61 | 193.87 (<.001) | | | 0.17 | | | 92.77 | .421 |
| **Upward tendency only** | | | | | | | | | | | | | |
| **Any psycho-pathology**  Any platform | 21 | **0.32** | 0.25; 0.39 | <.001 | -0.03; 0.60 | 279.20  (<.001) | | | 0.18 | | | 95.17 | .729 |
| Facebook | 4 | **0.30** | 0.08; 0.50 | <.001 | -0.20; 0.68 | 50.14 (<.001) | | | 0.23 | | | 93.61 | ref. group |
| Unspecified | 15 | **0.34** | 0.25; 0.42 | <.001 | -0.02; 0.62 | 226.57 (<.001) | | | 0.18 | | | 96.12 | .729 |
| **Depression**  Any platform | 11 | **0.25** | 0.19; 0.31 | <.001 | 0.05; 0.43 | 56.91  (<.001) | | | 0.10 | | | 86.03 | n.a. |
| Unspecified | 7 | **0.26** | 0.18; 0.34 | <.001 | 0.04; 0.47 | 46.40  (.001) | | | 0.11 | | | 90.98 |  |
| **Body dissatisfaction**  Any platform | 7 | **0.54** | 0.36; 0.68 | <.001 | -0.02; 0.84 | 408.73  (<.001) | | | 0.30 | | | 97.65 | n.a. |
| Unspecified | 4 | **0.41** | 0.26; 0.55 | <.001 | 0.06; 0.67 | 31.35  (<.001) | | | 0.17 | | | 92.74 |  |
| **Life satisfaction**  Any platform | 5 | **-0.27** | -0.38; -0.15 | <.001 | -0.52; 0.03 | 54.91  (<.001) | | | 0.14 | | | 93.93 | n.a. |
| Unspecified | 4 | **-0.31** | -0.41; -0.20 | <.001 | -0.51; -0.07 | 42.04  (<.001) | | | 0.11 | | | 91.89 |  |
| **Non-directional tendency only** | | | | | | | | | | | | | |
| **Any psycho-pathology**  Any platform | 43 | **0.35** | 0.30; 0.40 | <.001 | -0.00; 0.63 | 535.69  (<.001) | | | 0.19 | | | 94.30 | .155 |
| Facebook | 13 | **0.29** | 0.18; 0.39 | <.001 | -0.10; 0.60 | 77.46 (<.001) | | | 0.20 | | | 89.67 | ref. group |
| Instagram | 8 | **0.43** | 0.35; 0.51 | <.001 | 0.19; 0.63 | 37.18  (<.001) | | | 0.13 | | | 82.51 | .052 |
| Multiple | 5 | **0.28** | 0.23; 0.32 | <.001 | 0.20; 0.36 | 9.20  (.056) | | | 0.04 | | | 53.66 | .912 |
| Unspecified | 17 | **0.38** | 0.29; 0.47 | <.001 | -0.03; 0.68 | 324.75 (<.001) | | | 0.21 | | | 96.27 | .151 |
| **Depression**  Any platform | 24 | **0.28** | 0.23; 0.32 | <.001 | 0.07; 0.46 | 148.43  (<.001) | | | 0.11 | | | 86.45 | .324 |
| Facebook | 11 | **0.24** | 0.15; 0.33 | <.001 | -0.05; 0.50 | 41.30 (<.001) | | | 0.14 | | | 81.66 | ref. group |
| Multiple | 4 | **0.27** | 0.22; 0.31 | <.001 | 0.19; 0.34 | 6.61 (.086) | | | 0.03 | | | 53.80 | .757 |
| Unspecified | 8 | **0.32** | 0.24; 0.39 | <.001 | 0.10; 0.50 | 66.48  (<.001) | | | 0.11 | | | 87.69 | .141 |
| **Eating disorders**  Any platform | 16 | **0.40** | 0.33; 0.47 | <.001 | 0.10; 0.63 | 151.15  (<.001) | | | 0.16 | | | 91.54 | .334 |
| Instagram | 7 | **0.42** | 0.33; 0.50 | <.001 | 0.17; 0.62 | 31.89 (<.001) | | | 0.13 | | | 83.28 | ref. group |
| Unspecified | 5 | **0.34** | 0.20; 0.47 | <.001 | -0.01; 0.62 | 91.38 (<.001) | | | 0.17 | | | 95.04 | .334 |
| **Anxiety**  Any platform | 12 | **0.32** | 0.18; 0.45 | <.001 | -0.21; 0.71 | 255.95  (<.001) | | | 0.27 | | | 97.06 | .080 |
| Facebook | 4 | **0.16** | 0.11; 0.22 | <.001 | 0.11; 0.22 | 1.97 (.579) | | | 0.00 | | | 0.00 | ref. group |
| Unspecified | 6 | **0.43** | 0.19; 0.63 | <.001 | -0.24; 0.83 | 200.71  (<.001) | | | 0.33 | | | 97.92 | .080 |
| **Body dissatisfaction**  Any platform | 31 | **0.38** | 0.32; 0.43 | <.001 | 0.03; 0.64 | 397.35 (<.001) | | | 0.18 | | | 93.73 | .076 |
| Facebook | 8 | **0.31** | 0.19; 0.43 | <.001 | -0.05; 0.60 | 70.10  (<.001) | | | 0.18 | | | 91.33 | ref. group |
| Instagram | 7 | **0.49** | 0.36; 0.60 | <.001 | 0.10; 0.75 | 70.10 (<.001) | | | 0.21 | | | 92.68 | **.033** |
| Unspecified | 13 | **0.35** | 0.27; 0.43 | <.001 | 0.04; 0.60 | 168.85  (.001) | | | 0.16 | | | 92.79 | .621 |
| **Negative affect**  Any platform | 6 | **0.33** | 0.17; 0.47 | <.001 | -0.08; 0.64 | 65.41  (<.001) | | | 0.20 | | | 91.80 | n.a. |
| **Positive affect**  Any platform | 5 | -0.10 | -0.31; 0.12 | .389 | -0.56; 0.41 | 106.33  (<.001) | | | 0.25 | | | 96.45 | n.a. |
| **Positive affect, outlier-corrected**  Any platform | 4 | **-0.19** | -0.35; -0.03 | .018 | n.a. | 22.77 (<.001) | | | n.a. | | | 90.36 | n.a. |
| **Life satisfaction**  Any platform | 8 | **-0.20** | -0.30; -0.10 | <.001 | -0.46; 0.09 | 68.28  (<.001) | | | 0.14 | | | 93.57 | n.a. |
| Unspecified | 4 | **-0.24** | -0.42; -0.04 | <.001 | -0.60; 0.20 | 58.87 (<.001) | | | 0.21 | | | 97.11 | n.a. |
| **Longitudinal bivariate correlations**  **Component of the social comparison process: Tendency** | | | | | | | | | | | | | |
| Outcome  platform-specific results | *k* | *r* | 95% CI | *p* | 95% PI | | *Q (p)* | | | *τ* | | *I*² | *p* mod. by platform |
| **Upward tendency or non-directional tendency** | | | | | | | | | | | | | |
| **Any psycho-pathology**  Any platform | 4 | **0.32** | 0.17; 0.46 | <.001 | -0.01; 0.59 | | 47.23 (<.001) | | | 0.16 | | 92.94 | n.a. |
| **Cross-sectional bivariate correlations**  **Component of the social comparison process: Self-evaluation** | | | | | | | | | | | | | |
| Outcome  platform-specific results | *k* | *r* | 95% CI | *p* | 95% PI | | *Q (p)* | | | *τ* | | *I*² | *p* mod. by platform |
| **Self-evaluation – upward or bipolar assessment (SC-based self-evaluation or social rank)** | | | | | | | | | | | | | |
| **Any psycho-pathology**  Any platform | 29 | **0.38** | 0.32; 0.45 | <.001 | 0.00; 0.67 | 385.94  (<.001) | | 0.20 | | | 93.19 | | .189 |
| Facebook | 6 | **0.37** | 0.30; 0.44 | <.001 | 0.21; 0.51 | 11.05  (.050) | | 0.08 | | | 55.17 | | ref. group |
| Instagram | 7 | **0.31** | 0.15; 0.45 | <.001 | -0.13; 0.65 | 59.67  (<.001) | | 0.21 | | | 90.67 | | .552 |
| Unspecified | 11 | **0.46** | 0.35; 0.56 | <.001 | 0.04; 0.74 | 183.29 (<.001) | | 0.22 | | | 94.48 | | .296 |
| **Depression**  Any platform | 18 | **0.34** | 0.28; 0.40 | <.001 | 0.09; 0.55 | 90.78  (<.001) | | 0.13 | | | 84.26 | | .348 |
| Facebook | 5 | **0.37** | 0.28; 0.45 | <.001 | 0.18; 0.54 | 11.05  (.026) | | 0.09 | | | 64.49 | | ref. group |
| Instagram | 5 | **0.27** | 0.20; 0.34 | <.001 | 0.15; 0.39 | 6.17 (.187) | | 0.05 | | | 34.79 | | .237 |
| Unspecified | 5 | **0.38** | 0.25; 0.50 | <.001 | 0.07; 0.63 | 27.66 (<.001) | | 0.15 | | | 89.89 | | .889 |
| **Eating disorders**  Any platform | 5 | **0.22** | 0.11; 0.33 | <.001 | -0.04; 0.46 | 22.37  (<.001) | | 0.12 | | | 85.51 | | n.a. |
| **Social media addiction**  Unspecified | 5 | **0.58** | 0.43; 0.70 | <.001 | 0.18; 0.82 | 55.56 (<.001) | | 0.22 | | | 92.85 | | n.a. |
| **Body dissatisfaction**  Any platform | 8 | **0.41** | 0.29; 0.52 | <.001 | 0.04; 0.68 | 125.44  (<.001) | | 0.19 | | | 92.98 | | n.a. |
| **Negative affect**  Any platform | 4 | **0.41** | 0.33; 0.48 | <.001 | 0.26; 0.54 | 13.08  (.005) | | 0.08 | | | 73.75 | | n.a. |
| **Life satisfaction**  Any platform | 9 | **-0.48** | -0.63; -0.29 | <.001 | -0.84; 0.19 | 370.72  (<.001) | | 0.34 | | | 98.84 | | n.a. |
| outlier-corrected | 8 | **-0.38** | -0.44; -0.33 | <.001 | -0.52; -0.23 | 45.87  (<.001) | | 0.08 | | | 83.51 | |  |
| Unspecified | 6 | **-0.51** | -0.72; -0.21 | <.001 | -0.90; 0.34 | 368.78 (<.001) | | 0.43 | | | 99.28 | |  |
| outlier-corrected | 5 | **-0.37** | -0.44; -0.29 | <.001 | -0.53; -0.18 | 38.73 (<.001) | | 0.09 | | | 88.26 | |  |
| **Self-evaluation – upward assessment or bipolar assessment (SC-based self-evaluation only)** | | | | | | | | | | | | | |
| **Any psycho-pathology**  Any platform | 20 | **0.44** | 0.36; 0.51 | <.001 | 0.06; 0.71 | 291.85  (<.001) | | 0.21 | | | 93.86 | | n.a. |
| Unspecified | 9 | **0.49** | 0.37; 0.60 | <.001 | 0.07; 0.77 | 157.64  (<.001) | | 0.23 | | | 94.60 | |  |
| **Depression**  Any platform | 11 | **0.38** | 0.30; 0.46 | <.001 | 0.10; 0.61 | 64.22  (<.001) | | 0.15 | | | 87.73 | | n.a. |
| Unspecified | 4 | **0.41** | 0.27; 0.54 | <.001 | 0.10; 0.65 | 16.03  (.001) | | 0.15 | | | 85.98 | |  |
| **Social media addiction**  Unspecified | 5 | **0.58** | 0.43; 0.70 | <.001 | 0.18; 0.82 | 55.56 (<.001) | | 0.22 | | | 92.85 | | n.a. |
| **Body dissatisfaction**  Any platform | 7 | **0.40** | 0.27; 0.53 | <.001 | 0.00; 0.69 | 114.46  (<.001) | | 0.20 | | | 93.81 | | n.a. |
| **Negative affect**  Any platform | 4 | **0.41** | 0.33; 0.48 | <.001 | 0.26; 0.54 | 13.08  (.005) | | 0.08 | | | 73.75 | | n.a. |
| **Life satisfaction**  Any platform | 8 | **-0.48** | -0.65; -0.26 | <.001 | -0.86; 0.24 | 370.15  (<.001) | | 0.37 | | | 99.05 | | n.a. |
| outlier-corrected | 7 | **-0.38** | -0.43; -0.32 | <.001 | -0.51; -0.22 | 43.06  (<.001) | | 0.08 | | | 84.66 | |  |
| Unspecified | 6 | **-0.51** | -0.72; -0.21 | <.001 | -0.90; 0.34 | 368.78 (<.001) | | 0.43 | | | 99.28 | |  |
| outlier-corrected | 5 | **-0.37** | -0.44; -0.29 | <.001 | -0.53; -0.18 | 38.73 (<.001) | | 0.09 | | | 88.26 | |  |
| **Self-evaluation – upward or bipolar assessment (SC-based self-evaluation or social rank) – female samples only** | | | | | | | | | | | | | |
| **Any psycho-pathology**  Any platform | 4 | **0.44** | 0.25; 0.60 | <.001 | 0.00; 0.74 | 23.64  (<.001) | | 0.21 | | | 89.06 | | n.a. |
| **Body dissatisfaction**  Any platform | 7 | **0.44** | 0.33; 0.54 | <.001 | 0.12, 0.68 | 52.29  (<.001) | | 0.17 | | | 88.11 | | n.a. |
| **Self-evaluation – upward or bipolar assessment (SC-based self-evaluation or social rank) – young people (sample mean age ≤ 21 years)** | | | | | | | | | | | | | |
| **Any psycho-pathology**  Any platform | 16 | **0.41** | 0.32; 0.50 | <.001 | 0.02; 0.69 | 253.00  (<.001) | | 0.21 | | | 93.01 | | n.a. |
| Unspecified | 9 | **0.46** | 0.32; 0.58 | <.001 | -0.01; 0.77 | 164.51 (<.001) | | 0.25 | | | 94.00 | |  |
| **Depression**  Any platform | 8 | **0.34** | 0.26; 0.42 | <.001 | 0.12; 0.53 | 26.82  (<.001) | | 0.11 | | | 78.10 | | n.a. |
| **Self-evaluation – upward or bipolar assessment (SC-based self-evaluation or social rank) – high quality studies only (quality sum score ≥ 7/8)** | | | | | | | | | | | | | |
| **Any psycho-pathology**  Any platform | 15 | **0.41** | 0.32; 0.50 | <.001 | 0.02; 0.70 | 226.70 (<.001) | | 0.21 | | | 94.20 | | n.a. |
| Unspecified | 8 | **0.50** | 0.37; 0.61 | <.001 | 0.08; 0.77 | 150.69 (<.001) | | 0.23 | | | 94.78 | |  |
| **Depression**  Any platform | 8 | **0.37** | 0.28; 0.45 | <.001 | 0.13; 0.56 | 26.59  (<.001) | | 0.12 | | | 80.14 | | n.a. |
| **Life satisfaction**  Any platform | 5 | **-0.53** | -0.76; -0.17 | .005 | -0.92; 0.40 | 368.56  (<.001) | | 0.47 | | | 99.38 | | n.a. |
| outlier-corrected | 4 | **-0.37** | -0.46; -0.26 | <.001 | -0.57; -0.12 | 38.68  (<.001) | | 0.12 | | | 91.77 | |  |
| **Social media addiction**  Unspecified | 4 | **0.60** | 0.41; 0.74 | <.001 | 0.14; 0.85 | 46.95 (<.001) | | 0.25 | | | 94.33 | | n.a. |
| **Appearance-related self-evaluation – upward or bipolar assessment (SC-based self-evaluation or social rank)** | | | | | | | | | | | | | |
| **Any psycho-pathology**  Any platform | 4 | **0.39** | 0.18; 0.57 | <.001 | -0.08; 0.72 | 27.99  (<.001) | | 0.22 | | | 91.73 | | n.a. |
| **Body dissatisfaction**  Any platform | 6 | **0.43** | 0.31; 0.53 | <.001 | 0.11; 0.67 | 95.57  (<.001) | | 0.16 | | | 91.28 | | n.a. |
| **Self-evaluation – upward assessment only** | | | | | | | | | | | | | |
| **Any psycho-pathology**  Any platform | 20 | **0.43** | 0.35; 0.51 | <.001 | 0.04; 0.71 | 300.70  (<.001) | | 0.21 | | | 94.15 | | .340 |
| Instagram | 4 | **0.42** | 0.23; 0.58 | <.001 | -0.01; 0.72 | 23.07  (<.001) | | 0.20 | | | 86.89 | | ref. group |
| Unspecified | 8 | **0.52** | 0.40; 0.62 | <.001 | 0.14; 0.77 | 145.09  (<.001) | | 0.21 | | | 94.17 | | .340 |
| **Depression**  Any platform | 12 | **0.37** | 0.29; 0.45 | <.001 | 0.09; 0.60 | 68.92  (<.001) | | 0.15 | | | 87.36 | | n.a. |
| **Social media addiction**  Unspecified | 5 | **0.58** | 0.43; 0.70 | <.001 | 0.18; 0.82 | 55.56  (<.001) | | 0.22 | | | 92.85 | | n.a. |
| **Negative affect**  Any platform | 4 | **0.41** | 0.33; 0.48 | <.001 | 0.26; 0.54 | 13.08  (.005) | | 0.08 | | | 73.75 | | n.a. |
| **Life satisfaction**  Any platform | 8 | **-0.48** | -0.65; -0.26 | <.001 | -0.86; 0.24 | 370.15  (<.001) | | 0.37 | | | 99.05 | | n.a. |
| outlier-corrected | 7 | **-0.38** | -0.43; -0.32 | <.001 | -0.51; -0.22 | 43.06  (<.001) | | 0.08 | | | 84.66 | |  |
| Unspecified | 6 | **-0.51** | -0.72; -0.21 | <.001 | -0.90; 0.34 | 368.75  (<.001) | | 0.43 | | | 99.28 | |  |
| outlier-corrected | 5 | **-0.37** | -0.44; -0.29 | <.001 | -0.53; -0.18 | 38.73 (<.001) | | 0.09 | | | 88.26 | |  |
| **Self-evaluation – downward assessment only** | | | | | | | | | | | | | |
| **Depression**  Any platform | 5 | 0.02 | -0.05; 0.08 | .572 | -0.05; 0.08 | 0.96  (.916) | | 0 | | | 0.00 | | n.a. |
| **Self-evaluation – bipolar assessment only (SC-based self-evaluation or social rank)** | | | | | | | | | | | | | |
| **Any psycho-pathology**  Any platform | 9 | **0.26** | 0.19; 0.34 | <.001 | 0.06; 0.45 | 30.30  (<.001) | | 0.10 | | | 72.39 | | n.a. |
| **Depression**  Any platform | 6 | **0.27** | 0.21; 0.33 | <.001 | 0.16; 0.37 | 7.65  (.177) | | 0.05 | | | 37.44 | | n.a. |
| **Body dissatisfaction**  Any platform | 5 | **0.40** | 0.26; 0.53 | <.001 | 0.05; 0.66 | 36.16  (<.001) | | 0.17 | | | 88.82 | | n.a. |
| **Self-evaluation – bipolar assessment only (SC-based self-evaluation)** | | | | | | | | | | | | | |
| **Body dissatisfaction**  Any platform | 4 | **0.38** | 0.20; 0.54 | <.001 | -0.03; 0.68 | 32.08  (<.001) | | 0.19 | | | 90.53 | | n.a. |
| **Self-evaluation – bipolar assessment only (social rank)** | | | | | | | | | | | | | |
| **Any psycho-pathology**  Any platform | 7 | **0.26** | 0.17; 0.34 | <.001 | 0.03; 0.46 | 26.14  (<.001) | | 0.11 | | | 76.38 | | n.a. |
| **Depression**  Any platform | 5 | **0.28** | 0.21; 0.34 | <.001 | 0.15; 0.39 | 7.17  (.127) | | 0.05 | | | 46.20 | | n.a. |
| **Cross-sectional bivariate correlations**  **Component of the social comparison process: Affective impact of social comparisons** | | | | | | | | | | | | | |
| Outcome  platform-specific results | *k* | *r* | 95% CI | *p* | 95% PI | | *Q (p)* | | | *τ* | | *I*² | *p* mod. by platform |
| **Affective impact to upward social comparisons** | | | | | | | | | | | | | |
| **Any psycho-pathology**  Any platform | 5 | **0.47** | 0.40; 0.53 | <.001 | 0.33; 0.58 | | 10.50 (.033) | | | 0.07 | | 64.31 | n.a. |
| **Standardized mean differences: Clinical vs. non-clinical sample (case control)**  **Component of the social comparison process: social comparison tendency** | | | | | | | | | | | | | |
| Outcome  platform-specific results | *k* | *g* | 95% CI | *p* | 95% PI | | *Q (p)* | | | *τ* | | *I*² | *p* mod. by platform |
| **Upward tendency** | | | | | | | | | | | | | |
| **Social comparison tendency** | 4 | **0.91** | 0.43; 1.39 | <.001 | -0.09; 1.91 | | 11.78  (.008) | | | 0.45 | | 89.25 | n.a. |

Abbreviations. *I^2^* = heterogeneity in outcomes in percent; *k* = number of independent effect sizes included in the given analysis; n.a. = not applicable; PI = Prediction Interval. **Bold** font indicates statistical significance at p < .050. Values are bivariate correlation coefficients (top) and Hedges’ g standardized mean differences (bottom) with 95% confidence intervals derived from the random effects meta-analysis.

^a^P-Values for total moderator (mod.) analyses (“any platform” row) and P-Values of all contrasts relative to reference (ref.) group, respectively. The moderator analyses examine the categorical variable social media platform as a potential moderator. This moderator analysis was conducted only when at least two platforms /platform categories presented with sufficient data (i.e., k ≥ 4). “Multiple” means that the measure assessed social comparison tendency for at least two specified platforms (e.g., Facebook and Instagram). “Unspecified” means that the measure assessed social comparison tendency for social media platforms without any specification (e.g., tendency generally on any applicable social media or social networking sites).

^b^Measures assessing non-directional social comparison tendency on social media either assessed tendency without restricting it to a specific direction or by combining multiple directions and reporting results across directions. ^c^Sub-analysis for measures that assessed social comparison tendency for multiple platforms (e.g., Facebook & Instagram).

^d^Sub-analysis for measures that assessed social comparison tendency but did not specify the social media platform(s) (e.g., referring to “social media” or “social networking sites” broadly).

**Appendix D – Social media platform-specific meta-analytic results for experimental studies assessing the effects of social comparison on social media on mental health outcomes**

| Outcome | *k* | SMD (*g*) | 95% CI | *p* | 95% PI | *Q (p)* | *τ* | *I*² | *p*^a^ mod. by platform |
| --- | --- | --- | --- | --- | --- | --- | --- | --- | --- |
| **Upward social comparison vs. control (e.g., landscape pictures)** | | | | | | | | | |
| **Negative affect**  Any platform  Instagram  TikTok | 21  13  7 | **0.33**  **0.28**  **0.45** | 0.23; 0.43  0.16; 0.41  0.26; 0.64 | < .001  <.001  <.001 | 0.04; 0.61  -0.00; 0.57  0.08; 0.82 | 32.67  (.037)  17.54  (.130)  10.71  (.098) | 0.14  0.13  0.16 | 35.74  30.98  41.61 | .148  ref. group  .148 |
| **Body dissatisfaction**  Any platform  Instagram  TikTok | 38  24  12 | **0.21**  **0.23**  **0.24** | 0.15; 0.28  0.16; 0.31  0.14; 0.34 | < .001  <.001  <.001 | 0.03; 0.39  0.16; 0.31  0.14; 0.34 | 39.74  (.349)  15.65  (.870)  12.33  (.339) | 0.09  0.00  0.00 | 17.89  0.00  0.01 | .921  ref. group  .921 |
| **Positive affect**  Any platform  Instagram  TikTok | 10  4  4 | **-0.29**  **-0.39**  **-0.37** | -0.43; -0.15  -0.56; -0.21  -0.55; -0.19 | <.001  <.001  <.001 | -0.55; -0.04  -0.58; -0.20  -0.55; -0.19 | 12.86  (.169)  2.89  (.409)  0.32  (.957) | 0.11  0.04  0.00 | 24.91  3.66  0.00 | .888  ref. group  .888 |
| **Upward social comparison vs. control - female samples** | | | | | | | | | |
| **Negative affect**  Any platform  Instagram  TikTok | 19  13  6 | **0.34**  **0.28**  **0.46** | 0.22; 0.45  0.16; 0.41  0.22; 0.70 | <.001  <.001  <.001 | 0.00; 0.67  -0.00; 0.57  -0.03; 0.95 | 30.64  (.032)  17.54  (.130)  10.70  (.058) | 0.16  0.13  0.22 | 39.48  30.98  53.18 | .174  ref. group  .174 |
| **Body dissatisfaction**  Any platform  Instagram  TikTok | 29  18  9 | **0.22**  **0.26**  **0.25** | 0.15; 0.30  0.17; 0.35  0.11; 0.38 | <.001  <.001  <.001 | 0.01; 0.44  0.17; 0.35  0.04; 0.46 | 33.47  (.219)  10.20  (.895)  10.62  (.224) | 0.10  0.00  0.08 | 24.43  0.00  16.89 | .843  ref. group  .843 |
| **Positive affect**  Any platform | 7 | **-0.28** | -0.43; -0.12 | <.001 | -0.43; -0.12 | 5.92  (.433) | 0.00 | 0.00 | n.a. |
| **Upward social comparison vs. control – young people (sample mean age ≤ 21)** | | | | | | | | | |
| **Negative affect**  Any platform  Instagram | 10  7 | **0.39**  **0.38** | 0.27; 0.51  0.23; 0.53 | <.001  <.001 | 0.27; 0.51  0.16; 0.61 | 8.89  (.448)  7.42  (.284) | 0.00  0.09 | 0.00  17.36 | n.a. |
| **Body dissatisfaction**  Any platform  Instagram  TikTok | 21  14  7 | **0.29**  **0.27**  **0.31** | 0.21; 0.37  0.17; 0.37  0.17; 0.44 | <.001  <.001  <.001 | 0.21; 0.37  0.17; 0.37  0.17; 0.44 | 13.21  (.868)  6.80  (.912)  6.26  (.395) | 0.00  0.00  0.00 | 0.00  0.00  0.03 | .705  ref. group  .705 |
| **Positive affect**  Any platform | 4 | **-0.29** | -0.55; -0.03 | .030 | -0.76; 0.18 | 6.76  (.080) | 0.20 | 56.28 | n.a. |
| **Thinspiration vs. control** | | | | | | | | | |
| **Body dissatisfaction**  Any platform  Any platform, outlier-adjusted  Instagram | 7  6  5 | 0.17  **0.26**  **0.23** | -0.01; 0.35  0.11; 0.40  0.07; 0.39 | .062  <.001  .006 | -0.21; 0.55  0.11; 0.40  0.07; 0.39 | 12.34 (.055)  2.69 (.748)  1.92  (.751) | 0.17  0.00  0.00 | 50.35  0.00  0.00 | n.a. |
| **Fitspiration vs. control** | | | | | | | | | |
| **Negative affect**  Any platform | 5 | **0.44** | 0.24; 0.65 | <.001 | 0.11; 0.77 | 5.52  (.238) | 0.13 | 31.51 | n.a. |
| **Body dissatisfaction**  Any platform  Instagram  TikTok | 15  10  4 | **0.18**  **0.29**  0.08 | 0.05; 0.32  0.18; 0.40  -0.09; 0.26 | .007  <.001  .339 | -0.23; 0.60  0.11; 0.47  -0.09; 0.26 | 38.06  (<.001)  10.15  (.338)  1.74  (.628) | 0.20  0.07  0.00 | 59.56  14.41  0.00 | **.048**  ref. group  .048 |
| **Upward social comparison vs. downward social comparison** | | | | | | | | | |
| **Positive affect**  Any platform | 4 | -1.29 | -4.64; 2.06 | .451 | -8.76; 6.18 | 180.16  (<.001) | 3.41 | 99.59 | n.a. |
| **Thinspiration vs. fitspiration** | | | | | | | | | |
| **Body dissatisfaction**  Any platform | 4 | -0.08 | -0.30; 0.14 | .480 | -0.46; 0.30 | 6.31 (.098) | 0.16 | 51.94 | n.a. |
| **Positive body image manipulation vs. control** | | | | | | | | | |
| **Negative affect**  Any platform | 12 | 0.05 | -0.09; 0.18 | .514 | -0.27; 0.36 | 17.03  (.083) | 0.15 | 36.25 | n.a.* |
| **Body dissatisfaction**  Any platform | 19 | -0.07 | -0.19; 0.05 | .264 | -0.45; 0.32 | 36.07  (.007) | 0.19 | 50.42 | n.a.* |
| **Positive affect**  Any platform | 10 | **0.19** | 0.07; 0.31 | .002 | 0.07; 0.31 | 8.30  (.505) | 0.00 | 0.00 | n.a.* |
| **Upward social comparison vs. positive body image manipulation** | | | | | | | | | |
| **Negative affect**  Any platform | 21 | **0.16** | 0.05; 0.28 | .006 | -0.24; 0.56 | 43.00  (.002) | 0.19 | 53.96 | n.a.* |
| trim-and-fill-adjusted | 24 | **0.12** | 0.00; 0.23 | .048 | n.a. | 53.01  (<.001) | 0.21 | 57.38 | n.a.* |
| **Body dissatisfaction**  Any platform | 35 | **0.32** | 0.19; 0.46 | <.001 | -0.43; 1.08 | 177.12  (<.001) | 0.38 | 82.96 | n.a.* |
| **Positive affect**  Any platform | 17 | **-0.31** | -0.43; -0.19 | <.001 | -0.65; 0.03 | 27.33  (.038) | 0.16 | 43.33 | n.a.* |

Abbreviations. *I^2^* = heterogeneity in outcomes in percent; *k* = number of independent effect sizes included in the given analysis; n.a. = not applicable; PI = Prediction Interval. **Bold** font indicates statistical significance at p < .050. Values are Hedges’ g standardized mean differences with 95% confidence intervals derived from the random effects meta-analysis.

^a^P-Values for moderator analyses concerning social media platform as potential moderator. This moderator analysis was conducted only when at least two platforms/platform categories presented with sufficient data (i.e., k ≥ 4), otherwise “n.a.” is denoted for “not applicable”.
*The meta-analytic review of positive body image manipulations was exploratory, which is why moderator analyses were not applied

**Appendix E – Search strategy**

Search date: inception up to 28.04.2026

1) PsycInfo & Medline combined search via Ebscohost - all fields search with the following search terms:
   "social compar*" OR upward comparison* OR downward comparison* OR "lateral comparison*"
   Hits: 18 094
2) Web of Science - all fields search with the following search terms:
   "social compar*" OR upward comparison* OR downward comparison* OR "lateral comparison*"
   Hits: 16 123

**Appendix F – References of screened reviews**

1. Appel, Markus; Marker, Caroline; Gnambs, Timo (2020): Are social media ruining our lives? A review of meta-analytic evidence. In *Review of General Psychology* 24 (1), pp. 60–74.
2. Arigo, Danielle; Bercovitz, Iris; Lapitan, Emmanuel; Gular, Sofia (2024): Social Comparison and Mental Health. In *Curr Treat Options Psych* 11 (2), pp. 17–33. DOI: 10.1007/s40501-024-00313-0.
3. Behera, Naresh; Khuntia, Sipra; Pandey, Kavita; Shankar, Shail (2025): Impact of Social Media Use on Physical, Mental, Social, and Emotional Health, Sleep Quality, Body Image, and Mood: Evidence from 21 Countries-A Systematic Literature Review with Narrative Synthesis. In *International journal of behavioral medicine*. DOI: 10.1007/s12529-025-10411-9.
4. Bonfanti, Rubinia Celeste; Melchiori, Francesco; Teti, Arianna; Albano, Gaia; Raffard, Stéphane; Rodgers, Rachel; Lo Coco, Gianluca (2025): The association between social comparison in social media, body image concerns and eating disorder symptoms: A systematic review and meta-analysis. In *Body Image* 52, p. 101841.
5. Bottaro, Rossella; Faraci, Palmira (2022): The Use of Social Networking Sites and Its Impact on Adolescents' Emotional Well-Being: a Scoping Review. In *Current addiction reports* 9 (4), pp. 518–539. DOI: 10.1007/s40429-022-00445-4.
6. Carraturo, Fabio; Di Perna, Tiziana; Giannicola, Viviana; Nacchia, Marco Alfonso; Pepe, Marco; Muzii, Benedetta et al. (2023): Envy, Social Comparison, and Depression on Social Networking Sites: A Systematic Review. In *European journal of investigation in health, psychology and education* 13 (2), pp. 364–376. DOI: 10.3390/ejihpe13020027.
7. Danthinne, Elisa S.; Giorgianni, Francesca E.; Rodgers, Rachel F. (2020): Labels to prevent the detrimental effects of media on body image: A systematic review and meta-analysis. In *The International journal of eating disorders* 53 (5), pp. 377–391. DOI: 10.1002/eat.23242.
8. Faelens, Lien; Hoorelbeke, Kristof; Cambier, Ruben; van Put, Jill; van de Putte, Eowyn; Raedt, Rudi de; Koster, Ernst H.W. (2021): The relationship between Instagram use and indicators of mental health: A systematic review. In *Comput Hum Behav Rep* 4, p. 100121. DOI: 10.1016/j.chbr.2021.100121.
9. Fioravanti, Giulia; Bocci Benucci, Sara; Ceragioli, Giulia; Casale, Silvia (2022): How the Exposure to Beauty Ideals on Social Networking Sites Influences Body Image: A Systematic Review of Experimental Studies. In *Adolescent Res Rev* 7 (3), pp. 419–458. DOI: 10.1007/s40894-022-00179-4.
10. Goldberg, Ximena (2023): Social comparison links social media use and well-being. In *Nature Reviews Psychology* 2 (11), p. 658. DOI: 10.1038/s44159-023-00244-2.
11. Haidt, J.; Rausch, Z.; Twenge, J. (ongoing): Social media and mental health: A collaborative review. New York University. Available online at tinyurl.com/SocialMediaMentalHealthReview.
12. Holland, Grace; Tiggemann, Marika (2016): A systematic review of the impact of the use of social networking sites on body image and disordered eating outcomes. In *Body Image* 17, pp. 100–110. DOI: 10.1016/j.bodyim.2016.02.008.
13. Kim, Bo Ra; Mackert, Michael (2022): Social media use and binge eating: An integrative review. In *Public health nursing (Boston, Mass.)* 39 (5), pp. 1134–1141. DOI: 10.1111/phn.13069.
14. Kostyrka-Allchorne, Katarzyna; Stoilova, Mariya; Bourgaize, Jake; Rahali, Miriam; Livingstone, Sonia; Sonuga-Barke, Edmund (2023): Review: Digital experiences and their impact on the lives of adolescents with pre-existing anxiety, depression, eating and nonsuicidal self-injury conditions - a systematic review. In *Child and adolescent mental health* 28 (1), pp. 22–32. DOI: 10.1111/camh.12619.
15. Kross, Ethan; Verduyn, Philippe; Sheppes, Gal; Costello, Cory K.; Jonides, John; Ybarra, Oscar (2021): Social Media and Well-Being: Pitfalls, Progress, and Next Steps. In *Trends in cognitive sciences* 25 (1), pp. 55–66. DOI: 10.1016/j.tics.2020.10.005.
16. Lei, Yuqing; Hu, Shirui; Sun, Yidan; Zheng, Lijun (2026): "Looking up" linked to feeling down: a meta-analysis of online upward social comparison and psychological maladjustment. In *Front Psychol* 17, p. 1825169. DOI: 10.3389/fpsyg.2026.1825169.
17. Marciano, Laura; Lin, Jeffrey; Sato, Taisuke; Saboor, Sundas; Viswanath, Kasisomayajula (2024): Does social media use make us happy? A meta-analysis on social media and positive well-being outcomes. In *SSM - Mental Health* 6, p. 100331. DOI: 10.1016/j.ssmmh.2024.100331.
18. Marciano, Laura; Ostroumova, Michelle; Schulz, Peter Johannes; Camerini, Anne-Linda (2021): Digital Media Use and Adolescents' Mental Health During the Covid-19 Pandemic: A Systematic Review and Meta-Analysis. In *Front Public Health* 9, p. 793868. DOI: 10.3389/fpubh.2021.793868.
19. McCarthy, Peter A.; Morina, Nexhmedin (2020): Exploring the association of social comparison with depression and anxiety: A systematic review and meta-analysis. In *Clinical psychology & psychotherapy* 27 (5), pp. 640–671. DOI: 10.1002/cpp.2452.
20. McComb, Carly A.; Vanman, Eric J.; Tobin, Stephanie J. (2023): A Meta-Analysis of the Effects of Social Media Exposure to Upward Comparison Targets on Self-Evaluations and Emotions. In *Media Psychology* 26 (5), pp. 612–635. DOI: 10.1080/15213269.2023.2180647.
21. Meier, Adrian; Reinecke, Leonard (2021): Computer-mediated communication, social media, and mental health: A conceptual and empirical meta-review. In *Communication Research* 48 (8), pp. 1182–1209.
22. Morina, Nexhmedin; Lemmel, Frederike K.; Schlechter, Pascal; Hoppen, Thole H. (2026): Unraveling Social Comparison in Mental Health: A Process-Based Model and a Systematic Review and Meta-Analysis of Current Evidence. In *Clinical Psychological Science*, 21677026261442164.
23. Myers, Taryn A.; Crowther, Janis H. (2009): Social comparison as a predictor of body dissatisfaction: A meta-analytic review. In *Journal of Abnormal Psychology* 118 (4), pp. 683–698. DOI: 10.1037/a0016763.
24. Orben, Amy; Meier, Adrian; Dalgleish, Tim; Blakemore, Sarah-Jayne (2024): Mechanisms linking social media use to adolescent mental health vulnerability. In *Nature Reviews Psychology* 3 (6), pp. 407–423.
25. Panayiotou, Margarita; Black, Louise; Carmichael-Murphy, Parise; Qualter, Pamela; Humphrey, Neil (2023): Time spent on social media among the least influential factors in adolescent mental health: preliminary results from a panel network analysis. In *Nat. Mental Health* 1 (5), pp. 316–326. DOI: 10.1038/s44220-023-00063-7.
26. Saiphoo, Alyssa N.; Vahedi, Zahra (2019): A meta-analytic review of the relationship between social media use and body image disturbance. In *Comput Hum Behav* 101, pp. 259–275. DOI: 10.1016/j.chb.2019.07.028.
27. Sarmiento, Irene G.; Olson, Chelsea; Yeo, GeckHong; Chen, Y. Anthony; Toma, Catalina L.; Brown, B. Bradford et al. (2020): How Does Social Media Use Relate to Adolescents’ Internalizing Symptoms? Conclusions from a Systematic Narrative Review. In *Adolescent Res Rev* 5 (4), pp. 381–404. DOI: 10.1007/s40894-018-0095-2.
28. So, Bohee; Kwon, Ki Han (2023): The Impact of Thin-Ideal Internalization, Appearance Comparison, Social Media Use on Body Image and Eating Disorders: A Literature Review. In *Journal of Evidence-Based Social Work* 20 (1), pp. 55–71. DOI: 10.1080/26408066.2022.2117582.
29. Turk, Fidan; Bozdogan, Aysesu; Hughes, Rachel; Longhurst, Phaedra; Chilvers, Millie; Fitzsimmons-Craft, Ellen E. (2026): Social Comparison and Its Association With Disordered Eating Symptoms: A Systematic Review and Meta-Analysis. In *Int J Eat Disord* 59 (6), pp. 1117–1194. DOI: 10.1002/eat.70058.
30. Valkenburg, Patti M.; Meier, Adrian; Beyens, Ine (2022): Social media use and its impact on adolescent mental health: An umbrella review of the evidence. In *Current Opinion in Psychology* 44, pp. 58–68.
31. Verduyn, Philippe; Gugushvili, Nino; Massar, Karlijn; Täht, Karin; Kross, Ethan (2020): Social comparison on social networking sites. In *Current Opinion in Psychology* 36, pp. 32–37. DOI: 10.1016/j.copsyc.2020.04.002.
32. Vidal, Carol; Lhaksampa, Tenzin; Miller, Leslie; Platt, Rheanna (2020): Social media use and depression in adolescents: a scoping review. In *International review of psychiatry (Abingdon, England)* 32 (3), pp. 235–253. DOI: 10.1080/09540261.2020.1720623.
33. Webster, Deborah; Dunne, Laura; Hunter, Ruth (2021): Association Between Social Networks and Subjective Well-Being in Adolescents: A Systematic Review. In *Youth & Society* 53 (2), pp. 175–210. DOI: 10.1177/0044118X20919589.
34. Yang, Qinghua; Liu, Jiangmeng; Rui, Jian (2022): Association between social network sites use and mental illness: A meta-analysis. In *Cyberpsychology* 16 (1). DOI: 10.5817/CP2022-1-1.
35. Yoon, Sunkyung; Kleinman, Mary; Mertz, Jessica; Brannick, Michael (2019): Is social network site usage related to depression? A meta-analysis of Facebook-depression relations. In *Journal of Affective Disorders* 248, pp. 65–72. DOI: 10.1016/j.jad.2019.01.026.

**Appendix G – Study quality assessment**

Quality of Narrative Studies

A. Was SC measured with a validated measure of SC on SNS [social networking site]?

(2) SC was assessed with an instrument that has been validated to measure SC on SNS.

(1) Comparison was assessed with an instrument that has been validated to measure SC in a non-SNS context; or only SNS reliability data reported.

(0) SC was assessed with an instrument that has not been validated OR only internal consistency was reported OR insufficient information was provided.

B. Did the assessment of SC take one or more direction (i.e., upward, and/or downward, and/or lateral) into account and were results reported per direction?

(2) The assessment of SC implied a clear direction of SC and authors report direction-specific results.

(0) The assessment of SC did not (clearly) take direction into account OR results not
 reported per direction OR insufficient information reported.

Only for within-effect sizes (i.e. when association between social comparison and another outcome is analysed):

C. Was the outcome measured with a validated measure?

(4) Outcome was assessed with an instrument that has been validated in an independent sample.

(2) Outcome was assessed with an instrument that has not been fully validated, but e.g., reliability was reported.

(0) Outcome was assessed with an instrument that has not been validated *or* only internal consistency was reported *or* insufficient information was provided.

Quality of Reaction Studies

A. Was exposure or manipulation of SC clearly related to one or more SC direction (i.e., upward, and/or downward, and/or lateral) and were results reported per direction?

(4) Exposure or manipulation of SC implied a clear direction of SC and results were reported per direction.

(2) Exposure or manipulation of SC implied a direction of SC that was likely perceived as such by the vast majority of participants.

(0) Exposure or manipulation of SC implied no clear direction of SC OR results were not reported per direction OR insufficient information supplied.

NOTE: This criterion A was not rated for studies applying a positive body image manipulations as these manipulations tend to use stimuli with varying SC directions.

B. Was the outcome (i.e., reaction) measured with a validated measure?

(4) Outcome was assessed with an instrument that has been validated in an independent sample.

(2) Outcome was assessed with an instrument that has not been fully validated, but e.g., reliability was reported.

(0) Outcome was assessed with an instrument that has not been validated *or* only internal consistency was reported *or* insufficient information was provided.

Quality of Selection Studies

A. How many SC direction choices (i.e., upward, downward, lateral) were available to study participants?

(4) All three choices: Upward, downward, and lateral.

(2) Two choices only: upward/downward; or lateral/upward; or lateral/downward.

(0) Insufficient information supplied.

NOTE: To the best of the authors knowledge, no selection study has been published yet. Thus, this quality criterion was not applied.
